# Normative transcriptomic clues to thalamic developmental vulnerability in *C9orf7*2-FTLD

**DOI:** 10.64898/2026.10.09.26365172

**Authors:** Isis So, Arabella Bouzigues, Eve Ferry-Bolder, Eva Cullen, Lucy L. Russell, Phoebe H. Foster, Sophie Farley, Rhian Convery, John C. van Swieten, Lize C. Jiskoot, Harro Seelaar, Daniela Galimberti, Rik Vandenberghe, Robert Laforce, Rose Bruffaerts, Maxime Bertoux, Thibaud Lebouvier, Eino Solje, Johannes Levin, Giuseppe di Fede, Alexander Thompson, Isabelle Le Ber, Raffaella Lara Migliaccio, Peter Körtvélyessy, Matthias L. Schroeter, Giancarlo Logroscino, Markus Otto, Zeljko Uzelac, Ignacio Illán-Gala, Johanna Krüger, Benedetta Nacmias, Alexander Gerhard, Tobias Langheinrich, Simon Ducharme, Isabel Santana, Maria Carmela Tartaglia, Mario Masellis, Alexandre de Mendonça, James B. Rowe, Caroline Graff, Fermin Moreno, Matthis Synofzik, Barbara Borroni, Raquel Sanchez-Valle, Rosa Rademakers, Danielle Brushaber, Leah K. Forsberg, Hilary W. Heuer, Kejal Kantarci, Tyler Kolander, Carly Mester, Bradley F. Boeve, Adam L. Boxer, Howard J. Rosen, Suzee E. Lee, Jonathan D. Rohrer, Tallulah S. Andrews, Elizabeth C. Finger, Frontotemporal Dementia Prevention Initiative (FPI) Investigators

## Abstract

*C9orf72* encodes a protein that is important for both typical neurodevelopment and homeostatic processes across the lifespan. Pathogenic *C9orf72* repeat expansions constitute the most common genetic cause of frontotemporal lobar degeneration (FTLD). Collectively, preclinical models and clinical neuroimaging studies have observed smaller thalamic volumes in *C9orf72*-associated FTLD, including in asymptomatic young adult *C9orf72* repeat expansion carriers across 18 to 30 years old, supportive of potential neurodevelopmental origins. However, it is unknown whether asymptomatic young adult *C9orf72* expansion carriers demonstrate volume differences in specific thalamic nuclei, as reported in older asymptomatic carriers and symptomatic patients; if so, normative transcriptomic patterns could reveal relevant neurodevelopmental and/or FTLD-related neurodegenerative processes. This study investigated volumetric MRI differences of the thalamic nuclei in asymptomatic young adult *C9orf72* expansion carriers compared to non-carriers, and examined spatial associations with normative thalamic transcriptomics from 8 weeks post-conception through middle age.

Young adult *C9orf72* expansion carriers (*n*=72) and non-carriers (*n*=48) were enrolled in the Frontotemporal Dementia Prevention Initiative across Europe, Canada, and the USA (ages: 18-30 years; sex: 57% female). Spatial associations between young adult thalamic nuclei volumes and normative transcriptomics from the Allen Human Brain Atlas were examined using complementary data-driven and hypothesis-driven approaches. Spatial expression patterns of transcripts of interest were characterized across prenatal and postnatal neurodevelopment using BrainSpan data.

Smaller volumes of the pulvinar, mediodorsal, and lateral nuclei were observed in young adult *C9orf72* expansion carriers compared to non-carriers. These nuclei showed higher normative expression of transcripts involved concurrently in neurodevelopmental and *C9orf72*-associated FTLD neurodegenerative processes (including lysosomal, autophagic, and small GTPase functions), and lower normative expression of *CALB1*, a marker of prenatal human thalamic development. Normative expression of *CALB2* (another human thalamic development marker) yielded positive correlations with *C9orf72* across prenatal development to adulthood.

These findings identify candidate transcripts that may provide insight into the molecular architecture leading to smaller thalamic nuclei volumes in *C9orf72*-associated FTLD.

## Introduction

Frontotemporal lobar degeneration (FTLD) is a neurodegenerative dementia that demonstrates heterogeneous age of onset, clinical symptomology, and neuropathology.^1^ Around 10-15% of all FTLD cases are caused by autosomal dominant variants, with 13-50% caused by hexanucleotide repeat expansions in *C9orf72*.^2,3^ Both fundamental research on *C9orf72*^4^ and preclinical models of *C9orf72-*associated FTD/amyotrophic lateral sclerosis in neurodevelopmental contexts^5–7^ have shown that *C9orf72* is important for typical neurodevelopmental processes, including regulating cortical and thalamic volume development, neural stem cell proliferation, self-renewal, and maintenance, neuronal survival, synaptogenesis, and axonogenesis. The question of whether *C9orf72* repeat expansions may affect neurodevelopment in FTLD is thus being increasingly explored,^8–14^ with efforts underway to test this hypothesis in human youth carriers of genetic FTLD variants.^15,16^

In asymptomatic young adult carriers of pathogenic *C9orf72* repeat expansions aged 18-30 years, arguably the last window of neurodevelopment where synaptic pruning, myelination, and maturation processes remain prominent,^17^ clinical neuroimaging studies have consistently observed smaller thalamic volumes at the group level.^8,10,11^ The earliest average age at which smaller thalamic volumes have been detected in young adult expansion carriers compared to non-carriers was 26 years old.^10^ Longitudinal modelling of cortical and subcortical volumes in young adult *C9orf72* expansion carriers across two international FTLD cohorts extended these findings, and did not find an interaction with with age across 18 to 30 years old, supporting the hypothesis that thalamic volume differences may have neurodevelopmental origins.^8^ Studies of slightly older asymptomatic *C9orf72* expansion carriers (aged 39.8±11.1 years) found differences in average gray matter volumes, white matter integrity, and neuropsychological test scores,^18^ as well as smaller volumes at the level of specific thalamic nuclei, with earliest differences affecting the laterodorsal, lateral geniculate, and pulvinar nuclei in carriers aged 43.9±11.7 years.^19^ A natural hypothesis arising from both preclinical and clinical neuroimaging literature, then, is that if volume differences within specific thalamic nuclei are present in young adult *C9orf72* expansion carriers, the potential underlying cellular and molecular mechanisms could reflect neurodevelopmental and/or FTLD-related neurodegenerative processes.

As the thalamus is reliably the earliest region where volumetric differences are observed in *C9orf72*-associated FTLD,^8,10,11^ understanding its early developmental characteristics may provide a framework for investigating mechanisms contributing to *C9orf72*-associated FTLD vulnerability and resilience. Human thalamic development primarily occurs during the first and second prenatal trimesters, with specific thalamic nuclei becoming discernible between 8-16 gestational weeks.^20^ Distinct molecular profiles of the developing human thalamus highlight that the involved processes are tightly spatially- and temporally-regulated.^21^ Thalamic neurogenesis is prominent during the first trimester, typically characterized by radial glia (neural stem cell) markers, including *PAX6*, *OLIG3*, and Wnt family and sonic hedgehog signaling ligands, related transcription factors, co-receptors, and feedback regulators.^21^ Differentiation of thalamic neurons is prominent during the second trimester, yielding markers of glutamatergic (e.g.: *LHX9*, *LHX2*, *SLC17A6, S100A6, SOX2, FOXP2, CALB1, PVALB, NECAB1, CALB2*), and GABAergic (e.g.: *GAD1, SLC32A1, OTX2, SOX14, FOXG1, CRABP1*) neurons, as well as glial progenitors, terminally-differentiated glial cells, and neurovascular cells.^21^ Examining correlates of thalamic neurodevelopment in *C9orf72*-associated FTLD thus requires multimodal genetics, transcriptomics, and neuroimaging data aligned across critical periods of human neurodevelopment.

In adult-onset neurodegenerative disorders, one approach is to examine spatial associations between human brain transcriptomics from healthy donors and neuroimaging data from disease cohorts.^21,22^ To examine the potential pathways and mechanisms that may predispose young adult *C9orf72* repeat expansion carriers to smaller thalamic volumes, this study investigated normative human brain transcriptomic patterns in thalamic nuclei of young adult *C9orf72* repeat expansion carriers. The first objective examined associations between normative transcriptomic patterns in the thalamic nuclei of healthy adult Allen Human Brain Atlas (AHBA) donors and volume differences between thalamic nuclei in young adult FTLD-causing *C9orf72* repeat expansion carriers and familial non-carriers. A combination of data- and hypothesis-driven approaches were used to examine the normative AHBA RNA microarray data. Given prior findings of smaller laterodorsal, lateral geniculate, and pulvinar thalamic nuclei in asymptomatic carriers in their early forties, these nuclei were hypothesized to be smaller in young adult *C9orf72* repeat expansion carriers compared to familial non-carriers. In healthy adults from the AHBA, these nuclei were posited to show differential spatial expression of transcripts involved in neurodevelopment, synaptic plasticity, and neurodegeneration compared to thalamic nuclei which do not show volume differences between groups, and potentially reveal candidate markers and/or mechanisms of *C9orf72*-associated FTLD effects on neurodevelopment. The second objective was to delineate the spatial expression of transcripts of interest specific to FTLD or neurodevelopment, and their association with *C9orf72*, across normative prenatal and postnatal neurodevelopment using BrainSpan RNA sequencing data. Spatial expression patterns of these transcripts were examined in the thalamus and in the occipital cortex (a region not typically affected by FTLD-related neurodegenerative atrophy). Expression patterns of FTLD- and neurodevelopment-related genes were expected to differ between the thalamus and occipital cortex across normative developmental and early adulthood stages, and expression levels of different transcripts were expected to correlate with *C9orf72* transcript expression in the thalamus compared to the occipital cortex. The transcripts identified in these objectives would identify candidate contributors to neurodevelopmental and neurodegenerative effects of *C9orf72* repeat expansion in the thalamus.

## Materials and methods

### Study design and participants

This study included young adult *C9orf72* repeat expansion carriers and familial non-carrier participants in the Genetic Frontotemporal Dementia Initiative (GENFI: https://www.genfi.org/), or ARTFL-LEFFTLDS Longitudinal Frontotemporal Lobar Degeneration Study (ALLFTLD: https://www.allFTLD.org/, NCT04363684; ARTFL=Advancing Research and Treatment for Frontotemporal Lobar Degeneration study: NCT02365922; LEFFTLDS=Longitudinal Frontotemporal Lobar Degeneration study, NCT02372773). Phases 1-3 of GENFI comprised 46 clinical research sites across Europe and Canada (GENFI1, 2012-15; GENFI2, 2015-21; GENFI3, 2021-26), and ALLFTLD comprised 28 sites across Canada and the USA. Individual sites obtained ethics approval. All participants provided written informed consent, aligning with the Declaration of Helsinki.

The parent FTLD consortia studies are prospective, longitudinal, and observational in design, collecting demographics, neuroimaging, neuropsychological, behavioural, and clinical outcomes at 1-to-2-year intervals. Details, including full inclusion and exclusion criteria, have been previously reported.^22,23^ Specific inclusion criteria for this study included young adults, aged 18-30 years, who were carriers or familial non-carriers of >30 repeat expansions in *C9orf72*, from whom baseline T1-weighted volumetric MRI data was analyzed.

### Transcriptomic Data

Transcriptomics were obtained from the open-source AHBA RNA microarray and BrainSpan RNA sequencing datasets. The AHBA and BrainSpan data were derived from six adult donors aged 24 to 57 years, and 35 donors across prenatal and postnatal ages from 8 weeks post-conception to 40 years of age, respectively. Spatial neuroimaging-transcriptomic correlations were computed using volumetric MRI from young adult FTLD *C9orf72* repeat expansion carriers and non-carriers from the FPI, and normative RNA microarray data from adults in the AHBA, using complementary data-driven and hypothesis-driven approaches **(Figure 1)**. Normative BrainSpan RNA sequencing data was analyzed with the same list of hypothesis-driven genes to delineate their transcript expression profiles across human brain development.

**Figure 1.**
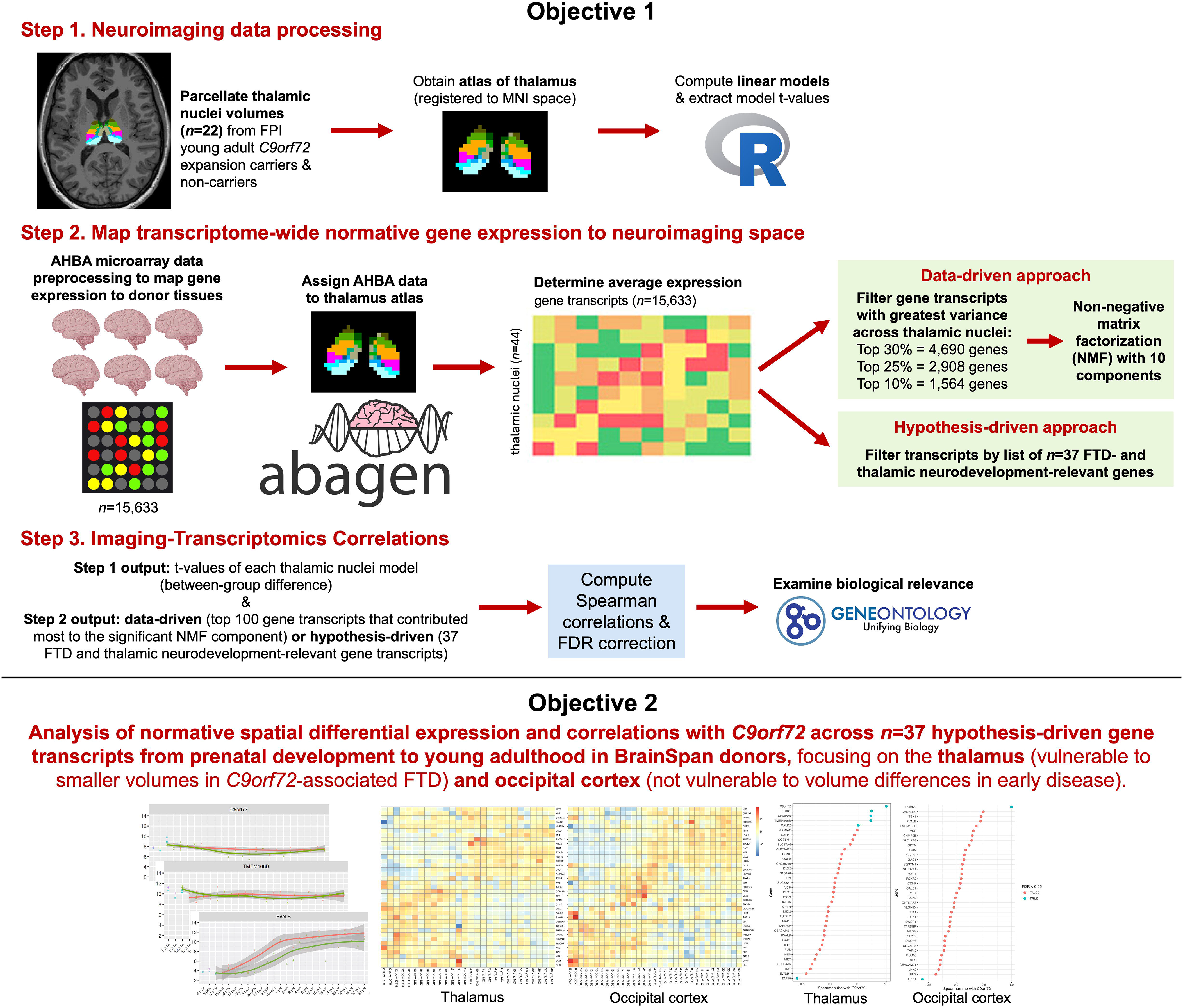
Graphical overview of the methodological pipeline. Data- and hypothesis-driven neuroimaging-transcriptomic correlations (top panel) and characterization of normative expression of hypothesis-driven gene transcripts across prenatal and postnatal development (bottom panel).

### Neuroimaging data acquisition and processing

GENFI and ALLFTLD neuroimaging protocols have been detailed elsewhere.^22,23^ GENFI employed T1-weighted MRI with a 1.5T (Siemens, GE) or 3T scanner (Siemens Trio, Siemens Skyra, Siemens Prisma, Philips Achieva, GE Discovery MR750) with the following sequence parameters: 256×256x208 matrix; 208 slices; 1.1mm isotropic voxel size; 8° flip angle; TR and TE varied by vendor. ALLFTLD employed T1-weighted MRI on 3T scanners (GE, Philips, Siemens) using these parameters: 240×256×256 matrix; 170 slices; voxel size=1.05×1.05×1.25 mm^3^; flip angle, TR, and TE varied by vendor.

Quality control and visual inspection was conducted by the respective core imaging groups per consortia, with images affected by excessive movements or artifacts removed. Image preprocessing employed FreeSurfer v7.4.1,^24^ and subcortical volume segmentation involved Freesurfer’s probabilistic atlas, Aseg.^25^ Probabilistic subregion segmentation for the thalamus was conducted per Iglesias *et al*.^26^ Twenty-two nuclei per hemisphere were retained for analysis; four nuclei that were not successfully segmented in all participants (ventromedial, paracentral, paratenial, reticular) were not analyzed. These nuclei are relatively small and have less distinct anatomical boundaries, which can lead to inconsistent labelling at the individual subject level.^26^

A representative individual thalamic nuclei atlas, which belonged to a non-carrier participant, was used for the spatial assignment of AHBA microarray expression data. Construction of a group atlas using all participant data led to the loss of several key nuclei subregions, which prompted the statistical determination of a representative atlas instead.^27^ This involved calculating Euclidean distances from the cohort mean from a participant-by-nucleus volume matrix, where the participant with the smallest distance to the mean was selected as the representative atlas.^27^

### Transcriptomic data acquisition

Regional microarray expression data used for the first objective of this study were obtained from 6 post-mortem brains (1 female, age [range] 24.0-57.0 years, [mean±SD] 42.50±13.38 years) provided by the AHBA (https://human.brain-map.org/).^28^ Data were processed with the abagen toolbox v0.1.3 (https://github.com/rmarkello/abagen) using a 44-region volumetric atlas in MNI space.

### Transcriptomic processing pipeline in Abagen

Microarray probes were reannotated using data provided by Arnatkevic iūtė et al^29^; probes not matched to a valid Entrez ID were discarded. Next, probes were filtered based on their expression intensity relative to background noise,^30^ such that probes with intensity less than the background in >=50.00% of samples across donors were discarded, yielding 31,569 probes. When multiple probes indexed the expression of the same gene, we selected and used the probe with the most consistent pattern of regional variation across donors (i.e., differential stability^31^), calculated with:

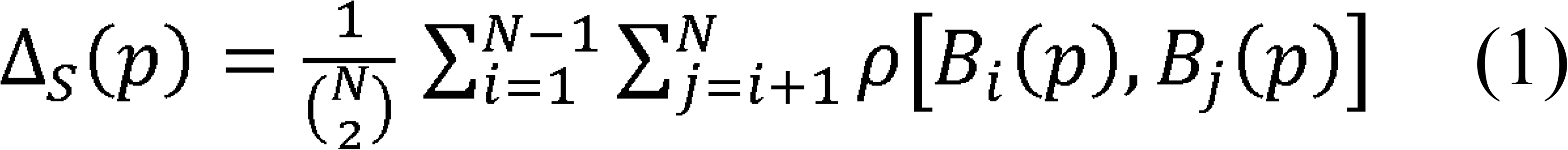

where is Spearman’s rank correlation of the expression of a single probe, p, across regions in two donors and, and N is the total number of donors. Here, regions correspond to the structural designations provided in the ontology from the AHBA.

The MNI coordinates of tissue samples were updated to those generated via non-linear registration using the Advanced Normalization Tools (ANTs; https://github.com/chrisfilo/alleninf). Samples were assigned to brain regions in the provided atlas if their MNI coordinates were within 2 mm of a given parcel. To reduce the potential for misassignment, sample-to-region matching was constrained by hemisphere and gross structural divisions (in this case, the “subcortex/brainstem” per abagen, such that e.g., a sample in the left subcortex could only be assigned to an atlas parcel in the left subcortex).^29^ If a brain region was not assigned a sample from any donor based on the above procedure, the tissue sample closest to the centroid of that parcel was identified independently for each donor. The average of these samples was taken across donors, weighted by the distance between the parcel centroid and the sample, to obtain an estimate of the parcellated expression values for the missing region. This procedure was performed for 22 regions that were not assigned tissue samples.

Inter-subject variation was addressed by normalizing tissue sample expression values across genes using a robust sigmoid function^32^:

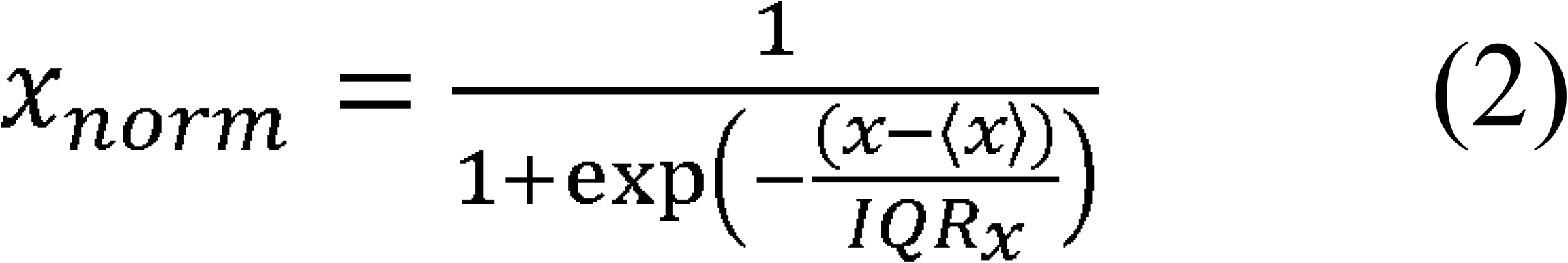

where 〈χ〉is the median and *IQR_χ_* is the normalized interquartile range of the expression of a single tissue sample across genes. Normalized expression values were then rescaled to the unit interval:

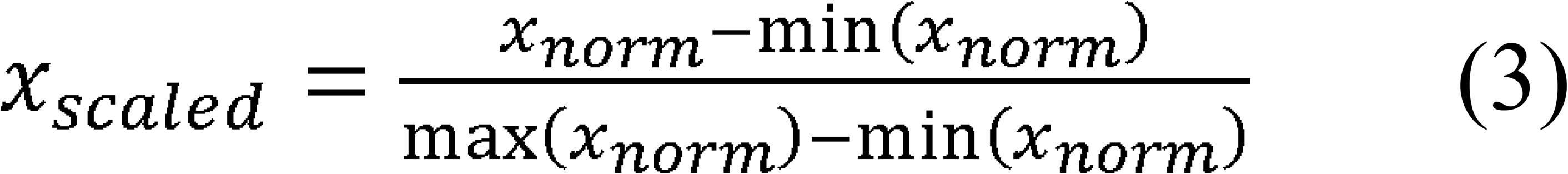

Gene expression values were then normalized across tissue samples using an identical procedure. All available tissue samples were used in the normalization process regardless of whether they were assigned to a brain region. Tissue samples not matched to a brain region were discarded after normalization. Samples assigned to the same brain region were averaged separately for each donor and then across donors, yielding a regional expression matrix with 44 rows, corresponding to brain regions, and 15,633 columns, corresponding to the retained genes.

RNA sequencing data used for the second objective of this study were obtained from the Allen Institute for Brain Science BrainSpan Atlas (http://www.brainspan.org/). Information regarding tissue collection, preparation, RNA sequencing, and processing are reported on the website.

### Statistical analysis

All statistical analysis was computed using R v4.1.1. Software packages are specified at the point of use below.

#### Thalamic nuclei volume differences between young adult *C9orf72* expansion carriers and non-carriers

A general linear model was fitted with bilateral thalamic nucleus volume as the dependent variable, group by carrier status as the predictor of interest, and baseline age, sex, total intracranial volume, and scanner differences as covariates. Per thalamic nuclei model, the t-statistic associated with the group coefficient was extracted and used for spatial transcriptomic correlation analyses,^33,34^ where a negative t-statistic reflected a smaller volume in the carrier group, and vice versa. To ease interpretation of downstream correlations with transcriptomic expression, imaging model t-statistics were multiplied by -1, such that larger values corresponded to larger effects of smaller nucleus volume in the carrier group.

Sensitivity analyses involved examining thalamic nuclei volumes between groups for ALLFTD participants alone, since some of the young adult GENFI participants may have been included in a prior study examining subcortical volumes.^19^

#### Spatial neuroimaging-transcriptomic associations across thalamic nuclei

##### Data-driven approach

For the data-driven approach, 15,633 transcripts were filtered by determining those which yielded the greatest spatial variance across all thalamic nuclei. The top 30%, 25%, and 10% of transcripts of greatest spatial variance yielded 4,690, 2906, and 1,564 transcripts, respectively. The top 10% of transcripts yielding greatest spatial variance were visually verified against mean expression levels to ensure that they did not overrepresent only transcripts which were most highly expressed. These transcripts were then inputted into a non-negative matrix factorization (NMF) model using R NMF v0.28 on the transcript-by-nucleus expression matrix with ten *a priori* latent components and fifty random initializations computed to improve model stability.^35^ The resulting component score matrix was used as predictors in a linear model implemented in Limma v3.62.2 to assess associations between latent expression patterns and transcript expression profiles across thalamic nuclei. From the resulting Limma output, the NMF component showing the strongest overall transcript-wise association signal was selected based on survival of Benjamini-Hochberg correction at α=0.05. The transcripts which comprise the significant NMF component were visualized using a volcano plot.

Transcripts were ranked by their weight contribution to the significant NMF component, and the top 100 transcripts were used in downstream analyses. This included fitting linear models with Limma to examine associations between normative expression patterns across the thalamic nuclei, and computing Spearman-rank correlations with *C9orf72*, with Benjamini-Hochberg corrections applied.

##### Hypothesis-driven approach

The hypothesis-driven approach examined the spatial normative expression profiles of 37 transcripts; these were for their relevance to genetic FTLD,^1^ their relevance to human thalamic neurodevelopment during the first two trimesters of birth,^21^ or their combined relevance to human neurodevelopment and neurodevelopmental disorders^36^ and *C9orf72* neurodevelopmental roles.^14^ The genes were: *GRN, MAPT, TMEM106B, TARDBP, FUS, VCP, TBK1, CHMP2B, SQSTM1, CHCHD10, OPTN, CCNF, TIA1, EWSR1, TAF15, NES, HES1, CALB1, CALB2, PVALB, CNTNAP2, MET, NLGN4X, NRGN, CEACAM21, SLC24A5, RGS16, FOXP2, TCF7L2, LHX2, SLC17A6, S100A6, SOX2, GAD1, SLC32A1, DLX1, DLX2*. (See **Supplementary Table 1** for descriptions per gene.) Next, as in the data-driven approach, Spearman-rank correlations with *C9orf72* were computed with the 37 FTLD- and thalamic neurodevelopment-related transcripts of interest.

#### Exploratory gene ontology (GO) analyses

GO analysis was performed using ClusterProfiler v4.14.6 for the top 100 data-driven transcripts, and then separately for the 70 transcripts that were significantly associated with *C9orf72*, to examine potential gene function relationships with *C9orf72*. For the hypothesis-driven analysis, the transcripts which survived Benjamini-Hochberg correction underwent GO analysis to examine their biological processes, cellular components, and molecular functions in relation to *C9orf72*.

#### Sensitivity analyses

Sensitivity analyses were computed to better delineate sex- and age-related findings without removing the single female AHBA donor brain from the primary analysis. This included examination of each individual donor separately, as well as the two younger donors (aged in their early 20s and 30s) in aggregate, and the remaining older donors (aged between 30-59 years) in aggregate, to verify the expression patterns of the top 100 data-driven transcripts derived from the NMF of the primary analyses, and of the 37 hypothesis-driven transcripts of interest. For sensitivity analyses of the individual donors, when multiple probes indexed the expression of the same gene, we selected and used the probe with the highest mean intensity across samples instead of applying the differential stability calculation meant for selecting a probe across multiple donors. Transcripts which were not captured by the microarray probes were not available to be assessed in the sensitivity analysis.

#### Spatial expression of FTLD- and neurodevelopment-related transcripts across healthy prenatal stages to young adulthood

Expression levels of the *n*=37 hypothesis-driven transcripts of interest were visualized in the developing thalamus and the occipital cortex per the BrainSpan Atlas, which offers 26 regions reflective of the human developing brain. The BrainSpan Atlas defined the thalamus as the “dorsal thalamus” during first trimester of prenatal life, and the “mediodorsal nucleus of the thalamus” thereafter, from second trimester of prenatal life to 40 years old, for which donor samples existed. Similarly, the occipital cortex was defined as the “occipital neocortex” during first trimester of prenatal life and the “primary visual cortex (striate cortex, area V1/17)” thereafter.

Differential spatial expression of these transcripts in normative development was examined with linear models employed with Limma. One model compared the thalamic structures, and another compared the occipital structures. Each model was computed on the gene expression matrix while adjusting for developmental stage and sex. Given the relatively small number of available donor samples, developmental stage was included as a covariate and operationalized as a factor with two levels: “prenatal” and “postnatal”. This approach was chosen in lieu of modelling age as a continuous covariate, as the available donor sample size was insufficient to robustly estimate a non-linear model to capture age-related influences on human neurodevelopment. Spearman-rank correlations with *C9orf72* were computed for normative spatial transcript expression in the thalamic and the occipital structures using the residual transcript expression values after regressing out the effects of developmental stage and sex using the Limma linear model.

## Results

### Participant demographics

Young adult *C9orf72* repeat expansion carriers (*n*=72) and non-carriers (*n*=48) from the FPI did not differ in age (range 18-30 years), sex, and total intracranial volume **(Table 1)**. Summary demographics of healthy donors who contributed to the AHBA and BrainSpan transcriptomic data are also outlined in Table 1. See **Supplementary Table 2** for information on individual AHBA and BrainSpan donors.

**Table 1:**
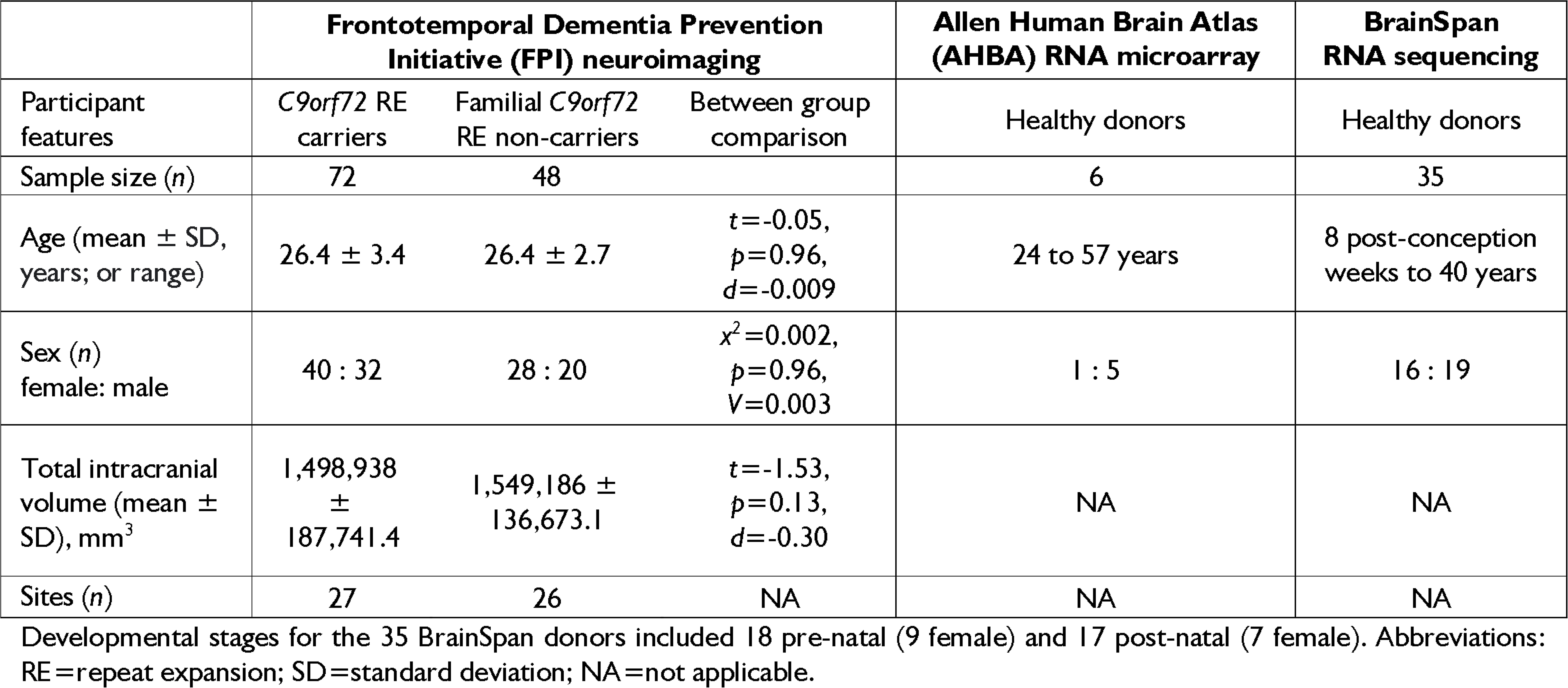
Characteristics of participants included from FPI, AHBA and BrainSpan.

### Group-level thalamic nuclei volume differences

Young adult *C9orf72* repeat expansion carriers demonstrated smaller volumes in primarily the bilateral pulvinar, medial, and posterior thalamic nuclei compared to non-carriers while adjusting for age, sex, scanner, and total intracranial volume **(Supplementary Table 3)**. Carriers had smaller average volumes in the right and left medial pulvinar (PuM): *F*(1,87)=28.8, *p*=6.54e-7, and *F*(1,87)=8.74, *p*=4.00e-3, respectively; right and left anterior pulvinar (PuA): *F*(1,87)=17.3, *p*=7.57e-5, and *F*(1,87)=6.84, *p*=0.011, respectively; right and left inferior pulvinar (PuI): *F*(1,87)=11.8, *p*=8.90e-4, and *F*(1,87)=4.91, *p*=0.029, respectively; right and left mediodorsal medial magnocellular nucleus (MDm): *F*(1,87)=8.74, *p*=4.02e-3, and *F*(1,87)=4.09, *p*=0.046, respectively; right and left mediodorsal lateral parvocellular nucleus (MDl): *F*(1,87)=7.22, *p*=8.62e-3 and *F*(1,87)=4.72, *p*=0.032, respectively; left lateral posterior (LP) nucleus: *F*(1,87)=5.43, *p*=0.022; and left laterodorsal (LD) nucleus: *F*(1,87)=4.37, *p*=0.040.

### Neuroimaging-transcriptomic associations across thalamic nuclei

#### Data-driven approach

Of the ten NMF-derived transcriptomic components, only Component 7 survived Benjamini-Hochberg correction, exhibiting significant spatial differential expression, with higher normative expression of transcripts in thalamic nuclei that are smaller in young adult *C9orf72* expansion carriers: *ß*=0.24, *t*=3.00, *p_adj_*=0.039 (**Figure 2; Supplementary Table 4)**. Analyses focused on the top 100 transcripts which contributed most weight to Component 7 (see **Supplementary Table 5** for transcript weights). Normative expression patterns of these top 100 transcripts were higher in thalamic nuclei which yielded group-level differences, namely the pulvinar, medial, and lateral nuclei **(Figure 3a)**. In contrast, normative expression levels of the top 100 transcripts were lower in the ventral nuclei, and were mixed in the intralaminar and anterior nuclei.

**Figure 2.**
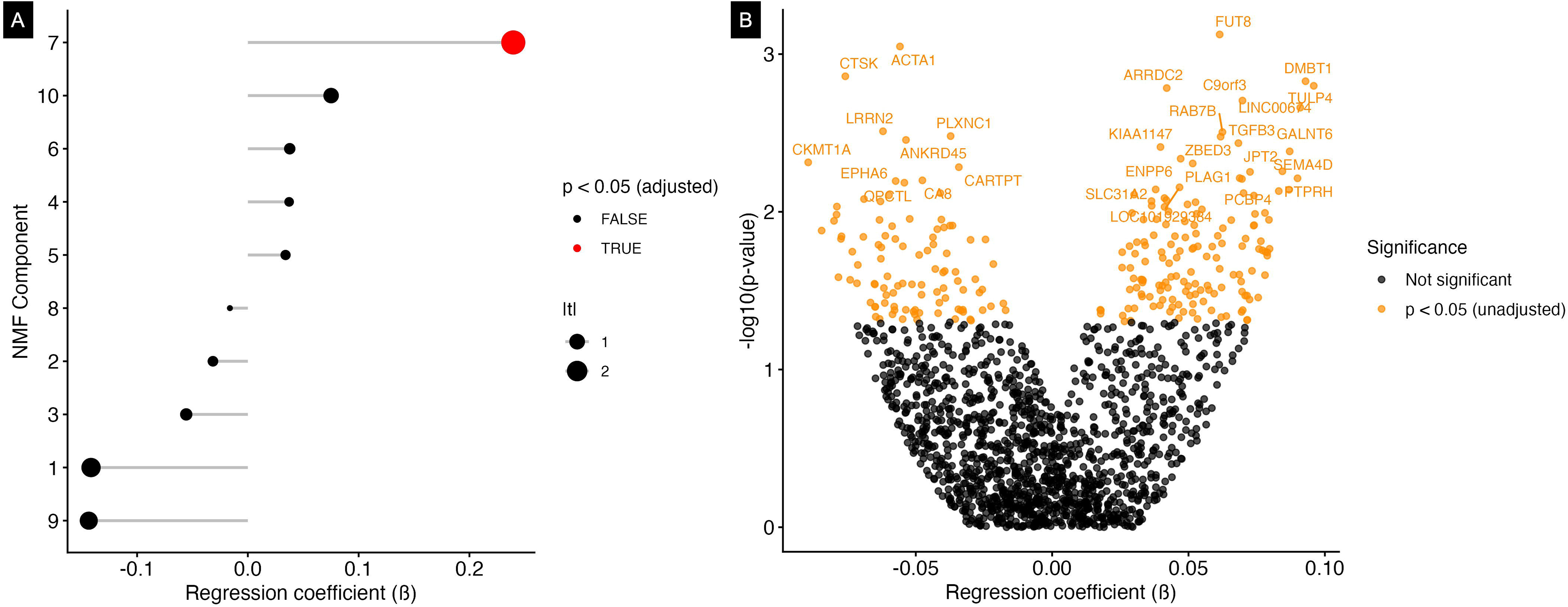
The top 10% of transcripts yielding the greatest spatial variance across all thalamic nuclei. **(A)** A non-negative matrix factorization (NMF) with ten components. **(B)** All transcripts from the NMF in a volcano plot. Component 7 of the NMF survived correction, where 152 transcripts were positively associated with volume differences across thalamic nuclei, and 100 were negatively associated. For display, the top 30 most positively and negatively associated transcripts are labeled. Downstream *a priori* analyses focused on the top 100 transcripts which contributed most weight to Component 7, which encompassed transcripts that showed higher normative expression (indicated by positive regression coefficients) in thalamic nuclei that are smaller in young adult *C9orf72* expansion carriers.

**Figure 3.**
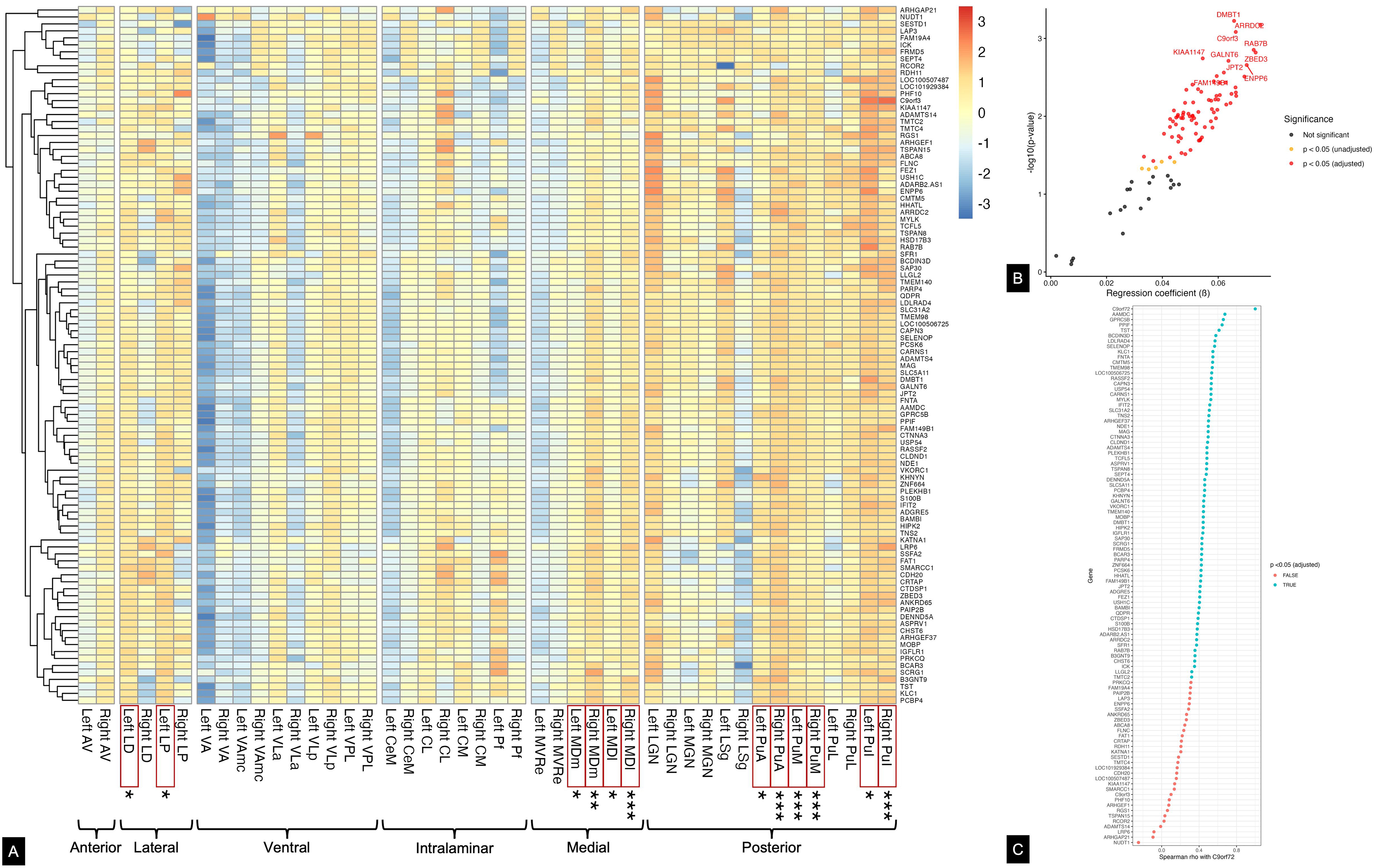
Neuroimaging-transcriptomic associations of the data-driven transcripts. **(A)** Normative spatial expression patterns of the top 100 data-driven transcripts across thalamic nuclei. Thalamic nuclei that are smaller in young adult *C9orf72* repeat expansion carriers compared to non-carriers are boxed in red (\**p*<0.05, \*\**p*<0.01, \*\*\**p*<0.001). **(B)** Higher normative spatial expression patterns of 76 transcripts were observed in thalamic nuclei that are smaller in young adult *C9orf72* repeat expansion carriers; only the top 10 are labeled for display. **(C)** Normative *C9orf72* expression was significantly correlated with normative expression patterns of 70 transcripts across the thalamic nuclei.

A linear model fitted for the *C9orf72* transcript and the top 100 transcripts contributing to Component 7 revealed 76 transcripts that survived correction for multiple comparisons. All 76 transcripts yielded higher normative expression in nuclei that have smaller volumes in *C9orf72* expansion carriers **(Figure 3b; Supplementary Table 6)**. Spearman-rank correlations of the top 100 transcripts with *C9orf72* yielded 70 that were positively associated with normative *C9orf72* expression across all the thalamic nuclei, and which survived correction **(Figure 3c; Supplementary Table 7)**.

Fifty-six transcripts showed significant associations between normative spatial expression patterns and all thalamic nuclei volumes, and significant correlations with normative *C9orf72* transcript expression: *DMBT1, ARRDC2, RAB7B, GALNT6, JPT2, FAM149B1, PCBP4, SLC31A2, SLC5A11, TSPAN8, ADAMTS4, TMEM98, KHNYN, USP54, PCSK6, TMTC2, CARNS1, NDE1, CLDND1, FEZ1, ASPRV1, MAG, HSD17B3, LDLRAD4, LLGL2, TMEM140, IFIT2, USH1C, BAMBI, CTNNA3, KLC1, TCFL5, LOC100506725, ADGRE5, ZNF664, SELENOP, SFR1, VKORC1, PARP4, ARHGEF37, SAP30, MYLK, CTDSP1, BCDIN3D, CAPN3, TNS2, ADARB2.AS1, B3GNT9, GPRC5B, RASSF2, HHATL, FNTA, CMTM5, SCRG1, CHST6, QDPR.* Twenty transcripts showed significant associations between normative spatial expression patterns and thalamic nuclei volumes but were not associated with *C9orf72*, and the normative expression of 13 transcripts correlated significantly with *C9orf72* but did not yield spatially differential patterns (see list in **Supplementary Material**).

#### Hypothesis-driven approach

Normative expression patterns of the 37 FTLD- and neurodevelopment-related transcripts across the thalamic nuclei are shown in **Figure 4a**. Lower normative expression of *CALB1* was significantly associated with thalamic nuclei that are smaller in young adult *C9orf72* expansion carriers, and survived Benjamini-Hochberg correction **(Figure 4b; Supplementary Table 8)**. Higher normative expression patterns of *NES* and *SQSTM1* were associated with thalamic nuclei that are smaller in young adult *C9orf72* expansion carriers, but did not survive correction. The 21 transcripts which demonstrated significant correlations with *C9orf72* expression were *DLX2, NES, DLX1, SQSTM1, FOXP2, MAPT, FUS, CNTNAP2, TCF7L2, PVALB, RGS16, CHCHD10, CHMP2B, SLC17A6, SOX2, TAF15, CCNF, MET, LHX2, CALB2,* and *S100A6* **(Figure 4c; Supplementary Table 9)**.

**Figure 4.**
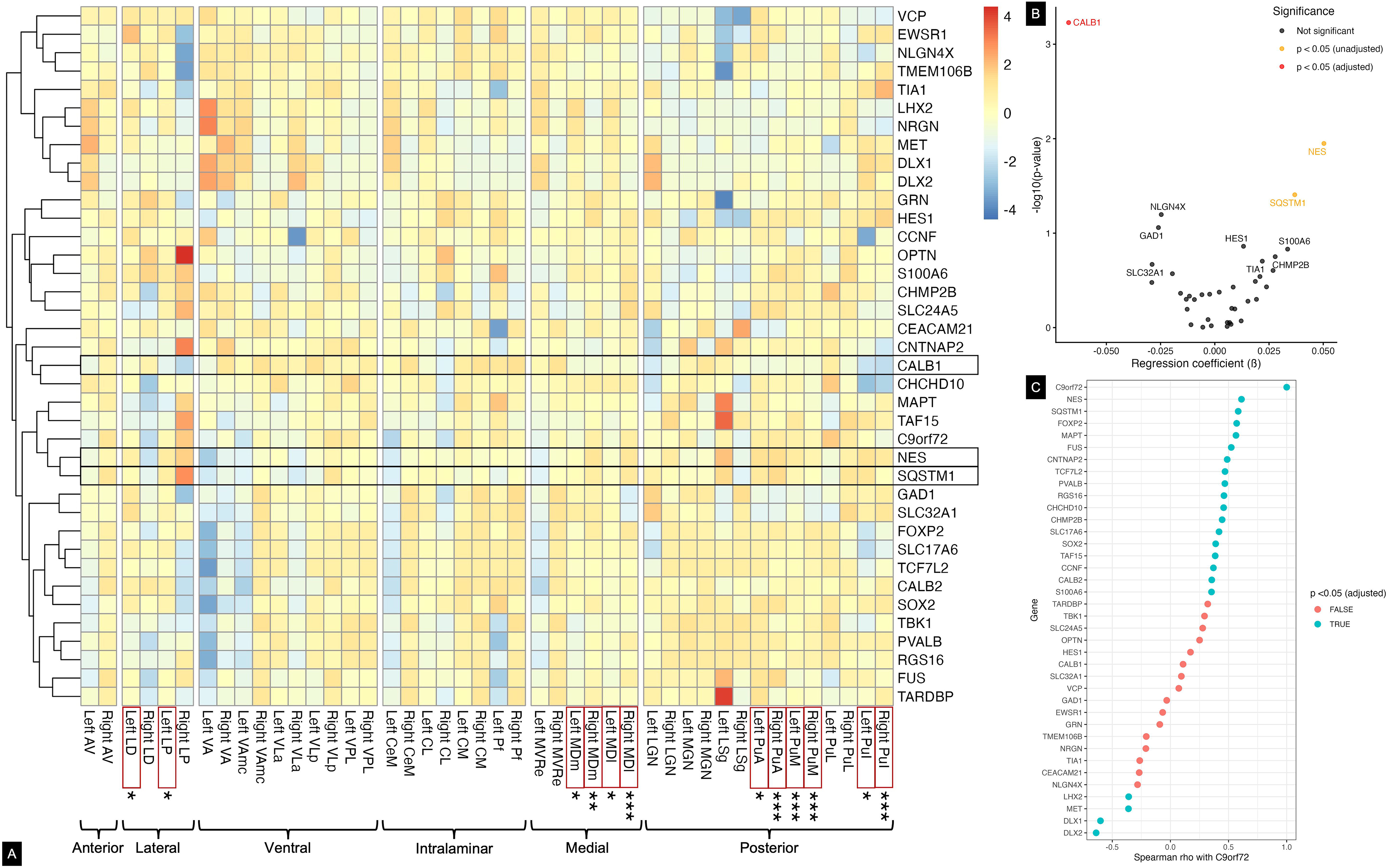
Neuroimaging-transcriptomic associations of the 37 FTLD- and neurodevelopment-related transcripts. **(A)** Normative spatial expression patterns of all hypothesis-driven transcripts across thalamic nuclei. Thalamic nuclei that are smaller in young adult *C9orf72* repeat expansion carriers compared to non-carriers are boxed in red (\**p*<0.05, \*\**p*<0.01, \*\*\**p*<0.001). Rows boxed in black belong to transcripts which showed associations between spatial expression patterns and imaging phenotypes, though only *CALB1* survived corrections for multiple comparisons. **(B)** Lower normative expression patterns of *CALB1* were associated with thalamic nuclei volume differences between groups. **(C)** Normative *C9orf72* expression was significantly correlated with the normative patterns of 21 transcripts across the thalamic nuclei.

#### Exploratory GO analyses

GO analysis of the top 100 data-driven transcripts revealed 144 terms (108 biological processes, 20 cellular components, and 16 molecular functions), though they did not survive Benjamini-Hochberg correction at *p*<0.05 and *q*<0.20 **(Supplementary Figure 1; Supplementary Tables 9-10)**. GO analysis of the 21 hypothesis-driven transcripts and *C9orf72* revealed 84 terms (72 biological processes and 12 cellular components), all of which survived Benjamini-Hochberg correction **(Supplementary Figure 2; Supplementary Table 11)**.

#### Sensitivity analyses

Sensitivity analyses of thalamic nuclei volumes in ALLFTD participants, who were not included in prior subcortical volume analyses,^19^ revealed findings generally consistent with the primary analyses **(Supplementary Table 13)**. Expression patterns of the data-driven and hypothesis-driven transcripts were characterized in each of the six AHBA donors individually, the two younger donors (early 20s and 30s) in aggregate, and the four older donors (aged between 30-59 years) in aggregate. The sensitivity analyses demonstrated consistent spatial normative expression patterns of the top 100 data-driven transcripts and 37 hypothesis-driven transcripts across all donors, though inter-individual variability was also present **(Supplementary Material and Supplementary Tables 14-15)**. Effect magnitudes differed depending on the donor, where the youngest and oldest male donors showed relatively smaller ß, three 30-55-year-old male donors showed relatively similar ß, and the female donor showed relatively larger ß for the top 100 transcripts. Expression patterns of *CALB1* were also consistent in effect direction and magnitude in sensitivity analyses in all donors, and in analyses which aggregated the two younger donors and the four older donors.

### Normative spatial expression in thalamic and occipital structures spanning prenatal stages to young adulthood

Normative expression of the 37 transcripts of interest was visualized in the developing thalamus and occipital structures (as a control region) from BrainSpan donors across 8 weeks post-conception to 40 years old **(Figure 5)**. Differential spatial expression was observed across the developing thalamic and occipital structures **(Figure 6a and b)**. *LHX2* showed higher normative expression during prenatal stages of development in the thalamus (8 to 24 post-conception weeks), with notably highest expression during the earliest stages of life (8 and 9 post-conception weeks) in the dorsal thalamus, whereas *GAD1, CEACAM21, CALB2*, and *SLC32A1* showed higher normative expression during postnatal stages (4 months to 40 years old) in the mediodorsal nucleus of the thalamus **(Figure 6c; Supplementary Table 16a).** In contrast, these transcripts were not significantly differentially expressed during normative occipital cortex development **(Figure 6d; Supplementary Table 16b)**. In the occipital cortex, *HES1, TCF7L2, RGS16, CCNF,* and *NES* expression levels were higher during prenatal development, and *OPTN* and *MAPT* were higher during postnatal stages.

**Figure 5.**
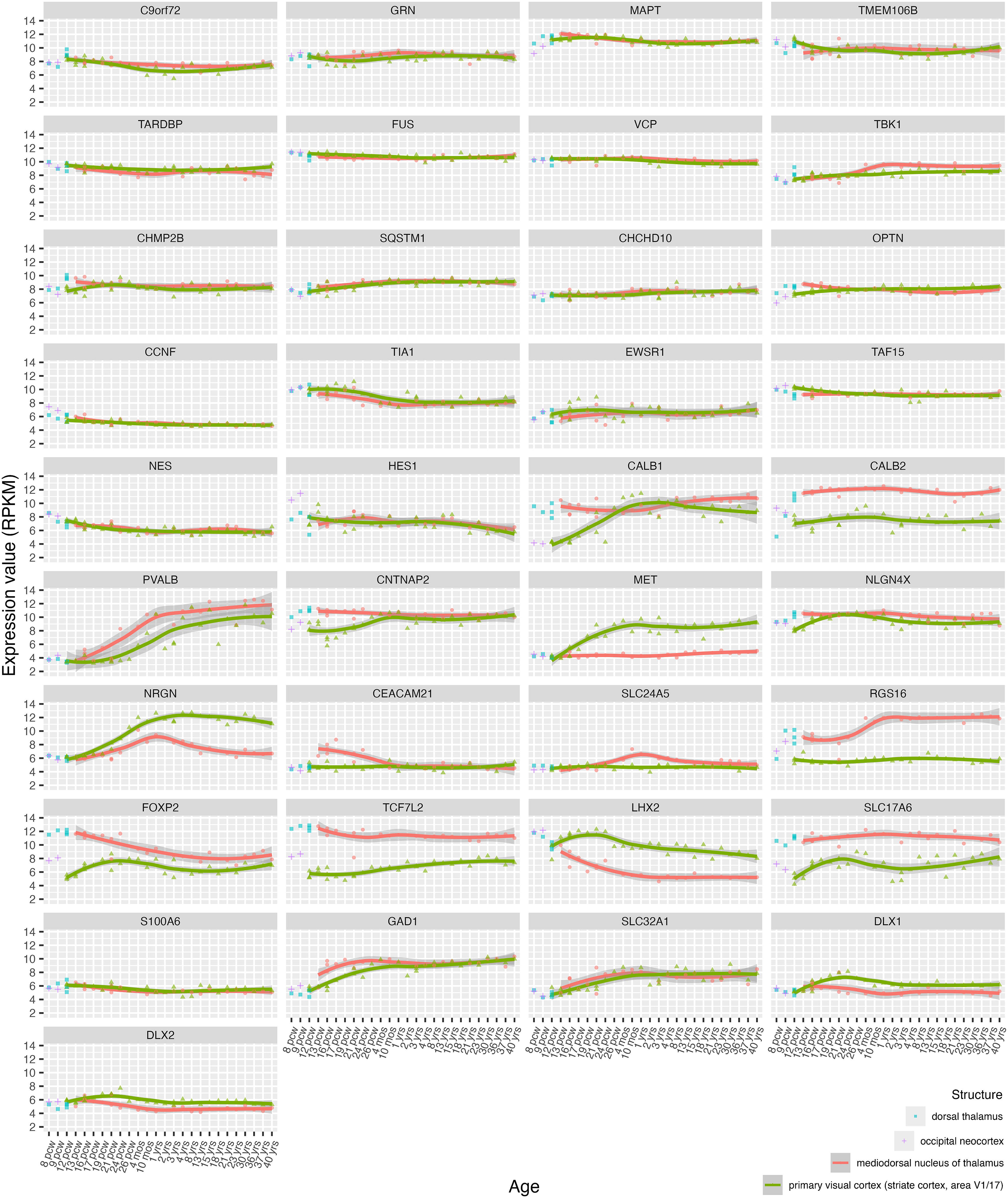
Normative expression of *C9orf72* and 37 FTLD- and neurodevelopment-related transcripts across prenatal to young adulthood ages. Spatial expression profiles were visualized in the thalamic structures (vulnerable to smaller volumes in young adult *C9orf72* expansion carriers) and occipital structures (not known to be affected by atrophy in FTLD). Structures were defined by the BrainSpan atlas of the developing human brain.

**Figure 6.**
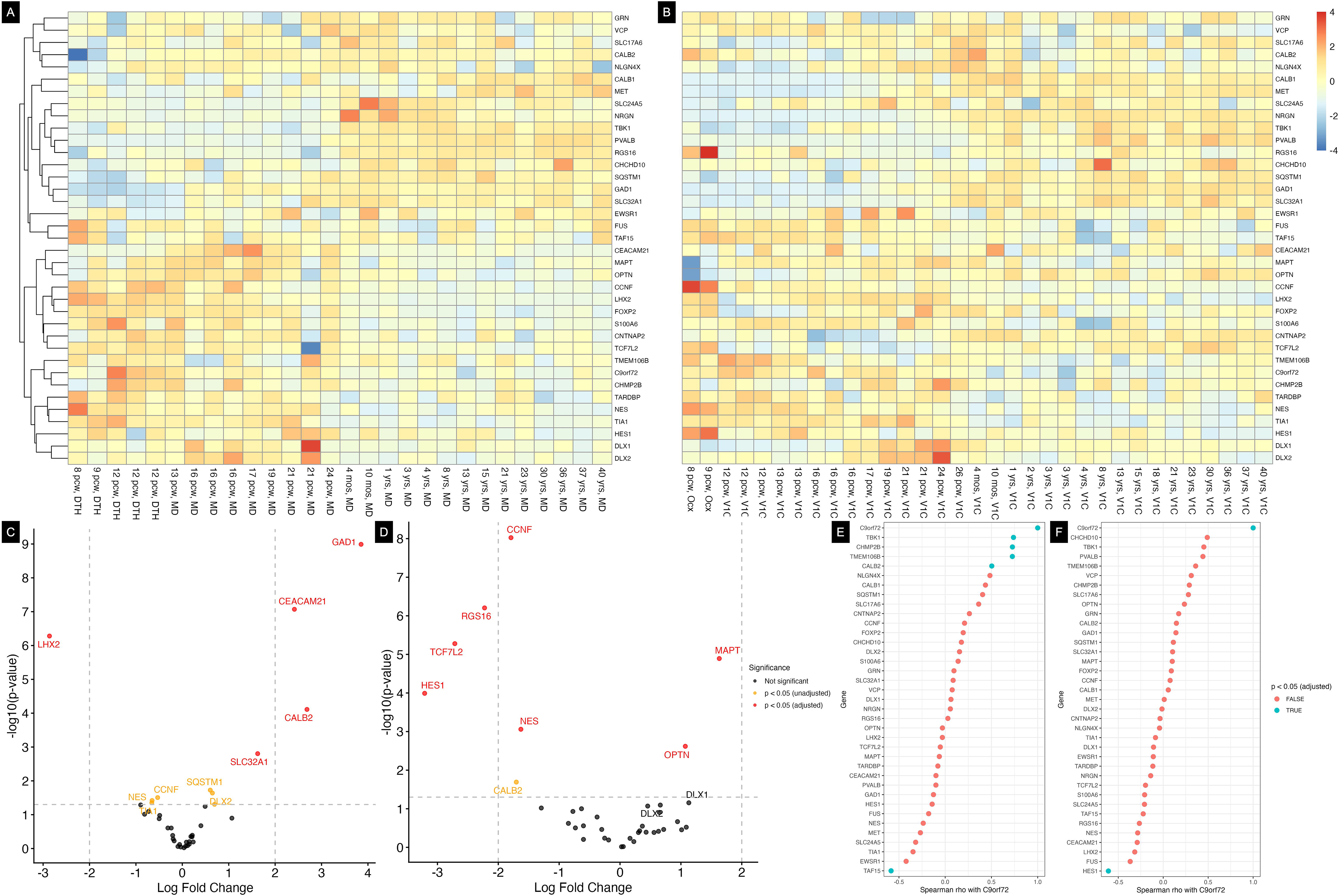
Spatial expression of FTLD- and neurodevelopment-specific transcripts in healthy BrainSpan donors across prenatal to young adulthood ages in. **(A)** thalamic and **(B)** occipital regions. Differential spatial expression of these transcripts was observed across the developing **(C)** thalamic and **(D)** occipital structures. Normative expression patterns associated with *C9orf72* in the **(E)** thalamus compared to **(F)** occipital structures.

Normative expression of different transcripts correlated with *C9orf72* in the thalamus compared to the occipital structures across prenatal stages to young adulthood. In the thalamus, normative expression patterns of *TBK1, CHMP2B, TMEM106B,* and *CALB2* were positively associated with *C9orf72*, and *TAF15* patterns were negatively associated **(Figure 6e; Supplementary Table 17a)**. In the occipital cortex, *HES1* was the sole transcript significantly negatively associated with *C9orf72*; no positive associations with *C9orf72* were observed **(Figure 6f; Supplementary Table 17b)**.

## Discussion

Specific thalamic nuclei, namely the pulvinar, mediodorsal, and lateral nuclei, showed smaller average volumes in asymptomatic young adult *C9orf72* repeat expansion carriers relative to non-carriers. Consistent with our hypotheses, associations of the young adult FTLD cohort neuroimaging data and normative AHBA transcriptomics data yielded spatial differential expression of normative transcripts involved in neurodevelopment and *C9orf72*-associated FTLD neurodegeneration. The hypothesis-driven findings further supported neurodevelopmental associations, where *CALB1*, a prominent marker of human prenatal thalamic development, demonstrated lower normative expression in the pulvinar, mediodorsal, and lateral thalamic nuclei. Normative expression of other prenatal thalamic markers, including *CALB2*, *PVALB*, and *FOXP2*, also correlated with *C9orf72* expression in AHBA donors, with *CALB2* yielding consistent correlations across the developing thalamus in BrainSpan donors.

Extending prior findings from older asymptomatic *C9orf72* repeat expansion carriers with a mean age of 43.9±11.7 years^19^ to the present results in young adulthood, this sample of young adult carriers aged 18-30 years demonstrated smaller volumes in the pulvinar (bilateral PuM, PuA, and PuI), mediodorsal (bilateral MDl and MDm), and lateral (left LD and LP) thalamic nuclei. This pattern of thalamic nuclei volume differences in *C9orf72* expansion carriers vs. non-carriers is consistent with the functional neuroanatomy of the affected thalamic nuclei^20,37–41^ and the associated neuropsychiatric symptoms in FTLD.^42,43^ Associations of the thalamic nuclei volumes in young adult FTLD neuroimaging data with normative transcriptomics from AHBA adults yielded spatial differential expression of transcripts involved concurrently in *C9orf72*-mediated FTLD neurodegeneration and in neurodevelopmental processes.

The functions of the significant transcripts from the data-driven approach consistently highlight relevant dual roles in *C9orf72*-associated FTLD neurodegeneration and neurodevelopment related to endosomal, lysosomal and autophagic processes. Indeed, *C9orf72* encodes the C9orf72 protein, which functions as a Rab guanine nucleotide exchange factor, and is a key regulator of endosomal transport, lysosomal homeostasis, and autophagy.^44^ Lysosomal and autophagy dysfunction is one pathophysiological mechanism of *C9orf72*-associated FTLD.^45^ Simultaneously, the identified data-driven transcripts are also involved in endosomal-lysosomal trafficking and autophagy: *RAB7B, DENND5A, ARRDC2, FEZ1, B3GNT9, SCRG1.* Other significant transcripts, including *RAB7B*, *DENND5A, KIAA1147* (also known as *DENND11*)*, ARHGEF37*, *RASSF2*, and *FNTA*, likewise encode small GTPases or regulator proteins that regulate membrane trafficking and signaling pathways implicated in both development and homeostasis. For example, *RASSF2* encodes a tumour suppressor protein involved in the Ras MAPK/ERK pathway and is a regulator of cell fate and apoptosis,^46^ and *FNTA* encodes subunits for enzymes that modify Ras signaling proteins involved in cell growth and migration, and its protein and co-effectors in the signaling pathway have been co-localized with mutant huntingtin in the striatum of a mouse model of Huntington’s disease.^51^

Aligning with our findings for *RAB7B*, which showed higher normative expression in thalamic nuclei that are smaller in *C9orf72* expansion carriers and was positively associated with normative patterns of *C9orf72*, the Rab7 protein has been co-localized with C9orf72, Rab5, and Rab11 in *C9orf72*-ALS patient motor neurons compared to healthy controls.^52^ One interpretation therefore is that the data-driven transcripts highlight neurodegenerative mechanisms relevant to *C9orf72*-associated FTLD, especially since the normative transcriptomics were derived primarily from adult brains past the last stages of neurodevelopment. Other identified data-driven transcripts provide additional support for this interpretation as they are involved in neurodegenerative disease mechanisms, including *GALNT6* (involved in O-glycosylation of amyloid precursor protein (APP) and related to Aß-40 and Aß-42 production in Alzheimer’s disease^54^), and *KLC1* (binds APP for anterograde transport^55^).

While some findings support potential neurodegenerative related candidate transcripts associated with thalamic nuclei volumes, many of the significant data-driven transcripts are also involved in prenatal neurodevelopmental processes, including prenatal thalamic development. An example is the DENN domain-related transcripts, such as *KIAA1147* (or *DENND11*; which showed spatial associations across thalamic nuclei) and *DENND5A* (which showed associations with normative *C9orf72* expression). Indeed, DENN-C9orf72 protein interactions have been reported as a potential neurodevelopmental mechanism involved in regulating hindbrain formation, axonogenesis, neuronal motility, and apoptosis in a zebrafish embryo model of *C9orf72*-associated FTLD/ALS.^7^ In a human-induced pluripotent stem cell model, the DENND5A protein was found to interact with the Crumbs apical polarity complex and thus crucial for proper neural progenitor cell division and neurogenesis.^56^ Other data-driven transcripts have roles in oligodendroglia progenitor cell differentiation, oligodendrocyte development, and myelinogenesis (*MAG, MOBP, FEZ1*)^57,58^; cortical neuron projection and polarity (*NDE1*)^59^; and embryonic left-right axis determination^60^ and brain lateralization and handedness in dyslexia (*PCSK6*).^61^ Transcripts such as *ZBED3,*^62^ *TMEM98,*^63^ and *LRP6*^21^ are also important for embryonic neural patterning and cell fate determination through Wnt/ß-catenin signaling, which is critical for prenatal thalamic patterning.^64^ Indeed, expression of *LRP6* (which encodes a Wnt co-receptor) in human radial glia cells has been observed during first trimester of thalamic neurogenesis.^21^ *TMEM98* was significantly differentially expressed across thalamic nuclei and associated with normative *C9orf72* expression, though *LRP6* and *ZBED3* were only associated with spatial differential expression across the thalamic nuclei. Intriguingly, *TMEM98,*^65^ alongside *HHATL*65 and *FAM149B1,*^66^ are transcripts involved in sonic hedgehog signaling, another pathway crucial for patterning, anterior-posterior axis determination, differentiation of the distinct thalamic nuclei,^64^ and for migration of GABAergic neurons in human thalamic tissues between 8 to 15 post-conception weeks.^67^ Loss of function variants in many of the identified data-driven transcripts have been associated with neurodevelopmental disorders, including *DENND5A* with developmental and epileptic encephalopathies,^56^ *NDE1* with microlissenencephaly,^68^ *FAM149B1* with Joubert syndrome and orofaciodigital syndrome VI,^69^ *FEZ1* with schizophrenia,^53^ *CTNNA3* with autism spectrum disorder (ASD).^65^

Roles of these transcripts may not be purely neurodevelopmental and may also be crucial for maintaining mature neuronal connectivity during adulthood, especially when considering the transcripts involved in neuroimmune and glial signaling, including *DMBT1*, *ARRDC*2, *ASPRV1, LDLRAD4, LLGL2*, *TMEM140, ADGRE5*, and *GPRC5B*. However, normative human cortical brain transcriptomics relevant to adolescent plasticity notably involves transcripts that are crucial for synaptic plasticity, learning, memory, and the neuroimmune system, and these transcriptomic patterns have been associated with atypical neuroimaging, brain tissue RNA-seq, and GWAS findings in schizophrenia.^66^ Similarly, differential normative transcriptomic patterns relevant to neuronal hierarchy, including GABAergic genes important in human prenatal thalamic neurodevelopment such as *CALB1*, *CALB2*, and *PVALB*, have been correlated with neuroimaging phenotypes of ASD.^66^

The hypothesis-driven investigation into associations between young adult *C9orf72*-associated FTLD neuroimaging and normative transcriptomics yielded consistent findings. Lower normative expression of *CALB1* was significantly associated with the nuclei that had smaller volumes in *C9orf72* expansion carriers, with sensitivity analyses separately computed in the two younger donors and four older donors both supporting these findings. *CALB1* (which encodes calbindin-D28k), *CALB2* (encodes calretinin), and *PVALB* (encodes parvalbumin) are prominent markers of prenatal human thalamic development.^21,62^ In humans, CALB1 protein expression has been observed at 10 weeks post-conception in more lateral thalamic regions, and when the nuclei become distinct at 14 weeks post-conception, in the LD nucleus and the LGN.^62^ Furthermore, in primates, thalamic neurons that are immunoreactive for CALB1 and CALB2 primarily project to the superficial cortical layers of the prefrontal cortex,^67,68^ and CALB2-positive thalamocortical neurons also project to the ventral striatum and limbic regions.^69^

Normative expression of *CALB1* was not significantly correlated with *C9orf72* in this study, but positive correlations of *CALB2*, *PVALB*, and *FOXP2* (another prominent marker of human thalamic development whose protein coordinates thalamic patterning^70^), alongside other hypothesis-driven transcripts, were observed with *C9orf72*. That *CALB2*, *PVALB*, and *FOXP2* did not show significant neuroimaging-transcriptomic correlations is perhaps unsurprising, because during human prenatal development, *CALB2* is normally expressed in the rostral intralaminar nuclei,^21^ *PVALB* in the reticular nucleus,^21^ and *FOXP2* throughout both medial and lateral aspects of the thalamus.^62^ Although RNA expression levels do not always correlate to protein levels, and this study was unable to investigate protein levels of the hypothesis-driven transcripts, these collective transcriptomic findings are consistent with literature that supports the spatial differential expression of their respective proteins. When expression levels of hypothesis-driven transcripts in prenatal development were characterized with donors in BrainSpan, *CALB2* showed highest normative expression during 4 months to 40 years old in the mediodorsal thalamus. *CALB2* was also significantly positively correlated with *C9orf72* expression in the developing thalamus of BrainSpan donors, extending the correlations observed in the AHBA adults.

Collectively, the data- and hypothesis-driven neuroimaging-transcriptomic associations yielded transcripts concurrently involved in neurodevelopment and *C9orf72*-associated FTLD neurodegeneration. As the normative transcriptomic patterns are specific to thalamic nuclei that were smaller in young adult *C9orf72* repeat expansion carriers, it is possible that these patterns reflect molecular architectures that define intrinsically vulnerable thalamic nuclei. That many of the identified data-driven transcripts are involved in housekeeping lysosomal and autophagic functions raises the question of whether the affected thalamic nuclei are more vulnerable to cellular stress across the lifespan. It is also intriguing that some identified transcripts are associated with *C9orf72* expression levels across developmental stages and adult life. A next crucial question is whether this pattern of downstream proteins during both early neurodevelopmental and later homeostatic and maintenance stages is also present under *C9orf72* haploinsufficiency, including in preclinical models, and whether these reflect C9orf72 loss-of-function, gain-of-function effects associated with dipeptide repeat proteins, or a combination of mechanisms.

This study had several limitations. No transcriptomic data was available from prenatal human FTLD nor neuroimaging data from youth FTLD. In the absence of their availability, normative transcriptomics enables indirect associations, and the use of young adult FTLD neuroimaging restricts the study scope to neuroimaging differences observed in young asymptomatic adults at the tail end ages of neurodevelopment. Additionally, the observed transcriptomic associations cannot be assumed to reflect protein expression.

In conclusion, pulvinar, mediodorsal, and lateral thalamic nuclei were smaller in young adult *C9orf72* repeat expansion carriers relative to non-carriers. These nuclei were associated with higher normative expression of transcripts concurrently involved in neurodevelopment and *C9orf72*-associated FTLD neurodegeneration, as well as lower normative expression of *CALB1*, a marker of human prenatal thalamic development. Normative expression levels of neurodevelopment-relevant transcripts were positively correlated with *C9orf72* expression, including *CALB2*, where correlations were consistent across prenatal development to adulthood. These findings provide a list of candidate transcripts that may illuminate the pathophysiology leading to smaller thalamic volumes in the presence of a *C9orf72* repeat expansion. Future study of neurodevelopment using preclinical models of *C9orf72*-associated FTLD, with a focus on relevant thalamic neurodevelopmental transcripts identified from this human data, will also be crucial for identifying mechanisms that underlie the relative vulnerability of specific thalamic nuclei.

## Abbreviations

FTLD: frontotemporal lobar degeneration;
AHBA: Allen Human Brain Atlas;
NMF: non-matrix factorization;
PuM: medial pulvinar;
PuA: anterior pulvinar;
PuI: inferior pulvinar;
MDm: mediodorsal medial magnocellular;
MDl: mediodorsal lateral parvocellular;
LP: lateral posterior;
LD: laterodorsal

## Data availability

Requests for neuroimaging data can be made by an FPI-affiliated investigator to the coordinating centre. Processing and analytic scripts can be shared upon reasonable request.

## Acknowledgements

The authors thank all FPI cohort participants and caregivers for their efforts and contributions to this study. I.So and E.C.F. additionally thank Dr. Robyn S. Klein for insightful discussions regarding human brain transcriptomics, and Dr. Emma G. Duerden and Dr. J. Bruce Morton for insightful discussions regarding human neurodevelopment.

## Supplementary material

Supplementary material is available at *Brain* online.

