## Supplementary Material for "Normative transcriptomic clues to thalamic developmental vulnerability in *C9orf7*2-FTLD"

#### GENFI Investigators

Amelia Blesius,[1] David Cash,[1] Clementine Cheyne,[1] Sophie Goldsmith,[1] Luna Nordenstrom,[1] Victoria Pauwels Romero,[1] Hannah Rosenheim,[1] David L. Thomas,[2] Maura Malpetti,[3] Alexander Murley,[3] Sean Tan,[3] Sonia Bellini,[4] Valentina Cantoni,[5] Roberto Gasparotti,[6] Roberta Ghidoni,[4] Enrico Premi,[7] Andrea Arighi,[8] Vittoria Borracci,[8] Chiara Fenoglio,[9] Manuela Pintus,[8] Emanuela Rotondo,[8] Luca Sacchi,[8] Maria Serpente,[8] Vittoria Aprea,[10] Paola Caroppo,[10] Marcella Catania,[10] Giorgio Gelosa,[10] Gemma Lombardi,[10] Pietro Tiraboschi,[10] Sandra Black,[11] Morris Freedman,[12] Sara Mitchell,[11] Ekaterina Rogaeva,[13] David Tang-Wai,[14] Olaiya Aro,[15] Liset de Boer,[15] Julie De Houwer,[15] Laura Donker Kaat,[15] Elise Dopper,[15] Jackie Poos,[15] Babette Reichard,[15] Tine Swartenbroekx,[15] Valentina Bessi,[16] Stefano Chiti,[17] Giulia Giacomucci,[16] Andrea Ginestroni,[18] Valentina Moschini,[18] Sonia Padiglioni,[18] Andrew Douglas,[19] Kevin Talbot,[19] Martin Turner,[19] Vesna Jelic,[20] Elena Rodriguez-Vieitez,[20] Melissa Taheri Rydell,[20] Henrik Viklund,[21] Caroline Dallaire-Théroux,[22] Anna Antonell,[23] Mircea Balasa,[23] Sergi Borrego-Ecija,[23] Paula Canasto,[23] Jordi Junca,[23] Albert Lladó,[23] Tiago Costa-Coelho,[24] Carolina Maruta,[24] Gabriel Miltenberger,[24] Ana Verdelho,[24] Patricia Alves,[25] Myriam Barandiaran,[26] Ioana Croitoru,[25] Alazne Gabilondo,[26] Iliana Quiñones,[25] Mikel Tainta,[26] Lukas Beichert,[27] Benjamin Bender,[28] Lisa Graf,[27] Mathieu Vandenbulcke,[29] Koen Poesen,[29] Sien Van Daele,[29] Aline Delva,[29] Maxime Montembeault,[30] Guy Rouleau,[31] Pierre-François Pradat,[32] Giorgia Querin,[33] Daisy Rinaldi,[33] Anna Stockbauer,[34] Alexander Bernhardt,[34] Ellen Merli,[34] Elisabeth Wlasich,[34] Georg Nübling,[34] Julia Kustermann,[34] Olivia Wagemann,[34] Sonja Schönecker,[34] Lena Marth,[34] Sarah Anderl-Straub,[35] Ilse Dewachter,[36] Stéphanie De Keulenaer,[37] Rosa Rademakers,[37] Marijne Vandebergh,[37] Thijs Vande Vyvere,[37] Sara Van Mossevelde,[37] Xavier Delbeuck,[38] Agnès Denève,[38] Simon Lecerf,[38] Juhana Hakumäki,[39] Sanna Hannonen,[39] Juho Kalapudas,[39] Annemari Kilpeläinen,[39] Ave Kivisild,[40] Mervi Könönen,[39] Inês Baldeiras,[41] Miguel Castelo-Branco,[41] João Durães,[41] Marisa Lima,[41] Maria Rosario Almeida,[41] Miguel Tábuas-Pereira,[41] Annerose Engel,[42] Maryna Polyakova,[42] Frank Regenbrecht,[42] Angelika Thöne-Otto,[42] Anna-Leena Heikkinen,[43] Vesa Kiviniemi,[43] Vesa Korhonen,[43] Noora-Maria Suhonen,[43] Laura Göschel,[44] Jonah Nietiet,[44] Anna Söllner,[44] Teresa Dell'Abbate,[45] Daniele Urso,[45] Daniel Alcolea,[46] Sara Bernal,[46] Oriol Dols,[46] Juan Fortea,[46] Jesus García-Castro,[46] Alberto Lleó,[46] Sara Rubio-Guerra,[46] Judit Selma-González,[46]

1. Department of Neurodegenerative Disease, Dementia Research Centre, UCL Queen Square Institute of Neurology, London, UK
2. Neuroimaging Analysis Centre, Department of Brain Repair and Rehabilitation, UCL Institute of Neurology, London, UK
3. Department of Clinical Neurosciences, University of Cambridge, Cambridge, UK
4. Molecular Markers Laboratory, IRCCS Istituto Centro San Giovanni di Dio Fatebenefratelli, Brescia, Italy
5. Department of Clinical and Experimental Sciences, University of Brescia, Brescia, Italy
6. Neuroradiology Unit, University of Brescia, Brescia, Italy
7. Stroke Unit, ASST Brescia Hospital, Brescia, Italy
8. Neurodegenerative Diseases Unit, Fondazione IRCCS Ca’ Granda Ospedale Maggiore Policlinico, Milan, Italy
9. Department of Biomedical, Surgical and Dental Sciences, University of Milan; Fondazione IRCCS Ca’ Granda Ospedale Maggiore Policlinico, Milan, Italy

10. Fondazione IRCCS Istituto Neurologico Carlo Besta, Milan, Italy
11. Sunnybrook Health Sciences Centre; Sunnybrook Research Institute; University of Toronto, Toronto, Canada
12. Baycrest Health Sciences, Rotman Research Institute, Toronto, Canada
13. Tanz Centre for Research in Neurodegenerative Diseases, University of Toronto, Toronto, Canada
14. Krembil Research Institute, University Health Network, Toronto, Canada
15. Department of Neurology, Erasmus Medical Centre, Rotterdam, Netherlands
16. Department of Neuroscience, Psychology, Drug Research and Child Health, University of Florence, Florence, Italy
17. Neuroradiology Unit, Department of Experimental and Clinical Biomedical Sciences, University of Florence, Florence, Italy
18. SOD Neurologia, Dipartimento di Neuroscienze degli Organi di Senso, AOU Careggi, Florence, Italy
19. Nuffield Department of Clinical Neurosciences, University of Oxford, Oxford, UK
20. Department of Neurobiology, Care Sciences and Society; Division of Clinical Geriatrics / Neurogeriatrics, Karolinska Institutet, Stockholm/Solna, Sweden
21. Theme Inflammation and Aging, Karolinska University Hospital, Sweden
22. Clinique Interdisciplinaire de Mémoire, CHU de Québec; CRCHU de Québec, Université Laval, Québec, Canada
23. Alzheimer's and Other Cognitive Disorders Unit, Hospital Clínic de Barcelona; FRCB-IDIBAPS; University of Barcelona; CIBERNED, Barcelona, Spain
24. Faculty of Medicine; Laboratory of Language Research, Centro de Estudos Egas Moniz, University of Lisbon, Lisbon, Portugal
25. Biogipuzkoa Health Research Institute, Neurosciences Area, Group of Neurodegenerative Diseases, San Sebastian, Spain
26. Cognitive Disorders Unit, Department of Neurology, Donostia University Hospital, San Sebastian, Spain
27. Division of Translational Genomics of Neurodegenerative Diseases, University of Tübingen, Germany
28. Department of Diagnostic and Interventional Neuroradiology, University of Tübingen, Germany
29. KU Leuven; University Hospitals Leuven; Department of Human Genetics; VIB Center for Molecular Neurology, Belgium
30. Douglas Research Centre, Department of Psychiatry, McGill University, Montreal, Canada
31. Montreal Neurological Institute, Department of Neurology & Neurosurgery, McGill University, Montreal, Canada
32. Reference Center for ALS, Département de Neurologie, AP-HP Hôpital Pitié-Salpêtrière, Paris, France
33. Paris Brain Institute (ICM), Sorbonne Université, Inserm U1127, CNRS UMR 7225, Paris, France
34. Neurologische Klinik, Ludwig-Maximilians-Universität München, Munich, Germany
35. Department of Neurology, University of Ulm, Ulm, Germany
36. Department of Neurosciences, Biomedical Research Institute BIOMED, Hasselt University, Belgium
37. VIB Center for Molecular Neurology; University of Antwerp; University Hospital of Antwerp (UZA), Belgium
38. Univ. Lille; Inserm; CHU Lille; Lille Neuroscience & Cognition; LabEx DISTALZ, Lille, France
39. University of Eastern Finland; Kuopio University Hospital, Kuopio, Finland
40. Kuopio University Hospital, Kuopio, Finland
41. University of Coimbra (multiple departments including Neurochemistry Laboratory; ICNAS; Neurology Department; Centre of Neurosciences and Cell Biology), Coimbra, Portugal
42. University of Leipzig; Max Planck Institute for Human Cognitive and Brain Sciences, Leipzig, Germany
43. University of Oulu; Oulu University Hospital; Research Unit of Clinical Medicine / Neurology, Finland
44. Department of Neurology, Charité Universitätsmedizin Berlin, Berlin, Germany
45. Center for Neurodegenerative Diseases and Aging Brain, University of Bari; Pia Fondazione Panico, Italy

46. Department of Neurology, Hospital de la Santa Creu i Sant Pau; FRCB-IDIBAPS; University of Barcelona; CIBERNED, Barcelona, Spain

#### ALLFTD Investigators

Muhammad Abdullah[1]; Liana Apostolova[2]; Brian Appleby[3]; Sami Barmada[4]; Ece Bayram[5]; Bradley Boeve[6]; Hugo Botha[6]; Adam L. Boxer[7]; Andrea Bozoki[8,9,10]; Danielle Brushaber[11]; David Clark[2]; Ciaran M. Considine[12]; R. Ryan Darby[12]; Gregory S. Day[13]; Bradford Dickerson[3]; Dennis Dickson[13]; Kimiko Domoto-Reilly[14]; Emily Dwosh[15]; Kelley Faber[16]; Anne Fagan[17]; Julie A. Fields[18]; Jamie C. Fong[19]; Tatiana Foroud[20]; Leah K. Forsberg[6]; Douglas R. Galasko[21]; Ralitza Gavrilova[6,22]; Tania Gendron[13]; Daniel Geschwind[23]; Nupur Ghoshal[24]; Jill Goldman[25]; Jonathan Graff-Radford[6]; Neill Graff-Radford[13]; Ian M. Grant[26]; Murray Grossman[27]; Chadwick M. Hales[28]; Matthew Hall[7]; Hilary W. Heuer[7]; Lawrence S. Honig[29,30,31]; Ging-Yuek (Robin) Hsiung[15]; Eric Huang[7,32]; Edward D. Huey[29,30,31]; David Irwin[27]; Noah Johnson[11]; David T. Jones[6]; Kejal Kantarci[6]; David Knopman[6]; Tyler Kolander[6]; John Kornak[33]; Walter Kremers[11]; Justin Kwan[34]; Argentina Lario Lago[7]; Maria Lapid[18]; Shannon B. Lavigne[28]; Suzee Lee[7]; Gabriel C. Léger[21]; Irene Litvan[21]; Peter Ljubenkov[7]; Diane Lucente[35]; Ian R. Mackenzie[36]; Joseph C. Masdeu[37]; Lauren Massimo[27]; Scott McGinnis[35]; Corey T. McMillan[27]; Mario F. Mendez[38]; Carly Mester[11]; Toji Miyagawa[28]; Joie Molden[39]; Chiadi Onyike[40]; Alexander Pantelyat[41]; Belen Pascual[37]; Henry Paulson[4]; Leonard Petrucelli[13]; Peter Pressman[39]; Rosa Rademakers[42,43,44]; Vijay Ramanan[6]; Eliana Marisa Ramos[38]; Katherine P. Rankin[7]; Meghana Rao[6]; Katya Rascovsky[27]; Kristoffer W. Rhoads[14]; Aaron Ritter[45]; Erik D. Roberson[46]; Emily Rogalski[26]; Julio C. Rojas[7]; Howard J. Rosen[7]; Rodolfo Savica[6]; William Seeley[7]; Allison Snyder[34]; Adam M. Staffaroni[7]; Anna Campbell Sullivan[1]; Jeremy Syrjanen[11]; M. Carmela Tartaglia[47]; Jack Taylor[7]; Philip W. Tipton[13]; Marijne Vandebergh[42,43,44]; Sandra Weintraub[26]; Dylan Wint[45]; Bonnie Wong[35]; Jennifer Yokoyama[7]

1. Glenn Biggs Institute for Alzheimer's and Neurodegenerative Diseases, UT Health San Antonio
2. Department of Neurology, Indiana University, Indianapolis, IN, USA
3. Department of Neurology, Case Western Reserve University, Cleveland, OH, USA
4. Department of Neurology, University of Michigan, Ann Arbor, MI, USA
5. Parkinson and Other Movement Disorders Center, Department of Neurosciences, University of California San Diego, La Jolla, CA, USA
6. Department of Neurology, Mayo Clinic, Rochester, MN, USA
7. Department of Neurology, Memory and Aging Center, Weill Institute for Neurosciences, University of California, San Francisco, CA, USA
8. Department of Radiology, Michigan State University, East Lansing, MI, USA
9. Department of Neurology, Michigan State University, East Lansing, MI, USA
10. Department of Neurology, University of North Carolina, Chapel Hill, NC, USA
11. Department of Quantitative Health Sciences, Mayo Clinic, Rochester, MN, USA
12. Department of Neurology, Vanderbilt University, Nashville, TN, USA
13. Department of Neuroscience, Mayo Clinic, Jacksonville, FL, USA
14. Department of Neurology, University of Washington, Seattle, WA, USA
15. Division of Neurology, University of British Columbia, Vancouver, BC, Canada
16. Indiana University School of Medicine, National Centralized Repository for Alzheimer's, Indianapolis, IN, USA
17. Department of Neurology, Washington University School of Medicine, St. Louis, MO, USA
18. Department of Psychiatry and Psychology, Mayo Clinic, Rochester, MN, USA
19. UCSF Health Center for Clinical Genetics and Genomics, University of California, San Francisco, CA, USA
20. Department of Medical and Molecular Genetics, Indiana University School of Medicine, Indianapolis, IN, USA

21. Department of Neurosciences, University of California San Diego, La Jolla, CA, USA
22. Department of Clinical Genomics, Mayo Clinic, Rochester, MN, USA
23. Institute for Precision Health; Departments of Neurology, Psychiatry and Human Genetics, UCLA, Los Angeles, CA, USA
24. Departments of Neurology and Psychiatry, Washington University School of Medicine, St. Louis, MO, USA
25. Taub Institute for Research on Alzheimer’s Disease and the Aging Brain, Department of Neurology, Columbia University Irving Medical Center, New York, NY, USA
26. Department of Psychiatry and Behavioral Sciences, Northwestern Feinberg School of Medicine, Chicago, IL, USA
27. Department of Neurology and Penn Frontotemporal Degeneration Center, Perelman School of Medicine, University of Pennsylvania, Philadelphia, PA, USA
28. Department of Neurology, Emory University School of Medicine, Atlanta, GA, USA
29. Taub Institute for Research on Alzheimer’s Disease and the Aging Brain, Columbia University, New York, NY, USA
30. Department of Neurology, Columbia University, New York, NY, USA
31. Department of Psychiatry, Columbia University, New York, NY, USA
32. Department of Pathology, University of California San Francisco, San Francisco, CA, USA
33. Department of Epidemiology and Biostatistics, University of California, San Francisco, CA, USA
34. National Institute of Neurological Disorders and Stroke, USA
35. Department of Neurology, Massachusetts General Hospital and Harvard Medical School, Boston, MA, USA
36. Department of Pathology, University of British Columbia, Vancouver, BC, Canada
37. Department of Neurology, Houston Methodist, Houston, TX, USA
38. Department of Neurology, University of California, Los Angeles, CA, USA
39. Department of Neurology, University of Colorado, Aurora, CO, USA
40. Department of Psychiatry and Behavioral Sciences, Johns Hopkins University, Baltimore, MD, USA
41. Department of Neurology, Johns Hopkins University School of Medicine, Baltimore, MD, USA
42. VIB Center for Molecular Neurology, University of Antwerp, Antwerp, Belgium
43. Department of Biomedical Sciences, University of Antwerp, Antwerp, Belgium
44. Department of Neuroscience, Mayo Clinic, Jacksonville, FL, USA
45. Cleveland Clinic Lou Ruvo Center for Brain Health, Las Vegas, NV, USA
46. Department of Neurology, University of Alabama at Birmingham, Birmingham, AL, USA
47. Tanz Centre for Research in Neurodegenerative Diseases, Division of Neurology, University of Toronto, Toronto, ON, Canada

#### Supplementary Table 1: Descriptions of hypothesis-driven transcripts.

| Transcripts | Reason for inclusion |
|---|---|
| *GRN, MAPT, TMEM106B, TARDBP, FUS, VCP, TBK1, CHMP2B, SQSTM1, CHCHD10, OPTN, CCNF, TIA1, EWSR1, TAF15* | relevance to genetic FTLD (Greaves & Rohrer, 2019, *Lancet Neurol*) |
| *CALB1, CALB2, PVALB, CEACAM21, SLC24A5, RGS16, FOXP2, TCF7L2, LHX2, SLC17A6, S100A6, SOX2, GAD1, SLC32A1, DLX1, DLX2* | relevance to relevance to human thalamic neurodevelopment during the first two trimesters of birth (Kim et al, 2023, *Science*) |
| *NES, HES1, CALB1, CALB2, PVALB, CNTNAP2, MET, NLGN4X, NRGN, MAPT, CEACAM21* | combined relevance to human neurodevelopment and neurodevelopmental disorders (Kang et al, 2011, *Nature*) and *C9orf72* neurodevelopmental roles (So et al, 2026, *Alz Dement*) |

#### Supplementary Table 2a: Demographics of 6 adult donors who contributed tissue to the Allen Human Brain Atlas, from whom normative RNA microarray data was analyzed.

| Anonymized Donor ID | Age range | Sex | Hemisphere available |
|---|---|---|---|
| H0351.2001 / 9861 | 20-25 | M | Bilateral |
| H0351.2002 / 10021 | 35-40 | M | Bilateral |
| H0351.1009 / 12876 | 55-60 | M | Left |
| H0351.1012 / 14380 | 30-34 | M | Left |
| H0351.1015 / 15496 | 45-50 | F | Left |
| H0351.1016 / 15697 | 50-55 | M | Left |

*Nb: exact ages can be found on the AHBA website, but cannot be revealed in the medRxiv version of the manuscript.*

#### Supplementary Table 2b: Demographics information of 35 donors who contributed tissue to BrainSpan, from whom normative RNA sequencing data was analyzed.

| Donor ID | Donor Name | Age range | Sex |
|---|---|---|---|
| 13058 | H376.IIA.51 | 8-10 pcw | M |
| 12833 | H376.IIA.50 | 8-10 pcw | M |
| 12835 | H376.IIB.50 | 11-15 pcw | F |
| 12960 | H376.IIB.51 | 11-15 pcw | F |
| 13060 | H376.IIB.52 | 11-15 pcw | F |

| | | | |
|---|---|---|---|
| 12820 | H376.IIIA.50 | 11-15 pcw | M |
| 12834 | H376.IIIA.51 | 11-15 pcw | F |
| 12888 | H376.IIIA.52 | 11-15 pcw | M |
| 12287 | H376.IIIB.50 | 16-20 pcw | M |
| 12837 | H376.IIIB.51 | 16-20 pcw | M |
| 12879 | H376.IIIB.52 | 16-20 pcw | M |
| 12880 | H376.IIIB.53 | 16-20 pcw | F |
| 12885 | H376.IV.53 | 16-20 pcw | F |
| 12365 | H376.IV.51 | 21-25 pcw | F |
| 12886 | H376.IV.54 | 21-25 pcw | M |
| 12288 | H376.IV.50 | 21-25 pcw | M |
| 12948 | H376.V.51 | 21-25 pcw | F |
| 12949 | H376.V.52 | 26-30 pcw | F |
| 12296 | H376.VI.50 | ≤1 year | M |
| 12977 | H376.VII.51 | ≤1 year | M |
| 12830 | H376.VIII.51 | ≤1 year | F |
| 12979 | H376.VIII.53 | 2-4 yrs | F |
| 12836 | H376.VIII.52 | 2-4 yrs | F |
| 12980 | H376.VIII.54 | 2-4 yrs | M |
| 12298 | H376.VIII.50 | 2-4 yrs | M |
| 12841 | H376.IX.51 | 5-10 yrs | M |
| 12831 | H376.X.51 | 11-15 yrs | F |
| 12299 | H376.X.50 | 11-15 yrs | M |
| 12984 | H376.X.53 | 15-20 yrs | M |
| 13057 | H376.XI.60 | 20-25 yrs | F |
| 12300 | H376.XI.50 | 20-25 yrs | M |
| 12290 | H376.XI.52 | 26-30 yrs | F |
| 12302 | H376.XI.53 | 35-40 yrs | M |
| 12303 | H376.XI.54 | 35-40 yrs | M |
| 12304 | H376.XI.56 | 35-40 yrs | F |

Abbreviations: pcw=post-conception weeks, yrs=years. *Nb: exact ages can be found on the BrainSpan website data, but cannot be revealed in the medRxiv version of the manuscript.*

#### Supplementary Table 3: Thalamic nuclei volume differences between young adult *C9orf72* repeat expansion carriers and non-carriers.

| Thalamic Nucleus | *ß* | SE | *t* | *t(-1)* | *p* |
|---|---|---|---|---|---|
| Left AV | -7.45 | 5.91 | -1.26 | 1.26 | 0.211 |
| Right AV | -2.5 | 4.93 | -0.51 | 0.51 | 0.613 |
| **Left LD** | **-3.93** | **1.88** | **-2.09** | **2.09** | **0.040*** |
| Right LD | -2.66 | 1.98 | -1.34 | 1.34 | 0.182 |

| | | | | | |
|---|---|---|---|---|---|
| **Left LP** | **-9.13** | **3.92** | **-2.33** | **2.33** | **0.022*** |
| Right LP | -6.24 | 4.26 | -1.47 | 1.47 | 0.146 |
| Left VA | -18.35 | 14.95 | -1.23 | 1.23 | 0.223 |
| Right VA | -5.65 | 11.62 | -0.49 | 0.49 | 0.628 |
| Left VAmc | -0.57 | 1.17 | -0.49 | 0.49 | 0.625 |
| Right VAmc | -0.54 | 1.02 | -0.53 | 0.53 | 0.598 |
| Left VLa | -14.74 | 17.6 | -0.84 | 0.84 | 0.405 |
| Right VLa | -2.93 | 15.24 | -0.19 | 0.19 | 0.848 |
| Left VLp | -10.71 | 21.61 | -0.5 | 0.50 | 0.621 |
| Right VLp | -1.76 | 20.27 | -0.09 | 0.09 | 0.931 |
| Left VPL | 4.07 | 27.4 | 0.15 | -0.15 | 0.882 |
| Right VPL | 9.28 | 26.44 | 0.35 | -0.35 | 0.727 |
| Left CeM | -2.58 | 2.50 | -1.03 | 1.03 | 0.305 |
| Right CeM | -3.11 | 2.09 | -1.49 | 1.49 | 0.139 |
| Left CL | 0.45 | 1.57 | 0.29 | -0.29 | 0.774 |
| Right CL | 0.62 | 1.63 | 0.38 | -0.38 | 0.704 |
| Left CM | 5.39 | 5.62 | 0.96 | -0.96 | 0.340 |
| Right CM | 5.79 | 5.49 | 1.06 | -1.06 | 0.294 |
| Left Pf | 1.95 | 1.66 | 1.18 | -1.18 | 0.243 |
| Right Pf | 2.64 | 1.71 | 1.54 | -1.54 | 0.127 |
| Left MVRe | -0.71 | 0.59 | -1.21 | 1.21 | 0.231 |
| Right MVRe | -1.01 | 0.55 | -1.83 | 1.83 | 0.071 |
| **Left MDm** | **-52.14** | **25.78** | **-2.02** | **2.02** | **0.046*** |
| **Right MDm** | **-61.89** | **20.94** | **-2.96** | **2.96** | **0.004**** |
| **Left MDl** | **-25.68** | **11.82** | **-2.17** | **2.17** | **0.032*** |
| **Right MDl** | **-23** | **8.56** | **-2.69** | **2.69** | **8.62E-03***** |
| Left LGN | -9.82 | 9.47 | -1.04 | 1.04 | 0.302 |
| Right LGN | -9.62 | 7.51 | -1.28 | 1.28 | 0.204 |
| Left MGN | -5.00 | 3.29 | -1.52 | 1.52 | 0.132 |
| Right MGN | -3.87 | 3.89 | -0.99 | 0.99 | 0.323 |
| Left LSg | -1.11 | 1.24 | -0.89 | 0.89 | 0.374 |
| Right LSg | -0.39 | 1.15 | -0.34 | 0.34 | 0.735 |
| **Left PuA** | **-19.23** | **7.35** | **-2.62** | **2.62** | **0.011*** |
| **Right PuA** | **-22.97** | **5.53** | **-4.16** | **4.16** | **7.57e-05***** |
| **Left PuM** | **-96.3** | **32.57** | **-2.96** | **2.96** | **4.00E-03***** |
| **Right PuM** | **-132.89** | **24.76** | **-5.37** | **5.37** | **6.54e-07***** |
| Left PuL | -6.91 | 6.35 | -1.09 | 1.09 | 0.28 |
| Right PuL | -2.29 | 6.01 | -0.38 | 0.38 | 0.704 |
| **Left PuI** | **-21.21** | **9.58** | **-2.22** | **2.22** | **0.029*** |
| **Right PuI** | **-23.97** | **6.96** | **-3.44** | **3.44** | **8.90E-04***** |

Only the main effect of group is shown for each thalamic nuclei model. A negative t-statistic reflects a smaller volume in the carrier group, and vice versa. To ease interpretation of downstream correlations with transcriptomic expression, imaging model t-statistics were multiplied by -1, such that larger values corresponded to larger effects of smaller nucleus volume in the carrier group. Thalamic nuclei with significantly smaller volumes in *C9orf72* repeat expansion carriers are bolded. *$p<0.05$, **$p<0.01$, ***$p<0.001$. Abbreviations: AV=anteroventral; LD=laterodorsal; LP=lateral posterior; VA=ventral anterior; VAmc=ventral anterior magnocellular; VLa=ventral lateral anterior; VLp=ventral lateral posterior; VPL=ventral posterolateral; CeM=central medial; CL=central lateral; CM=centromedian; Pf=parafascicular; MVRe=reuniens (medial ventral); MDm=mediodorsal medial magnocellular; MDl=mediodorsal lateral parvocellular; LGN=lateral geniculate; MGN=medial geniculate; LSg=limitans (suprageniculate); PuA=pulvinar anterior; PuM=pulvinar medial; PuL=pulvinar lateral; PuI=pulvinar inferior.

### Supplementary Table 4: Limma linear model statistics for normative spatial expression of NMF-derived transcriptomic components.

| Component | *ß* | *t* | *p* | Adjusted *p* |
|---|---|---|---|---|
| 7 | 0.240 | 3.00 | 0.00391 | 0.0391 |
| 1 | -0.142 | -1.72 | 0.090 | 0.450 |
| 9 | -0.144 | -1.46 | 0.151 | 0.502 |
| 10 | 0.075 | 0.97 | 0.337 | 0.841 |
| 3 | -0.056 | -0.54 | 0.591 | 0.841 |
| 6 | 0.038 | 0.45 | 0.653 | 0.841 |
| 2 | -0.032 | -0.38 | 0.702 | 0.841 |
| 5 | 0.034 | 0.35 | 0.726 | 0.841 |
| 4 | 0.037 | 0.31 | 0.757 | 0.841 |
| 8 | -0.016 | -0.20 | 0.843 | 0.843 |

Abbreviations: NMF=non-negative matrix factorization.

### Additional Details for Data-Driven Findings

A linear model fitted for *C9orf72* and the top 100 transcripts contributing to Component 7 revealed 76 transcripts that survived correction for multiple comparisons **(Suppl. Table 5)**. All 76 transcripts yielded higher normative expression in nuclei that have smaller volumes in *C9orf72* expansion carriers. The ten transcripts which showed highest normative expression in smaller nuclei ($p_{adj}$=0.021 for all) were: *DMBT1* (*ß*=0.070, *t*=3.44), *AARDC2* (*ß*=0.070, *t*=3.40), *C9orf3* (*ß*=0.068, *t*=3.35), *RAB7B* (*ß*=0.065, *t*=3.20), *ZBED3* (*ß*=0.065, *t*=3.17), *KIAA1147* (*ß*=0.064, *t*=3.12), *GALNT6* (*ß*=0.063, *t*=3.10), *ENPP6* (*ß*=0.062, *t*=3.06), *JPT2* (*ß*=0.061, *t*=3.00), and *FAM149B1* (*ß*=0.060, *t*=2.96).

Spearman-rank correlations of the top 100 transcripts with *C9orf72* yielded 70 that were positively associated with normative *C9orf72* expression across the thalamic nuclei, and which survived correction **(Fig. 3b; Suppl. Table 6)**. The top ten transcripts most strongly associated with *C9orf72* expression included: *AAMDC* ($\rho$=0.68, $p_{adj}$=5.68e-5), *GPRC5B* ($\rho$=0.66, $p_{adj}$=7.71e-5), *PPIF* ($\rho$=0.65, $p_{adj}$=1.01e-4), *TST* ($\rho$=0.61, $p_{adj}$=3.01e-4), *BCDIN3D* ($\rho$=0.58, $p_{adj}$=8.96e-4), *LDLRAD4* ($\rho$=0.57, $p_{adj}$=1.04e-3), *SELENOP* ($\rho$=0.57, $p_{adj}$=1.07e-3), *KLC1* ($\rho$=0.55, $p_{adj}$=1.55e-3), *FNTA* ($\rho$=0.55, $p_{adj}$=1.55e-3), and *CMTM5* ($\rho$=0.54, $p_{adj}$=1.55e-3).

Twenty transcripts showed significant associations between normative spatial expression patterns and thalamic nuclei volumes, but did not correlate with *C9orf72* expression: *C9orf3, ZBED3, KIAA1147, ENPP6, LOC101929384, TSPAN15, PHF10, PRKCQ, ARHGAP21, ANKRD65, SMARCC1, ABCA8, FLNC, NUDT1, ADAMTS14, LRP6, LOC100507487, SSFA2, TMTC4, CRTAP*. The normative expression of 13 transcripts correlated significantly with *C9orf72*, but were not associated with thalamic nuclei volume patterns: *AAMDC, PPIF, TST, PLEKHB, SEPT4, DENND5A, MOBP, HIPK2, IGFLR1, FRMD5, BCAR3, S100B, ICK*.

#### Supplementary Table 5: Weights of all 1564 transcripts that contribute to Component 7.

| Transcript | Weight |
|---|---|
| *FAM149B1* | 0.27732562 |
| *C9orf3* | 0.27482445 |
| *SCRG1* | 0.26745554 |
| *TMTC2* | 0.26674147 |
| *B3GNT9* | 0.264857 |
| *ZBED3* | 0.26472537 |
| *ABCA8* | 0.26416993 |
| *IFIT2* | 0.26268188 |
| *BCDIN3D* | 0.26213538 |
| *SLC31A2* | 0.26174864 |
| *TMEM98* | 0.26071185 |
| *ANKRD65* | 0.26055357 |
| *CTNNA3* | 0.26004657 |
| *KLC1* | 0.26001585 |
| *SLC5A11* | 0.25955889 |
| *PAIP2B* | 0.25913759 |
| *KIAA1147* | 0.25846573 |
| *USP54* | 0.25802954 |
| *FLNC* | 0.25771528 |
| *S100B* | 0.25689834 |
| *TMTC4* | 0.25630338 |
| *MAG* | 0.2558033 |
| *HHATL* | 0.25573933 |
| *RASSF2* | 0.25400119 |
| *SELENOP* | 0.25361016 |
| *CLDND1* | 0.2530866 |
| *CAPN3* | 0.25280057 |
| *ADAMTS4* | 0.2527793 |
| *ADGRE5* | 0.25272123 |
| *NDE1* | 0.25268746 |
| *JPT2* | 0.25246559 |
| *MYLK* | 0.25230027 |
| *PHF10* | 0.25226564 |
| *FNTA* | 0.25225914 |
| *LRP6* | 0.2520635 |
| *FAT1* | 0.25183494 |

| | |
|---|---|
| *SAP30* | 0.25131094 |
| *ARHGEF1* | 0.25106525 |
| *PPIF* | 0.25042049 |
| *CTDSP1* | 0.25039235 |
| *CMTM5* | 0.25033878 |
| *LOC100506725* | 0.25028307 |
| *ARRDC2* | 0.24977591 |
| *PCSK6* | 0.24974853 |
| *PCBP4* | 0.24972434 |
| *LOC100507487* | 0.24945738 |
| *FEZ1* | 0.24929638 |
| *SESTD1* | 0.24881476 |
| *PARP4* | 0.24756606 |
| *RDH11* | 0.24638722 |
| *BAMBI* | 0.24561791 |
| *MOBP* | 0.24501782 |
| *ADAMTS14* | 0.24458798 |
| *CDH20* | 0.24421889 |
| *ENPP6* | 0.24404415 |
| *ARHGAP21* | 0.24398703 |
| *LOC101929384* | 0.24392309 |
| *TSPAN15* | 0.2438286 |
| *CHST6* | 0.24356703 |
| *SEPT4* | 0.24348934 |
| *CARNS1* | 0.24342159 |
| *USH1C* | 0.24330783 |
| *HIPK2* | 0.24283744 |
| *KATNA1* | 0.24224379 |
| *IGFLR1* | 0.24179587 |
| *TCFL5* | 0.24175696 |
| *SSFA2* | 0.24161088 |
| *FAM19A4* | 0.24124283 |
| *ASPRV1* | 0.2411449 |
| *TNS2* | 0.24081517 |
| *ZNF664* | 0.24061932 |
| *BCAR3* | 0.24043001 |
| *QDPR* | 0.24014073 |
| *HSD17B3* | 0.23947707 |
| *PRKCQ* | 0.23928255 |
| *SFR1* | 0.23851498 |
| *CRTAP* | 0.23836815 |
| *TSPAN8* | 0.23828912 |
| *ADARB2.AS1* | 0.23816012 |

| | |
|---|---|
| *ICK* | 0.2381278 |
| *KHNYN* | 0.2381137 |
| *TMEM140* | 0.23805644 |
| *NUDT1* | 0.23778328 |
| *LDLRAD4* | 0.23754055 |
| *TST* | 0.23718527 |
| *SMARCC1* | 0.23711232 |
| *RAB7B* | 0.23693162 |
| *AAMDC* | 0.23687485 |
| *RGS1* | 0.236782 |
| *ARHGEF37* | 0.23677367 |
| *GPRC5B* | 0.23666414 |
| *DENND5A* | 0.23641996 |
| *DMBT1* | 0.23633421 |
| *LLGL2* | 0.23632347 |
| *PLEKHB1* | 0.23621608 |
| *GALNT6* | 0.23604551 |
| *RCOR2* | 0.2354474 |
| *LAP3* | 0.23519802 |
| *FRMD5* | 0.23495933 |
| *VKORC1* | 0.2346181 |
| *CCEPR* | 0.23415486 |
| *MTRR* | 0.23398236 |
| *ADARB2* | 0.23363814 |
| *MCM7* | 0.23338429 |
| *HSD11B1* | 0.2333415 |
| *HAGLROS* | 0.23308011 |
| *TULP4* | 0.23292598 |
| *CNKSR3* | 0.23272004 |
| *SLC6A8* | 0.23210044 |
| *SEC22C* | 0.23205155 |
| *ARHGAP22* | 0.2316935 |
| *ANXA5* | 0.2312351 |
| *FUT8* | 0.22972085 |
| *NDRG1* | 0.22958441 |
| *C11orf54* | 0.22954727 |
| *MBOAT1* | 0.22945996 |
| *TGFB3* | 0.22941888 |
| *LINC01158* | 0.22915622 |
| *MBOAT2* | 0.22912644 |
| *COL16A1* | 0.22910137 |
| *GCLC* | 0.22850485 |
| *LOC100506544* | 0.22825631 |

| | |
|---|---|
| *CHD1L* | 0.22809293 |
| *KANK4* | 0.22765703 |
| *PTPRH* | 0.22750302 |
| *CRNDE* | 0.22738384 |
| *PLAG1* | 0.22729482 |
| *LINC00674* | 0.22699056 |
| *FPR1* | 0.22688608 |
| *KLF3.AS1* | 0.226518 |
| *ACP5* | 0.22635509 |
| *PTCD3* | 0.22566734 |
| *HMBOX1* | 0.2254948 |
| *CPQ* | 0.22542696 |
| *CRYL1* | 0.2252745 |
| *ABTB2* | 0.22522483 |
| *DIP2B* | 0.22491301 |
| *SEMA4D* | 0.22438472 |
| *C6orf48* | 0.22361975 |
| *YIF1A* | 0.22356354 |
| *ERBB2* | 0.22312456 |
| *ABHD6* | 0.22296124 |
| *SERHL2* | 0.22281333 |
| *METAP2* | 0.22271724 |
| *UNC5B* | 0.22250342 |
| *CCNB3* | 0.22227652 |
| *ITGB4* | 0.22198072 |
| *PLLP* | 0.22179212 |
| *SETD9* | 0.22169128 |
| *PLA2G4C* | 0.22159936 |
| *G6PC* | 0.22123848 |
| *ST3GAL4* | 0.22116133 |
| *PCCB* | 0.22115354 |
| *FOXO4* | 0.22105435 |
| *PROX1* | 0.22090864 |
| *PLPP1* | 0.22027108 |
| *PSRC1* | 0.22015404 |
| *NPAS3* | 0.2196469 |
| *LOC390705* | 0.21933239 |
| *PAQR6* | 0.21921096 |
| *STXBP6* | 0.21902979 |
| *RAB3IP* | 0.21863996 |
| *RBBP9* | 0.21843505 |
| *CDK2AP1* | 0.21840756 |
| *SNX31* | 0.2178085 |

| | |
|---|---|
| *SERPINA3* | 0.21774212 |
| *ATP6V0E1* | 0.21746768 |
| *TTC23* | 0.21691954 |
| *TDP1* | 0.21661648 |
| *RHCE* | 0.2164951 |
| *LAMA2* | 0.21627769 |
| *ADAMTS7* | 0.21619534 |
| *CHRNA2* | 0.21511454 |
| *TRIP6* | 0.21510115 |
| *CDKN1B* | 0.214683 |
| *TAS2R20* | 0.21455776 |
| *LOC100286906* | 0.21449606 |
| *TLN1* | 0.21438714 |
| *MYH9* | 0.21371815 |
| *RAB33A* | 0.21325007 |
| *EIF3J* | 0.21291501 |
| *PRDX4* | 0.21278874 |
| *TSC22D4* | 0.21246607 |
| *COX20* | 0.21199745 |
| *TBL1X* | 0.21172829 |
| *SH3GL3* | 0.21101697 |
| *TMEM201* | 0.2108123 |
| *WIPI1* | 0.21075558 |
| *COX14* | 0.21064115 |
| *GATAD1* | 0.21050486 |
| *TIMELESS* | 0.21028074 |
| *PHGDH* | 0.21020052 |
| *LRRC58* | 0.21011302 |
| *CUTC* | 0.20996408 |
| *LAPTM4B* | 0.20976363 |
| *FAM228B* | 0.20950128 |
| *PLEKHG1* | 0.20943629 |
| *C21orf62.AS1* | 0.20943144 |
| *LGR5* | 0.2093988 |
| *SLC35D2* | 0.20870643 |
| *ASCC1* | 0.2086266 |
| *PRDX1* | 0.20850512 |
| *SAMD4A* | 0.20844928 |
| *IP6K2* | 0.20827588 |
| *ZNF189* | 0.20823695 |
| *TET1* | 0.20815234 |
| *CASC3* | 0.20809132 |
| *SRP14.AS1* | 0.20754392 |

| | |
|---|---|
| *PIK3IP1* | 0.20745243 |
| *HEATR6* | 0.2074205 |
| *SYNGR2* | 0.2073554 |
| *MGA* | 0.20704325 |
| *DTNB* | 0.20697117 |
| *USP40* | 0.20686838 |
| *GRIK4* | 0.20515516 |
| *LAMA4* | 0.20504381 |
| *EEF1D* | 0.20499736 |
| *SLC2A4RG* | 0.20428369 |
| *COLGALT2* | 0.20391279 |
| *FOXN3* | 0.20367311 |
| *P3H2* | 0.20334975 |
| *PAAF1* | 0.20334957 |
| *ST3GAL6.AS1* | 0.20308876 |
| *LIG1* | 0.20298852 |
| *PGBD2* | 0.20288033 |
| *SNX30* | 0.20284286 |
| *UXT* | 0.20244497 |
| *TMEM99* | 0.20216711 |
| *MID1IP1* | 0.2019514 |
| *PIN4P1* | 0.2018656 |
| *EPM2A* | 0.20156373 |
| *TMEM79* | 0.20140419 |
| *TNRC6C* | 0.20082205 |
| *NFASC* | 0.20062454 |
| *AGTRAP* | 0.2003934 |
| *STK39* | 0.19982233 |
| *RASSF8.AS1* | 0.19971536 |
| *DEFB1* | 0.19873934 |
| *PMF1* | 0.19853358 |
| *HIRIP3* | 0.19744369 |
| *SPEM2* | 0.19742905 |
| *LOC440311* | 0.19729396 |
| *SPART* | 0.19662512 |
| *TLR2* | 0.19638968 |
| *NFIA* | 0.19633128 |
| *GAD1* | 0.19612868 |
| *ALKBH2* | 0.19591265 |
| *ACAN* | 0.19565829 |
| *NPB* | 0.19519898 |
| *TP53I3* | 0.19507377 |
| *DNAJB2* | 0.19506561 |

| | |
|---|---|
| *TMEM50B* | 0.19492164 |
| *GAD2* | 0.19482417 |
| *NFATC3* | 0.19481884 |
| *RBM8A* | 0.19472258 |
| *JAKMIP3* | 0.19439942 |
| *FER* | 0.1943295 |
| *ATG3* | 0.19427585 |
| *GTF2IRD1* | 0.19419619 |
| *PMFBP1* | 0.19383457 |
| *CTSL* | 0.19383249 |
| *SLC4A2* | 0.19381389 |
| *SLC16A8* | 0.1937181 |
| *PHLDA3* | 0.19354676 |
| *RB1* | 0.19314126 |
| *UPF3A* | 0.19310817 |
| *DTX2* | 0.19266789 |
| *ACY3* | 0.1924026 |
| *CTBP2* | 0.19201286 |
| *HLA.DQB1* | 0.19159253 |
| *OPRM1* | 0.19153189 |
| *TIMMDC1* | 0.1906997 |
| *KXD1* | 0.19063292 |
| *TP53INP1* | 0.19062287 |
| *HSPB8* | 0.19055599 |
| *CEBPD* | 0.19001284 |
| *NFKBIA* | 0.18909683 |
| *SLC35E3* | 0.18884016 |
| *RPS6* | 0.18863091 |
| *RPS6KA2* | 0.18834814 |
| *SCAMP2* | 0.18826653 |
| *MEIS2* | 0.18824367 |
| *GAREM1* | 0.18776046 |
| *SLC32A1* | 0.18743412 |
| *SETD7* | 0.18669358 |
| *CD63* | 0.18589085 |
| *FAM210A* | 0.1858604 |
| *SYMPK* | 0.18583668 |
| *ECHDC1* | 0.1857456 |
| *KCTD11* | 0.18574457 |
| *TTN* | 0.18483162 |
| *HLA.A* | 0.18464532 |
| *MIS12* | 0.18459145 |
| *RABGGTB* | 0.18416431 |

| | |
|---|---|
| *DAZAP1* | 0.18404891 |
| *DPYSL2* | 0.18363777 |
| *ATP8A1* | 0.18332567 |
| *AIMP1* | 0.18323904 |
| *FAM96B* | 0.18318722 |
| *CSE1L.AS1* | 0.18258327 |
| *RGS16* | 0.18241418 |
| *PSMD9* | 0.18238127 |
| *IDI2.AS1* | 0.18225229 |
| *SCOC.AS1* | 0.18214295 |
| *CRB2* | 0.18190117 |
| *NPR3* | 0.18175056 |
| *C8orf31* | 0.18171386 |
| *MRPL22* | 0.18122785 |
| *HACD2* | 0.18109099 |
| *MRE11* | 0.18105936 |
| *LOC100132831* | 0.18099801 |
| *HNRNPA2B1* | 0.18067192 |
| *SCAMP1.AS1* | 0.18055888 |
| *DHRS4* | 0.18026686 |
| *CEP78* | 0.17969207 |
| *RPL17* | 0.17953167 |
| *BAG1* | 0.17919827 |
| *NOP53* | 0.17919745 |
| *ANKHD1* | 0.1791016 |
| *CNIH4* | 0.17830073 |
| *KRTCAP3* | 0.17822152 |
| *FIS1* | 0.17783676 |
| *GRHPR* | 0.17781075 |
| *CLNS1A* | 0.17769065 |
| *PPFIA1* | 0.17731906 |
| *LCAT* | 0.17722702 |
| *PIEZO1* | 0.17643031 |
| *PLIN2* | 0.17624159 |
| *CKMT2* | 0.17619743 |
| *PSMA2* | 0.17611376 |
| *MOCS1* | 0.17586979 |
| *RFESD* | 0.17563168 |
| *PHYHD1* | 0.17543212 |
| *RANBP17* | 0.17520848 |
| *PNO1* | 0.1748215 |
| *SELENOS* | 0.1740371 |
| *RAP1GDS1* | 0.17362155 |

| | |
|---|---|
| *JAM2* | 0.17353538 |
| *ZNHIT6* | 0.17346682 |
| *SAMD11* | 0.17334769 |
| *MMACHC* | 0.17322887 |
| *MGAT5* | 0.17312404 |
| *GNAI2* | 0.17263017 |
| *SMIM1* | 0.17238798 |
| *HMGCL* | 0.17216718 |
| *EIF4B* | 0.17202371 |
| *CPAMD8* | 0.17178345 |
| *TRADD* | 0.17169101 |
| *EIF2AK3* | 0.17151557 |
| *SLAMF7* | 0.17125502 |
| *RETREG1* | 0.17107622 |
| *HEPN1* | 0.17105802 |
| *KLHL21* | 0.17092339 |
| *TMEM80* | 0.1706864 |
| *TMEM116* | 0.17062007 |
| *ZNF727* | 0.17056573 |
| *P3H1* | 0.17012507 |
| *TBL1Y* | 0.16995713 |
| *SURF4* | 0.16995244 |
| *NINJ1* | 0.16995061 |
| *SST* | 0.16986271 |
| *FAM126A* | 0.16963962 |
| *ACSS2* | 0.1695378 |
| *KCTD20* | 0.16892533 |
| *MYH3* | 0.16874894 |
| *EML2* | 0.16871084 |
| *PLCB4* | 0.16854013 |
| *L3MBTL4* | 0.16806755 |
| *TP73.AS1* | 0.16801062 |
| *TRIM47* | 0.16738063 |
| *KSR1* | 0.1671246 |
| *SEC23B* | 0.167004 |
| *LSM2* | 0.16700382 |
| *RHBDF1* | 0.16699405 |
| *SNAPIN* | 0.16635865 |
| *ASB4* | 0.16626639 |
| *HERC4* | 0.16615583 |
| *NAPEPLD* | 0.16583448 |
| *ANKRD18A* | 0.16574035 |
| *PSMC3IP* | 0.16547061 |

| | |
|---|---|
| *SLC7A14* | 0.16523815 |
| *SMG6* | 0.16515779 |
| *PLD5* | 0.16498806 |
| *TINF2* | 0.16497307 |
| *WRN* | 0.16434142 |
| *EIF4A3* | 0.16428186 |
| *TMEM14C* | 0.16379867 |
| *CHRNA4* | 0.16379223 |
| *PIK3AP1* | 0.16377769 |
| *GREB1L* | 0.16363905 |
| *FAM96A* | 0.16352133 |
| *CD109* | 0.16322015 |
| *C21orf33* | 0.16298891 |
| *SPATA1* | 0.1627326 |
| *PEX11G* | 0.16235272 |
| *ZNF580* | 0.16181794 |
| *NR3C1* | 0.16167774 |
| *C22orf46* | 0.16148352 |
| *MIR7515HG* | 0.16143293 |
| *NUP155* | 0.1610647 |
| *PEX7* | 0.1602154 |
| *GGNBP1* | 0.16008803 |
| *DUSP1* | 0.15992653 |
| *GSTT1* | 0.15942517 |
| *SUMF2* | 0.15939987 |
| *SLC25A5.AS1* | 0.15928254 |
| *MRPS14* | 0.15923081 |
| *FAM122B* | 0.15916809 |
| *SLC24A2* | 0.15913375 |
| *A4GALT* | 0.15910388 |
| *ATP9B* | 0.15899513 |
| *REPIN1* | 0.15887881 |
| *MUM1* | 0.15876934 |
| *RPL23AP7* | 0.15835641 |
| *ZNF185* | 0.1581058 |
| *MIF4GD* | 0.15795352 |
| *RAB4A* | 0.15793378 |
| *CREBRF* | 0.15757257 |
| *CHRNA3* | 0.15736412 |
| *DOK7* | 0.15717451 |
| *HERPUD1* | 0.15690013 |
| *RPS27* | 0.15681284 |
| *MGC34796* | 0.15678384 |

| | |
|---|---|
| *PTPRE* | 0.15677652 |
| *THNSL2* | 0.15658017 |
| *LUC7L* | 0.15607905 |
| *GPR179* | 0.1559596 |
| *CGB1* | 0.15586864 |
| *SCAMP3* | 0.1555633 |
| *GCC1* | 0.15531074 |
| *GNA14* | 0.15451615 |
| *RPL19* | 0.15441389 |
| *MFSD11* | 0.15439772 |
| *SMPD1* | 0.15431552 |
| *LGALS1* | 0.15424511 |
| *RALY* | 0.15364891 |
| *EIF4EBP2* | 0.15355957 |
| *RPS11* | 0.15351668 |
| *PTPRT* | 0.15338339 |
| *FAM9B* | 0.15313992 |
| *HCG18* | 0.15301796 |
| *CYB5A* | 0.15291102 |
| *SPR* | 0.15279994 |
| *TBC1D9B* | 0.15225317 |
| *RPL23AP82* | 0.15221696 |
| *KIRREL3* | 0.15215098 |
| *ZBTB2* | 0.15188588 |
| *TTPA* | 0.15169415 |
| *GABARAPL2* | 0.15154229 |
| *FGF13* | 0.15143962 |
| *GRM8* | 0.15135405 |
| *LDLR* | 0.15127056 |
| *USP10* | 0.15114292 |
| *TCF7L2* | 0.15101088 |
| *CHI3L1* | 0.1509128 |
| *ZIC1* | 0.15017152 |
| *EEF1G* | 0.14982639 |
| *GPR4* | 0.1496657 |
| *ECH1* | 0.14964256 |
| *HRH3* | 0.14943496 |
| *LOC148413* | 0.14940226 |
| *CNPY2* | 0.14928696 |
| *LGI2* | 0.14887679 |
| *GAL* | 0.14815796 |
| *LOC100506990* | 0.14758941 |
| *SIGLEC11* | 0.14756084 |

| | |
|---|---|
| *PRDM5* | 0.14715346 |
| *C14orf119* | 0.14695671 |
| *FUNDC2* | 0.14683513 |
| *RPS12* | 0.14680089 |
| *PNOC* | 0.14658954 |
| *MSANTD4* | 0.14653165 |
| *SULF2* | 0.14634362 |
| *ZNF330* | 0.1462759 |
| *AKAIN1* | 0.14611089 |
| *SDHAF2* | 0.14483121 |
| *LINC01881* | 0.14383635 |
| *DPYSL3* | 0.14317367 |
| *PCGF5* | 0.1426266 |
| *GDI2* | 0.14190332 |
| *CCDC115* | 0.1416389 |
| *OBSCN* | 0.14122412 |
| *CNPPD1* | 0.14107994 |
| *KCNS3* | 0.1405876 |
| *CDADC1* | 0.14025533 |
| *C1orf43* | 0.14024336 |
| *ST8SIA4* | 0.14011462 |
| *EPN3* | 0.14006498 |
| *TCN2* | 0.13995059 |
| *SLC6A1* | 0.13979929 |
| *PCDHB4* | 0.1397056 |
| *PTPN21* | 0.13962067 |
| *NAGK* | 0.13934275 |
| *PDCL3P4* | 0.13931726 |
| *MED12* | 0.13878326 |
| *CACNA2D2* | 0.13853409 |
| *PXDC1* | 0.13833287 |
| *EPRS* | 0.13823253 |
| *PPA1* | 0.13773669 |
| *PRKCH* | 0.13763955 |
| *VAV3* | 0.13703389 |
| *BCL2L1* | 0.13702281 |
| *AFG1L* | 0.13695076 |
| *UBE2A* | 0.13678099 |
| *SPOCK1* | 0.1367674 |
| *C1orf174* | 0.13614189 |
| *TTC37* | 0.13600331 |
| *SHPRH* | 0.13587131 |
| *WDYHV1* | 0.13568121 |

| | |
|---|---|
| *PRCP* | 0.13485565 |
| *FAM35A* | 0.13474742 |
| *EPHA1* | 0.1344427 |
| *ZKSCAN2* | 0.13391148 |
| *KCNC2* | 0.13370926 |
| *SWT1* | 0.13342525 |
| *RAMP1* | 0.13329129 |
| *SLC25A10* | 0.13309058 |
| *SLC12A7* | 0.13301648 |
| *FOXP4* | 0.13299265 |
| *DLK1* | 0.13265361 |
| *LIPC* | 0.13252014 |
| *GPR108* | 0.13226579 |
| *DENND2D* | 0.13214318 |
| *LAMTOR5* | 0.13214073 |
| *ACAT1* | 0.13195041 |
| *RPL8* | 0.13163971 |
| *ADAM19* | 0.13149005 |
| *RNF10* | 0.13144396 |
| *PRSS23* | 0.13124725 |
| *TMEM255A* | 0.13124499 |
| *ALK* | 0.13113098 |
| *ATRIP* | 0.13051463 |
| *CD83* | 0.13037655 |
| *SNRPB2* | 0.13010871 |
| *PIP4P2* | 0.13009939 |
| *SYNJ2BP* | 0.13002817 |
| *PCSK5* | 0.12989797 |
| *NR2F1.AS1* | 0.12986283 |
| *CUL4A* | 0.12980261 |
| *SLC25A5* | 0.12967407 |
| *TBL3* | 0.12940817 |
| *SUCLG1* | 0.12932463 |
| *VAT1L* | 0.12930897 |
| *GALNS* | 0.12898918 |
| *ISOC2* | 0.12893375 |
| *TMEM41A* | 0.12871959 |
| *RPL13* | 0.12854983 |
| *ACTC1* | 0.12854094 |
| *PAIP2* | 0.12841151 |
| *PRKX* | 0.12813299 |
| *RNF5* | 0.12808882 |
| *RPLP0* | 0.12805834 |

| | |
|---|---|
| *ZMAT4* | 0.12758536 |
| *SLC6A12* | 0.12709035 |
| *S100A6* | 0.12708916 |
| *SAMD5* | 0.12691796 |
| *CDPF1* | 0.1259337 |
| *QRFPR* | 0.12531009 |
| *RNF128* | 0.12514285 |
| *SEMA3G* | 0.12448448 |
| *PHYHIPL* | 0.12439371 |
| *ISL1* | 0.12411231 |
| *OSBPL5* | 0.12411016 |
| *GSTO2* | 0.12391681 |
| *NXF1* | 0.12355809 |
| *MRPL53* | 0.12353775 |
| *RSRC1* | 0.12339602 |
| *RASSF3* | 0.1233392 |
| *FGFBP3* | 0.12333186 |
| *SLITRK6* | 0.12301603 |
| *PCBP3* | 0.12236227 |
| *GNG13* | 0.12229198 |
| *GRM7* | 0.12200314 |
| *SURF1* | 0.12105403 |
| *RORA* | 0.12104991 |
| *TBK1* | 0.12082397 |
| *DNASE1* | 0.12071345 |
| *ELMO1* | 0.12069604 |
| *EGFEM1P* | 0.12022911 |
| *MIS18A* | 0.12016344 |
| *MYL6* | 0.11961893 |
| *PAQR9* | 0.11933672 |
| *ZNF45* | 0.11832906 |
| *MAB21L2* | 0.11762158 |
| *KCNMB2* | 0.11756696 |
| *RNF149* | 0.11749137 |
| *TMEM65* | 0.11745373 |
| *SNTG2* | 0.1174044 |
| *RALGAPA2* | 0.11737069 |
| *RPS28* | 0.1167469 |
| *EPB41L1* | 0.11648434 |
| *TMEM132C* | 0.11621186 |
| *GRID2* | 0.11609502 |
| *KCNJ5* | 0.1160327 |
| *PDGFRL* | 0.11594843 |

| | |
|---|---|
| *DDIT4L* | 0.11561008 |
| *MYOZ3* | 0.11542876 |
| *UTRN* | 0.1150926 |
| *COL10A1* | 0.11484167 |
| *PDLIM3* | 0.11458903 |
| *LOC100132057* | 0.11432626 |
| *THUMPD2* | 0.11399032 |
| *NDUFB2* | 0.11381766 |
| *ARHGAP6* | 0.11377071 |
| *PIGS* | 0.11362816 |
| *CEP57* | 0.11341295 |
| *TMA7* | 0.11214302 |
| *KIFC3* | 0.11206795 |
| *CECR7* | 0.11125737 |
| *NOS1* | 0.1111079 |
| *ADAMTSL3* | 0.11083515 |
| *WDHD1* | 0.11009689 |
| *ANKRD35* | 0.11008297 |
| *DCDC1* | 0.11001719 |
| *MRFAP1* | 0.10959067 |
| *UBTD1* | 0.10954725 |
| *PSAP* | 0.10923837 |
| *GPX3* | 0.10879013 |
| *TLE6* | 0.10874058 |
| *BCKDHA* | 0.1086628 |
| *SCAPER* | 0.1085769 |
| *CASP7* | 0.10786991 |
| *FAM204A* | 0.1077104 |
| *ANXA3* | 0.10754784 |
| *ESD* | 0.10688286 |
| *S100A8* | 0.10641178 |
| *NT5DC2* | 0.10634465 |
| *RNF4* | 0.10611949 |
| *MORF4L1* | 0.10592998 |
| *ABHD12B* | 0.1057684 |
| *C1QTNF2* | 0.10559668 |
| *KLHL41* | 0.10547645 |
| *ZMIZ1* | 0.10536798 |
| *CTSD* | 0.10530232 |
| *TRPC3* | 0.10528542 |
| *ZDHHC7* | 0.10484442 |
| *BTC* | 0.10454733 |
| *IDI1* | 0.10452857 |

| | |
|---|---|
| *KLHDC10* | 0.10437266 |
| *TMEM243* | 0.1042474 |
| *GNAS* | 0.10421657 |
| *RPRD2* | 0.10419196 |
| *SMPX* | 0.1037388 |
| *MYO1B* | 0.10372314 |
| *KCNIP1* | 0.10342126 |
| *VAV1* | 0.10325861 |
| *NYNRIN* | 0.10323432 |
| *GOLGA2* | 0.10294141 |
| *CCDC149* | 0.10273351 |
| *RESP18* | 0.1026041 |
| *KIAA0100* | 0.1024164 |
| *NFIB* | 0.10237735 |
| *ANK1* | 0.1023196 |
| *C11orf87* | 0.10213951 |
| *VASN* | 0.10189568 |
| *KRT18* | 0.10153614 |
| *ZNF585A* | 0.10135804 |
| *C10orf82* | 0.10111892 |
| *ISCU* | 0.10094458 |
| *GPKOW* | 0.10055515 |
| *TSC22D3* | 0.10032898 |
| *WTIP* | 0.10021182 |
| *EPCAM* | 0.10020322 |
| *PPP1R17* | 0.09997593 |
| *TCEAL3* | 0.09954461 |
| *SP5* | 0.09942705 |
| *TMEM242* | 0.09904622 |
| *MCUB* | 0.09902873 |
| *FAM174B* | 0.09895179 |
| *PENK* | 0.09886253 |
| *HGSNAT* | 0.09882361 |
| *GPAA1* | 0.09872526 |
| *MATR3* | 0.09866832 |
| *RPL6* | 0.09865805 |
| *EVPL* | 0.09849063 |
| *SGSH* | 0.09843048 |
| *EMC4* | 0.09816821 |
| *UTP14C* | 0.09796581 |
| *GPR137C* | 0.09771304 |
| *TMEM62* | 0.09749595 |
| *FAM155B* | 0.09743529 |

| | |
|---|---|
| *MTRF1L* | 0.09728414 |
| *AREL1* | 0.09717271 |
| *ARFGAP2* | 0.09710094 |
| *MAGED2* | 0.097087 |
| *KCNN4* | 0.09687741 |
| *RNASEL* | 0.09684368 |
| *SDHA* | 0.09677364 |
| *AUP1* | 0.09654894 |
| *GPC6* | 0.09623899 |
| *HS3ST1* | 0.09595026 |
| *MPHOSPH8* | 0.09557255 |
| *POMZP3* | 0.0954728 |
| *DEDD* | 0.0953033 |
| *CSRP2* | 0.09525938 |
| *ITGA7* | 0.09510865 |
| *TCEAL6* | 0.09488997 |
| *GUCY1A1* | 0.09486793 |
| *CLIC5* | 0.09425459 |
| *KRT31* | 0.09415159 |
| *GALM* | 0.09393885 |
| *GRIP2* | 0.09386845 |
| *TXN2* | 0.09376312 |
| *KIAA1468* | 0.09355272 |
| *SENP2* | 0.09315031 |
| *PIGH* | 0.09267707 |
| *SCPEP1* | 0.09260024 |
| *GRAP* | 0.09256021 |
| *GPC4* | 0.0924363 |
| *LOXL1* | 0.09234542 |
| *HDDC2* | 0.09232779 |
| *AFAP1L2* | 0.09230138 |
| *TMEM8B* | 0.09213615 |
| *JMJD4* | 0.09195509 |
| *TNFAIP2* | 0.0918778 |
| *NDUFS2* | 0.09168498 |
| *PTH1R* | 0.09160017 |
| *HNRNPLL* | 0.09159239 |
| *DNAJA3* | 0.09143233 |
| *INAFM1* | 0.09128498 |
| *CASP5* | 0.09126109 |
| *CYTH2* | 0.09118145 |
| *ADAMTSL1* | 0.09108996 |
| *CITED2* | 0.09088593 |

| | |
|---|---|
| *TSPAN12* | 0.09087349 |
| *PPP3CC* | 0.09061576 |
| *NIP7* | 0.09055506 |
| *PRKAG1* | 0.09047553 |
| *OPA3* | 0.09041551 |
| *KIT* | 0.090373 |
| *AKR1C3* | 0.09032745 |
| *C1QTNF4* | 0.09011056 |
| *UBAP2* | 0.09007506 |
| *MDFIC* | 0.09005041 |
| *MARC1* | 0.09001965 |
| *CITED4* | 0.08990015 |
| *PCCA* | 0.08979322 |
| *RDH12* | 0.08976842 |
| *ASCL2* | 0.08953371 |
| *SLA* | 0.08942023 |
| *PIGX* | 0.08940852 |
| *TMOD3* | 0.08938802 |
| *ANXA2P1* | 0.08935743 |
| *ASB5* | 0.08920571 |
| *CDC26* | 0.08895966 |
| *SLC30A10* | 0.08888819 |
| *RIMKLB* | 0.08884217 |
| *FOXP2* | 0.08866379 |
| *VWC2* | 0.08862158 |
| *CTNNB1* | 0.08805579 |
| *PLN* | 0.08804701 |
| *MRPS23* | 0.08800529 |
| *CYR61* | 0.08779199 |
| *PHB* | 0.08768553 |
| *ZSCAN2* | 0.08746705 |
| *ZDHHC22* | 0.0871517 |
| *FGD5* | 0.08708873 |
| *METTL1* | 0.0869878 |
| *C12orf57* | 0.08698765 |
| *EFEMP2* | 0.08682549 |
| *MCC* | 0.08678745 |
| *FLNA* | 0.08653815 |
| *CARTPT* | 0.08644706 |
| *RRS1* | 0.08636334 |
| *VSIG8* | 0.08634965 |
| *LRPPRC* | 0.0861343 |
| *LRRC66* | 0.08579642 |

| | |
|---|---|
| *SAXO2* | 0.08573973 |
| *GSR* | 0.0855218 |
| *TRIQK* | 0.08543967 |
| *TMEM45B* | 0.08529557 |
| *ACTG2* | 0.08526212 |
| *TMEM173* | 0.08515995 |
| *STK35* | 0.08512115 |
| *SLC26A11* | 0.08509099 |
| *ADRA1B* | 0.08500736 |
| *SLC2A13* | 0.08486109 |
| *SLC44A5* | 0.08485077 |
| *KCNJ12* | 0.08473608 |
| *HBG1* | 0.08412796 |
| *SLC47A1* | 0.08409748 |
| *PCP4L1* | 0.08404749 |
| *CD247* | 0.08399032 |
| *B3GLCT* | 0.08397985 |
| *NPNT* | 0.08389625 |
| *RFLNB* | 0.08375258 |
| *BSPRY* | 0.0834918 |
| *ASIC4* | 0.08312309 |
| *TRIB1* | 0.08296953 |
| *PPT1* | 0.08286031 |
| *FAM19A5* | 0.08264283 |
| *ANOS1* | 0.08251499 |
| *LOC100419583* | 0.08219551 |
| *UBE2L6* | 0.08218481 |
| *EXTL2* | 0.0820482 |
| *ULK4* | 0.0817611 |
| *CHST2* | 0.08144428 |
| *ADRA1D* | 0.0813231 |
| *ZNF280B* | 0.08116183 |
| *NSDHL* | 0.08084569 |
| *INPP5A* | 0.08073078 |
| *C7orf57* | 0.08053254 |
| *MEI1* | 0.08051977 |
| *SPTSSB* | 0.08048889 |
| *SNX12* | 0.08026616 |
| *TCF4* | 0.08021458 |
| *FDPSP2* | 0.08021179 |
| *ARPC1B* | 0.07993034 |
| *SCAMP1* | 0.07985337 |
| *FAM20A* | 0.07984047 |

| | |
|---|---|
| *ZNF251* | 0.07909762 |
| *DIRAS3* | 0.07877561 |
| *ZNF667* | 0.07864669 |
| *HSF1* | 0.07848719 |
| *SKAP1* | 0.07833321 |
| *KCNK3* | 0.07809027 |
| *CNTN4* | 0.0779242 |
| *GPR83* | 0.07780191 |
| *SPECC1L* | 0.07723704 |
| *ADAMTSL2* | 0.07706392 |
| *MRM1* | 0.07703574 |
| *PRKG2* | 0.07700437 |
| *CCSAP* | 0.07690521 |
| *ZNF641* | 0.07686293 |
| *NUS1* | 0.07680861 |
| *IL11* | 0.07678522 |
| *ACYP1* | 0.07678366 |
| *PCDHB10* | 0.07675955 |
| *CNTNAP3B* | 0.0767163 |
| *VMO1* | 0.07659737 |
| *LRRC39* | 0.07656463 |
| *PRMT5* | 0.07632002 |
| *SMCO2* | 0.0761078 |
| *LMBRD2* | 0.07594598 |
| *NPRL2* | 0.07577636 |
| *GSG1L* | 0.07574024 |
| *KLHL15* | 0.07570489 |
| *CTSF* | 0.07557254 |
| *SUSD4* | 0.07543737 |
| *LOC149373* | 0.07516987 |
| *MPP6* | 0.07493379 |
| *PPP4C* | 0.07490633 |
| *THBS3* | 0.07411068 |
| *IL1RAP* | 0.07403386 |
| *SDHB* | 0.07377213 |
| *LRRTM1* | 0.07353788 |
| *GALNT3* | 0.07298301 |
| *GNPDA1* | 0.07294656 |
| *CD6* | 0.0729224 |
| *LYPLA1* | 0.07288737 |
| *TEX30* | 0.072885 |
| *GDF10* | 0.07271056 |
| *PGM1* | 0.07250191 |

| | |
|---|---|
| *HIST1H1A* | 0.07225529 |
| *TBC1D16* | 0.07197679 |
| *MYCL* | 0.07158184 |
| *FDPS* | 0.07138876 |
| *NLGN4Y* | 0.0710177 |
| *DHRS2* | 0.07067258 |
| *KRT18P55* | 0.07044716 |
| *PEG10* | 0.07027855 |
| *RCHY1* | 0.07001383 |
| *PKP2* | 0.0699484 |
| *PRPS1* | 0.06980933 |
| *LRTM2* | 0.06975564 |
| *TRIM71* | 0.06974706 |
| *NDUFV2* | 0.06946448 |
| *WIPF3* | 0.06943682 |
| *CNTNAP5* | 0.06936376 |
| *IFIT1* | 0.06922229 |
| *NSG1* | 0.06890576 |
| *DUSP23* | 0.06885081 |
| *FRMD3* | 0.06877587 |
| *PTPRM* | 0.06876034 |
| *TNS3* | 0.06874813 |
| *RERG* | 0.06860283 |
| *LDHB* | 0.06848308 |
| *C15orf59* | 0.06831708 |
| *GFM2* | 0.06830178 |
| *GABRE* | 0.06829685 |
| *TMEM196* | 0.06818055 |
| *ZNF280A* | 0.06811505 |
| *ANKS1B* | 0.06810595 |
| *STPG1* | 0.06806747 |
| *SF3A3* | 0.06789179 |
| *VSIG4* | 0.0678413 |
| *CLU* | 0.06774938 |
| *PDSS2* | 0.06772234 |
| *SLC35B1* | 0.0675505 |
| *GSTM1* | 0.06726281 |
| *MYLK4* | 0.06720842 |
| *SRP68* | 0.06719134 |
| *RIPOR2* | 0.06682965 |
| *C1QTNF12* | 0.06681604 |
| *CRYGD* | 0.06673477 |
| *CDKL5* | 0.06652888 |

| | |
|---|---|
| *RASSF5* | 0.06619793 |
| *CPLX3* | 0.06610324 |
| *TINCR* | 0.06606758 |
| *RALYL* | 0.06591855 |
| *LOC441204* | 0.06582628 |
| *ARHGAP36* | 0.06554578 |
| *CLPTM1* | 0.06535758 |
| *TPD52* | 0.06532498 |
| *XKR4* | 0.06511189 |
| *IPCEF1* | 0.06490092 |
| *CYTOR* | 0.06481734 |
| *HCG23* | 0.064688 |
| *NEK6* | 0.06445002 |
| *RGS14* | 0.06434173 |
| *FAM71F1* | 0.06432443 |
| *FBXO3* | 0.0642947 |
| *PPEF1* | 0.0641336 |
| *TRIM58* | 0.06402294 |
| *AP1S2* | 0.06394708 |
| *MFN2* | 0.06387719 |
| *NDUFA6.AS1* | 0.06386456 |
| *GRM1* | 0.06361203 |
| *MARCO* | 0.06349879 |
| *MRPS7* | 0.06322716 |
| *MARCH2* | 0.06308897 |
| *YY1P2* | 0.06308882 |
| *ZNF26* | 0.06279098 |
| *FAR2P1* | 0.06273698 |
| *NTNG1* | 0.06253379 |
| *LRRN1* | 0.06249298 |
| *PRKCD* | 0.06218286 |
| *UROS* | 0.06217407 |
| *MED19* | 0.06201659 |
| *KLHL1* | 0.06191313 |
| *C5orf49* | 0.06185245 |
| *LAMB1* | 0.06154093 |
| *FRAS1* | 0.06153153 |
| *SLC8A1* | 0.06129918 |
| *ITGA8* | 0.06080087 |
| *RADIL* | 0.06067512 |
| *OTUB2* | 0.06039484 |
| *ATP2A2* | 0.06031132 |
| *MBLAC2* | 0.06028049 |

| | |
|---|---|
| *LINC.PINT* | 0.06026056 |
| *PARL* | 0.06014996 |
| *RIMS3* | 0.05997632 |
| *GNG4* | 0.05988167 |
| *COL13A1* | 0.05984376 |
| *MRPS2* | 0.05975147 |
| *ADCYAP1* | 0.05969775 |
| *MAGEF1* | 0.05968095 |
| *BBS7* | 0.05949916 |
| *ATXN7L2* | 0.05926839 |
| *CCDC171* | 0.05914483 |
| *B3GALT1* | 0.05888455 |
| *CYCS* | 0.05885372 |
| *GDPD5* | 0.05856834 |
| *TIRAP* | 0.05847446 |
| *SLC17A6* | 0.05843505 |
| *GALNT14* | 0.05834809 |
| *ILDR2* | 0.05816857 |
| *NRAP* | 0.0580628 |
| *RPS6KA6* | 0.05802986 |
| *TSPAN14* | 0.05795732 |
| *ASIC2* | 0.05783411 |
| *SLC5A3* | 0.05781263 |
| *ADAR* | 0.0575149 |
| *CBLN1* | 0.05748137 |
| *NR3C2* | 0.05730052 |
| *CA8* | 0.05714326 |
| *SYTL5* | 0.05708973 |
| *ANKRD50* | 0.05706953 |
| *SULF1* | 0.05702236 |
| *CCDC68* | 0.05700161 |
| *STARD4* | 0.05685809 |
| *ARL15* | 0.05674573 |
| *GLI1* | 0.05654284 |
| *HUNK* | 0.05645456 |
| *DGKH* | 0.05626077 |
| *PSD3* | 0.0562291 |
| *FAM86HP* | 0.05621248 |
| *HENMT1* | 0.05614133 |
| *ATXN1* | 0.05612517 |
| *FHL2* | 0.05580889 |
| *TRABD2A* | 0.05564641 |
| *CALM2* | 0.0555763 |

| | |
|---|---|
| *VWDE* | 0.0555544 |
| *SELENOM* | 0.05532073 |
| *PTX3* | 0.05514412 |
| *B3GNTL1* | 0.05509772 |
| *RFK* | 0.05482306 |
| *HCN3* | 0.05477573 |
| *CACNA1G* | 0.0547309 |
| *DPP10.AS1* | 0.05467833 |
| *ACTA2* | 0.05467179 |
| *FAM220A* | 0.05450427 |
| *ATAD2* | 0.05445812 |
| *CEP112* | 0.05427719 |
| *ACTR3C* | 0.05427687 |
| *TMEM203* | 0.0541366 |
| *RAB40AL* | 0.05413125 |
| *LRRC7* | 0.05382263 |
| *STUM* | 0.0536636 |
| *PCDHB2* | 0.05352267 |
| *TMEM97* | 0.05351417 |
| *LKAAEAR1* | 0.05327364 |
| *TSPYL4* | 0.05326229 |
| *CKMT1B* | 0.0531281 |
| *KIAA0895* | 0.05310739 |
| *LINC02470* | 0.05305972 |
| *ACP2* | 0.05267872 |
| *THSD1* | 0.05252747 |
| *MELTF* | 0.05251242 |
| *TMEM215* | 0.0525114 |
| *PCDH18* | 0.05244785 |
| *ELL* | 0.05240284 |
| *FZD8* | 0.05228997 |
| *CAMK2N2* | 0.05219051 |
| *WNT5B* | 0.0521515 |
| *DNASE2* | 0.05213155 |
| *ZNF287* | 0.05207508 |
| *RPL27A* | 0.05206726 |
| *UST* | 0.05162527 |
| *CHERP* | 0.05158937 |
| *PIF1* | 0.05156862 |
| *PCYOX1L* | 0.05149375 |
| *TMEM164* | 0.05148768 |
| *SDR16C5* | 0.05136051 |
| *LOC642852* | 0.05131699 |

| | |
|---|---|
| *ACSL3* | 0.05127733 |
| *CPNE4* | 0.0512216 |
| *SUPT3H* | 0.05088755 |
| *TP53AIP1* | 0.05046589 |
| *DCC* | 0.05045064 |
| *RASD2* | 0.05036612 |
| *SDAD1* | 0.05034897 |
| *CYP4F35P* | 0.05014452 |
| *SIAH3* | 0.05011841 |
| *BAIAP3* | 0.04981704 |
| *CMBL* | 0.04979994 |
| *SCARF2* | 0.04955062 |
| *LRRIQ1* | 0.04925252 |
| *YTHDC2* | 0.04923893 |
| *HTR7* | 0.04911961 |
| *CLVS1* | 0.04890019 |
| *MRPS26* | 0.04882885 |
| *MGLL* | 0.0487943 |
| *COCH* | 0.04854036 |
| *RAB3A* | 0.04836819 |
| *ST6GALNAC5* | 0.04835185 |
| *DENND6B* | 0.04819064 |
| *TMEM185A* | 0.04814993 |
| *GSPT2* | 0.04790949 |
| *KCNH5* | 0.04775423 |
| *LAMTOR3* | 0.04730244 |
| *MEG8* | 0.04727431 |
| *ADORA2B* | 0.04719915 |
| *CPNE6* | 0.0471889 |
| *HERC2* | 0.04708288 |
| *OVGP1* | 0.04707173 |
| *TUBB4A* | 0.04701993 |
| *MRAP2* | 0.04680116 |
| *GRIN2B* | 0.04663214 |
| *MAP10* | 0.04649526 |
| *LOC285097* | 0.04635985 |
| *CDH11* | 0.04635847 |
| *PLCXD3* | 0.04632944 |
| *ULBP2* | 0.04632518 |
| *TMEM91* | 0.04626622 |
| *AKT1* | 0.04598201 |
| *CLEC4G* | 0.04595568 |
| *RASGRP2* | 0.04591564 |

| | |
|---|---|
| *EAF1* | 0.04590136 |
| *ARSJ* | 0.04588208 |
| *CIT* | 0.04583678 |
| *PRPS2* | 0.04579486 |
| *ERGIC1* | 0.04572409 |
| *LOC100128239* | 0.0457004 |
| *ETV1* | 0.04568299 |
| *COQ10A* | 0.04557916 |
| *LRP1B* | 0.04556928 |
| *TRAF7* | 0.04534432 |
| *PEG3* | 0.04534363 |
| *EPHA6* | 0.0452497 |
| *PTPN4* | 0.04502404 |
| *CPNE7* | 0.04501411 |
| *NUDT18* | 0.0447612 |
| *PPP2CA* | 0.04469361 |
| *GULP1* | 0.04457711 |
| *DYNC1LI1* | 0.04436181 |
| *PCDH1* | 0.04435784 |
| *NIPA2* | 0.04413606 |
| *ECT2* | 0.04408369 |
| *DHX58* | 0.04398357 |
| *MSANTD1* | 0.04385475 |
| *ANKRD33B* | 0.04383097 |
| *ZBTB18* | 0.04381798 |
| *GADD45A* | 0.04378902 |
| *STOML2* | 0.04369764 |
| *BHLHE40* | 0.04369164 |
| *STRIP2* | 0.04347317 |
| *RIMS2* | 0.04343558 |
| *RIOX1* | 0.04333336 |
| *SH2B2* | 0.04323449 |
| *FLYWCH1* | 0.04312928 |
| *KCNQ3* | 0.0429094 |
| *PTTG2* | 0.04276659 |
| *CHRNA6* | 0.0427503 |
| *NGB* | 0.0425083 |
| *POLR3H* | 0.04237638 |
| *TNK2* | 0.04224234 |
| *KCNAB1* | 0.04222102 |
| *MARC2* | 0.04220647 |
| *ATRNL1* | 0.04217699 |
| *NOL9* | 0.04210704 |

| | |
|---|---|
| *ADGRA1* | 0.04202266 |
| *PUS7* | 0.04193286 |
| *CAP2* | 0.0419195 |
| *EPHA7* | 0.04187639 |
| *HIPK1* | 0.04181393 |
| *CCDC6* | 0.04169355 |
| *ISPD* | 0.04166994 |
| *MSRB3* | 0.0416574 |
| *DDX10* | 0.04129803 |
| *ZNF385B* | 0.04119453 |
| *FOXG1* | 0.04116444 |
| *CDH4* | 0.04113444 |
| *RAB35* | 0.0411335 |
| *THAP4* | 0.04112687 |
| *NPAS2* | 0.04101258 |
| *CTSK* | 0.04096176 |
| *WASF1* | 0.04076454 |
| *HTR2C* | 0.04074083 |
| *TMEM248* | 0.04056753 |
| *KITLG* | 0.04053423 |
| *ZIC5* | 0.04050511 |
| *RAD54B* | 0.04039872 |
| *HOMER2* | 0.04037339 |
| *OLFM3* | 0.04029166 |
| *DPM2* | 0.03994231 |
| *GSTM3* | 0.03958106 |
| *SLF1* | 0.03957306 |
| *TPBG* | 0.03955412 |
| *RHOF* | 0.03937646 |
| *TCTE1* | 0.03933977 |
| *PCDHB11* | 0.03933641 |
| *MRC1* | 0.03933359 |
| *AQP11* | 0.03907991 |
| *WDR82* | 0.03877454 |
| *MEF2A* | 0.03875439 |
| *SLC17A5* | 0.03875256 |
| *HS6ST2* | 0.03875061 |
| *TMEM74* | 0.03870244 |
| *CBLN2* | 0.03850774 |
| *HDC* | 0.03845611 |
| *SPATA18* | 0.03817423 |
| *POMGNT2* | 0.03814777 |
| *COX11* | 0.03811608 |

| | |
|---|---|
| *SLC9A1* | 0.03796566 |
| *CACNA1C* | 0.03783185 |
| *RBP4* | 0.03781545 |
| *MRPL3* | 0.03779658 |
| *TLK1* | 0.03766401 |
| *B4GALT6* | 0.03750614 |
| *NDRG4* | 0.03727004 |
| *ZNF711* | 0.03726301 |
| *BAAT* | 0.0371207 |
| *FAM32A* | 0.03708407 |
| *FAM86B3P* | 0.03673189 |
| *EXTL3* | 0.03665281 |
| *GFRA2* | 0.03662568 |
| *TRPM4* | 0.03660007 |
| *CDKL4* | 0.03656357 |
| *ZWINT* | 0.03654963 |
| *ANXA6* | 0.03652036 |
| *FAM219B* | 0.03633712 |
| *SNN* | 0.03619873 |
| *STK38L* | 0.0360252 |
| *RETREG2* | 0.03597048 |
| *DHX57* | 0.03590719 |
| *AIFM3* | 0.03580199 |
| *ATP5F1A* | 0.03569435 |
| *C19orf66* | 0.03557847 |
| *XPNPEP1* | 0.03556018 |
| *PRRT4* | 0.0354285 |
| *ARNTL* | 0.03536854 |
| *FKBP11* | 0.03535048 |
| *GPD2* | 0.03512597 |
| *NAP1L3* | 0.03507794 |
| *ANKRD20A12P* | 0.03489431 |
| *C1orf109* | 0.03481339 |
| *TMEM9* | 0.03472978 |
| *CACNA2D1* | 0.03458545 |
| *CDK16* | 0.03447421 |
| *CCDC92* | 0.03445562 |
| *GLRA2* | 0.03443489 |
| *SHANK3* | 0.03429633 |
| *NUDT17* | 0.03419199 |
| *C9orf116* | 0.03401307 |
| *PRKCI* | 0.03397566 |
| *MEX3B* | 0.03390018 |

| | |
|---|---|
| *PPP2R1A* | 0.03389172 |
| *SYT7* | 0.03378474 |
| *PCNX2* | 0.03361888 |
| *PCDHB14* | 0.0335669 |
| *NEU1* | 0.03356659 |
| *KIAA0319* | 0.03334688 |
| *KCTD17* | 0.03331969 |
| *UNC119B* | 0.03327677 |
| *ADCY3* | 0.03320596 |
| *PBX4* | 0.03320034 |
| *CRH* | 0.03318084 |
| *ARPC4* | 0.03304559 |
| *NHS* | 0.03280035 |
| *USP43* | 0.03249389 |
| *ENPP5* | 0.03218974 |
| *ZNF749* | 0.03205355 |
| *ZDHHC8P1* | 0.03198632 |
| *BRINP3* | 0.03192224 |
| *ZNF483* | 0.03187195 |
| *TMEM177* | 0.03182149 |
| *TBPL1* | 0.03138892 |
| *PLEKHA6* | 0.03136164 |
| *ANKRD45* | 0.03121041 |
| *FAR2* | 0.031167 |
| *SLIT1* | 0.03053461 |
| *PARM1* | 0.03049715 |
| *NCK2* | 0.03046588 |
| *COL25A1* | 0.03018335 |
| *FEM1C* | 0.03012417 |
| *SPRN* | 0.02966625 |
| *ASB2* | 0.02963354 |
| *LYPD6* | 0.02942283 |
| *TENM3* | 0.02939571 |
| *PABPC1L2B* | 0.02902998 |
| *DUSP5* | 0.02895583 |
| *OPCML* | 0.02895127 |
| *EHD3* | 0.02891221 |
| *PLXNC1* | 0.02888834 |
| *GABBR2* | 0.02886195 |
| *CENPP* | 0.02880611 |
| *CCDC32* | 0.02867144 |
| *LSM11* | 0.02856823 |
| *ANKRD18DP* | 0.02854788 |

| | |
|---|---|
| *GOLT1A* | 0.02796871 |
| *GABRA5* | 0.02789787 |
| *TNFRSF11A* | 0.02788178 |
| *ADAT2* | 0.02780904 |
| *GPC5* | 0.02761008 |
| *ICMT* | 0.02740723 |
| *ZNF331* | 0.02737752 |
| *DENR* | 0.027376 |
| *PRDX2* | 0.02733604 |
| *SLC17A7* | 0.02719021 |
| *IQSEC2* | 0.02715187 |
| *CCDC65* | 0.02714489 |
| *IL1RAPL2* | 0.0270523 |
| *RIIAD1* | 0.02694968 |
| *CAMSAP1* | 0.02680917 |
| *STK32C* | 0.02666778 |
| *CCDC189* | 0.02666528 |
| *RHEB* | 0.02632411 |
| *ARL6* | 0.02630027 |
| *GALNT18* | 0.02612537 |
| *LOC105376805* | 0.02611944 |
| *SIN3B* | 0.02584161 |
| *UBR1* | 0.02579969 |
| *ANKRD20A8P* | 0.02574273 |
| *KALRN* | 0.02540804 |
| *DNAH5* | 0.02540219 |
| *MAL2* | 0.02521983 |
| *PIP5K1B* | 0.02520471 |
| *PDE3B* | 0.02501717 |
| *MPPED2* | 0.02489286 |
| *BAIAP2* | 0.02463158 |
| *PLAGL1* | 0.02462013 |
| *TDRD9* | 0.02458771 |
| *VILL* | 0.02455265 |
| *SYNPR* | 0.02440229 |
| *CALHM6* | 0.02438699 |
| *RBFOX1* | 0.02424272 |
| *NSMF* | 0.02418889 |
| *NIPSNAP1* | 0.02415615 |
| *BCL11B* | 0.02412915 |
| *TAF1B* | 0.02401909 |
| *SRC* | 0.0238721 |
| *HTR4* | 0.02376489 |

| | |
|---|---|
| *PTHLH* | 0.02373257 |
| *LGALS8* | 0.02361577 |
| *GDA* | 0.02358416 |
| *FSTL4* | 0.02345891 |
| *VDAC3* | 0.02344281 |
| *C1orf52* | 0.02341579 |
| *TUBA8* | 0.02336027 |
| *MRPS6* | 0.02314841 |
| *NRCAM* | 0.02302803 |
| *HSPA12A* | 0.02292393 |
| *RGS7BP* | 0.02289038 |
| *ARHGEF3* | 0.0226606 |
| *UBE2W* | 0.02264391 |
| *CFAP46* | 0.02253773 |
| *TRAPPC5* | 0.02253226 |
| *TRIM17* | 0.02234007 |
| *RAD18* | 0.02212006 |
| *CTXN2* | 0.02185266 |
| *PRDX5* | 0.02171665 |
| *CPLX2* | 0.02144237 |
| *NEK11* | 0.02134318 |
| *SYT10* | 0.02124895 |
| *CDK5R1* | 0.02109698 |
| *MTUS2* | 0.02098692 |
| *PNMA6E* | 0.02095095 |
| *CCNC* | 0.02081792 |
| *FAM95A* | 0.02056716 |
| *RNF6* | 0.02052405 |
| *LRRN2* | 0.02035935 |
| *ATP7B* | 0.02007084 |
| *GRM5* | 0.01980252 |
| *GALNT11* | 0.01972187 |
| *KLHL36* | 0.01962123 |
| *MATK* | 0.01958787 |
| *TPI1* | 0.01950352 |
| *COL23A1* | 0.01934375 |
| *SYNGAP1* | 0.01932742 |
| *C17orf67* | 0.01921394 |
| *PPP2R5B* | 0.01919977 |
| *C3orf33* | 0.01917107 |
| *CD24* | 0.01910666 |
| *FZD7* | 0.01897943 |
| *HIVEP2* | 0.01889324 |

| | |
|---|---|
| *C1orf53* | 0.01880195 |
| *PCTP* | 0.01880038 |
| *CNTN5* | 0.01854729 |
| *ST3GAL2* | 0.01840842 |
| *EPHA5.AS1* | 0.01837896 |
| *ATP2B1* | 0.0183604 |
| *OLFM2* | 0.01835781 |
| *GEMIN7* | 0.01822841 |
| *MPC2* | 0.01817777 |
| *TENM4* | 0.01817659 |
| *CAMK2A* | 0.01805452 |
| *MCF2L.AS1* | 0.01764574 |
| *IFT57* | 0.01763506 |
| *KIAA1671* | 0.01760919 |
| *MRPS10* | 0.01749659 |
| *MIR7.3HG* | 0.01735741 |
| *CALN1* | 0.01725079 |
| *MICU3* | 0.01715903 |
| *PREP* | 0.01713214 |
| *LGI1* | 0.01702995 |
| *SHISA9* | 0.01688344 |
| *ALDOC* | 0.0168252 |
| *HARS* | 0.01675051 |
| *SLC16A2* | 0.01671829 |
| *HTR7P1* | 0.01671375 |
| *WASL* | 0.01658662 |
| *PCDH8* | 0.01642156 |
| *SLC16A14* | 0.01640364 |
| *SYT3* | 0.01636885 |
| *PTPN3* | 0.01635789 |
| *PYCR3* | 0.01624505 |
| *TRPC4* | 0.01573413 |
| *RSPO3* | 0.01554962 |
| *ESYT3* | 0.01546728 |
| *TTC39C* | 0.01544571 |
| *DCAF6* | 0.01536824 |
| *EGR4* | 0.01514361 |
| *AKR1B1* | 0.01495038 |
| *RARRES1* | 0.01478815 |
| *FAM213A* | 0.01474204 |
| *ANKRD20A5P* | 0.01458204 |
| *MYH10* | 0.01452753 |
| *NCKAP1* | 0.01451637 |

| | |
|---|---|
| *SPATA20* | 0.01448049 |
| *FBXO2* | 0.01436826 |
| *NXPH1* | 0.01402519 |
| *NOS1AP* | 0.01380861 |
| *C8orf34* | 0.01354515 |
| *PLPPR5* | 0.01348343 |
| *ANKRD20A9P* | 0.01345766 |
| *COL12A1* | 0.01341116 |
| *SGSM1* | 0.01335742 |
| *LRP12* | 0.01332122 |
| *FADS6* | 0.01327732 |
| *GABRD* | 0.01322231 |
| *ZC3H12B* | 0.01317474 |
| *AP1S3* | 0.01313365 |
| *ANKDD1A* | 0.01300904 |
| *NDUFAF4* | 0.01292184 |
| *EPHA4* | 0.01276683 |
| *DOK6* | 0.0126805 |
| *CA7* | 0.01266024 |
| *DLGAP4* | 0.01263028 |
| *STK24* | 0.0123192 |
| *CENPVL3* | 0.0120015 |
| *SYNGR3* | 0.01192007 |
| *INSM2* | 0.01184572 |
| *SLC27A2* | 0.01176759 |
| *SNTG1* | 0.01162536 |
| *H2AFY* | 0.01157808 |
| *SLC35G2* | 0.0115137 |
| *UBE2B* | 0.0113871 |
| *PFKM* | 0.01138229 |
| *FAM189A1* | 0.01105322 |
| *ZNF91* | 0.0110342 |
| *SPINK2* | 0.01098886 |
| *PCSK1N* | 0.01097948 |
| *RTN4RL1* | 0.01092673 |
| *CLSTN2* | 0.01088713 |
| *GPR63* | 0.0104673 |
| *SRM* | 0.01037754 |
| *OCIAD2* | 0.01029574 |
| *GALNT17* | 0.01016257 |
| *PTGR1* | 0.01008546 |
| *TESK1* | 0.01005624 |
| *PITPNM1* | 0.01001769 |

| | |
|---|---|
| *IL27RA* | 0.00994027 |
| *DRC1* | 0.00987071 |
| *NMT2* | 0.00976087 |
| *LOR* | 0.00968223 |
| *PPP3CB* | 0.00956342 |
| *DGKI* | 0.0094902 |
| *TMEM158* | 0.00942457 |
| *TAC1* | 0.00940734 |
| *F12* | 0.00939449 |
| *KCNA4* | 0.00934011 |
| *CPE* | 0.00911649 |
| *GRK6* | 0.0090394 |
| *ANKRD20A4.ANKRD20A20P* | 0.00882208 |
| *PPP4R4* | 0.00881994 |
| *DACH2* | 0.00873518 |
| *TMEM163* | 0.008685 |
| *STXBP5L* | 0.00864526 |
| *IFT140* | 0.0086198 |
| *RRAS2* | 0.00838416 |
| *PACS1* | 0.00816205 |
| *TTC7B* | 0.00794132 |
| *HTR1A* | 0.00790449 |
| *CACNB2* | 0.00767777 |
| *PJA2* | 0.0074935 |
| *TMEM200A* | 0.00738192 |
| *ZNF554* | 0.00717664 |
| *SEZ6L* | 0.00708034 |
| *ICA1* | 0.00706967 |
| *GOLGA2P7* | 0.00682273 |
| *DBNDD1* | 0.00663055 |
| *EFCAB6* | 0.00650199 |
| *DDAH1* | 0.00644541 |
| *QPCTL* | 0.00644123 |
| *CLDN12* | 0.0064318 |
| *LOC728392* | 0.0063831 |
| *SLC35E1* | 0.00627704 |
| *ANKRD19P* | 0.00607059 |
| *FNDC9* | 0.00583067 |
| *ROCK2* | 0.00556437 |
| *SSTR2* | 0.00548189 |
| *SYT1* | 0.00545683 |
| *RASGRP1* | 0.00505863 |
| *GRIA1* | 0.00493313 |

| | |
|---|---|
| *CCDC181* | 0.00490741 |
| *NDST3* | 0.0047772 |
| *FEZF2* | 0.0047651 |
| *CLSTN3* | 0.00473516 |
| *FAM110B* | 0.00471817 |
| *C2CD2L* | 0.00464545 |
| *ZNF831* | 0.00393268 |
| *FRMPD4* | 0.0036095 |
| *CACNA1E* | 0.00309006 |
| *BRINP1* | 0.00299697 |
| *NEUROD2* | 0.00288623 |
| *NEUROD6* | 0.00283916 |
| *CCBE1* | 0.00275363 |
| *KIAA1257* | 0.00269343 |
| *SLC7A4* | 0.00266088 |
| *CCDC28A* | 0.00248023 |
| *OPRK1* | 0.00238699 |
| *CCSER1* | 0.00189661 |
| *ATP8A2* | 0.00146886 |
| *HSPH1* | 0.00137246 |
| *GRIPAP1* | 0.00117948 |
| *PHF14* | 0.00110864 |
| *CRMP1* | 0.0008285 |
| *ADGRL1* | 0.00077107 |
| *GABBR1* | 0.00066358 |
| *ADD2* | 0.00055813 |
| *CSRNP3* | 2.19484918752593e-05 |
| *GALNT16* | 1.47208659569047e-06 |
| *KCNIP4* | 1.41651422824335e-06 |
| *EEF2K* | 1.32097591513313e-06 |
| *NRIP3* | 9.54243154303381e-07 |
| *CCDC120* | 1.12079432680073e-08 |
| *SMARCA1* | 2.53867543284352e-09 |
| *CEP126* | 7.61958849363857e-10 |
| *PPM1J* | 1.44965134035425e-11 |
| *KCNJ6* | 1.23987366000897e-12 |
| *SCG2* | 1.0380149635618e-12 |
| *GPC1* | 5.14182623420643e-13 |
| *DLG3* | 4.36157975126982e-14 |
| *EVL* | 3.07725784055035e-15 |
| *ACTA1* | 2.22044604925031e-16 |
| *APLP2* | 2.22044604925031e-16 |
| *ARHGAP44* | 2.22044604925031e-16 |

| | |
|---|---|
| *ARHGEF17* | 2.22044604925031e-16 |
| *ATL1* | 2.22044604925031e-16 |
| *C11orf49* | 2.22044604925031e-16 |
| *CCNA1* | 2.22044604925031e-16 |
| *CDK14* | 2.22044604925031e-16 |
| *CDR2* | 2.22044604925031e-16 |
| *CEBPG* | 2.22044604925031e-16 |
| *CKMT1A* | 2.22044604925031e-16 |
| *CNIH2* | 2.22044604925031e-16 |
| *CPNE9* | 2.22044604925031e-16 |
| *CYFIP2* | 2.22044604925031e-16 |
| *DEAF1* | 2.22044604925031e-16 |
| *DLGAP3* | 2.22044604925031e-16 |
| *DOK4* | 2.22044604925031e-16 |
| *FAM135B* | 2.22044604925031e-16 |
| *FAM69B* | 2.22044604925031e-16 |
| *FMNL1* | 2.22044604925031e-16 |
| *FOCAD* | 2.22044604925031e-16 |
| *FRRS1L* | 2.22044604925031e-16 |
| *GABRB1* | 2.22044604925031e-16 |
| *GNB5* | 2.22044604925031e-16 |
| *GPI* | 2.22044604925031e-16 |
| *GPR68* | 2.22044604925031e-16 |
| *HMGCLL1* | 2.22044604925031e-16 |
| *IQCJ* | 2.22044604925031e-16 |
| *KCNB2* | 2.22044604925031e-16 |
| *L3MBTL2* | 2.22044604925031e-16 |
| *LMTK3* | 2.22044604925031e-16 |
| *LYSMD2* | 2.22044604925031e-16 |
| *MAP2* | 2.22044604925031e-16 |
| *MARK4* | 2.22044604925031e-16 |
| *ME3* | 2.22044604925031e-16 |
| *NAT14* | 2.22044604925031e-16 |
| *NCDN* | 2.22044604925031e-16 |
| *NEDD4L* | 2.22044604925031e-16 |
| *NEK10* | 2.22044604925031e-16 |
| *NEUROD1* | 2.22044604925031e-16 |
| *NPTN* | 2.22044604925031e-16 |
| *ORAI2* | 2.22044604925031e-16 |
| *PAIP1* | 2.22044604925031e-16 |
| *PANK2* | 2.22044604925031e-16 |
| *PELI3* | 2.22044604925031e-16 |
| *PIM2* | 2.22044604925031e-16 |

| | |
|---|---|
| *PPP1R9B* | 2.22044604925031e-16 |
| *PTK2B* | 2.22044604925031e-16 |
| *RAB15* | 2.22044604925031e-16 |
| *RASGEF1A* | 2.22044604925031e-16 |
| *RBFOX2* | 2.22044604925031e-16 |
| *RIMS1* | 2.22044604925031e-16 |
| *RND3* | 2.22044604925031e-16 |
| *RTN2* | 2.22044604925031e-16 |
| *SCN3A* | 2.22044604925031e-16 |
| *SCN3B* | 2.22044604925031e-16 |
| *SLC12A5* | 2.22044604925031e-16 |
| *SLC1A1* | 2.22044604925031e-16 |
| *SLC26A8* | 2.22044604925031e-16 |
| *SLC41A2* | 2.22044604925031e-16 |
| *SLC8A3* | 2.22044604925031e-16 |
| *SLC9A7* | 2.22044604925031e-16 |
| *SMPD3* | 2.22044604925031e-16 |
| *SNPH* | 2.22044604925031e-16 |
| *SPINT2* | 2.22044604925031e-16 |
| *SRXN1* | 2.22044604925031e-16 |
| *TBC1D30* | 2.22044604925031e-16 |
| *TSPYL2* | 2.22044604925031e-16 |
| *YWHAZ* | 2.22044604925031e-16 |

#### Supplementary Table 6: Limma linear model statistics for normative spatial expression of *C9orf72* and the top 100 transcripts that contribute to Component 7.

| Transcript | ß | t | p | Adjusted p |
|---|---|---|---|---|
| *DMBT1* | 0.070 | 3.44 | 5.95E-04 | 0.021 |
| *ARRDC2* | 0.069 | 3.40 | 6.68E-04 | 0.021 |
| *C9orf3* | 0.068 | 3.35 | 8.25E-04 | 0.021 |
| *RAB7B* | 0.065 | 3.20 | 1.40E-03 | 0.021 |
| *ZBED3* | 0.065 | 3.17 | 1.52E-03 | 0.021 |
| *KIAA1147* | 0.064 | 3.12 | 1.80E-03 | 0.021 |
| *GALNT6* | 0.063 | 3.10 | 1.94E-03 | 0.021 |
| *ENPP6* | 0.062 | 3.06 | 2.20E-03 | 0.021 |
| *JPT2* | 0.061 | 3.00 | 2.74E-03 | 0.021 |
| *FAM149B1* | 0.060 | 2.96 | 3.04E-03 | 0.021 |
| *PCBP4* | 0.060 | 2.96 | 3.09E-03 | 0.021 |
| *LOC101929384* | 0.059 | 2.92 | 3.55E-03 | 0.021 |
| *SLC31A2* | 0.059 | 2.91 | 3.68E-03 | 0.021 |
| *TSPAN15* | 0.059 | 2.90 | 3.70E-03 | 0.021 |
| *SLC5A11* | 0.059 | 2.89 | 3.92E-03 | 0.021 |
| *TSPAN8* | 0.058 | 2.86 | 4.26E-03 | 0.021 |
| *ADAMTS4* | 0.058 | 2.84 | 4.48E-03 | 0.021 |
| *PHF10* | 0.058 | 2.84 | 4.57E-03 | 0.021 |
| *TMEM98* | 0.057 | 2.82 | 4.85E-03 | 0.021 |
| *KHNYN* | 0.057 | 2.81 | 4.94E-03 | 0.021 |
| *USP54* | 0.057 | 2.80 | 5.15E-03 | 0.021 |
| *PCSK6* | 0.057 | 2.79 | 5.34E-03 | 0.021 |
| *TMTC2* | 0.057 | 2.78 | 5.47E-03 | 0.021 |
| *CARNS1* | 0.057 | 2.78 | 5.48E-03 | 0.021 |
| *NDE1* | 0.056 | 2.74 | 6.08E-03 | 0.021 |
| *CLDND1* | 0.056 | 2.74 | 6.11E-03 | 0.021 |
| *FEZ1* | 0.056 | 2.74 | 6.17E-03 | 0.021 |
| *PRKCQ* | 0.056 | 2.73 | 6.30E-03 | 0.021 |
| *ARHGAP21* | 0.055 | 2.72 | 6.64E-03 | 0.021 |
| *ASPRV1* | 0.055 | 2.71 | 6.72E-03 | 0.021 |
| *MAG* | 0.055 | 2.71 | 6.77E-03 | 0.021 |
| *HSD17B3* | 0.055 | 2.70 | 7.07E-03 | 0.021 |
| *ANKRD65* | 0.054 | 2.65 | 8.09E-03 | 0.021 |
| *LDLRAD4* | 0.054 | 2.64 | 8.26E-03 | 0.021 |
| *LLGL2* | 0.054 | 2.64 | 8.42E-03 | 0.021 |
| *TMEM140* | 0.053 | 2.62 | 8.79E-03 | 0.021 |
| *IFIT2* | 0.053 | 2.62 | 8.91E-03 | 0.021 |

| | | | | |
|---|---|---|---|---|
| *SMARCC1* | 0.053 | 2.62 | 8.94E-03 | 0.021 |
| *USH1C* | 0.053 | 2.60 | 9.27E-03 | 0.021 |
| *ABCA8* | 0.053 | 2.59 | 9.55E-03 | 0.021 |
| *BAMBI* | 0.053 | 2.59 | 9.59E-03 | 0.021 |
| *FLNC* | 0.053 | 2.58 | 9.82E-03 | 0.021 |
| *CTNNA3* | 0.052 | 2.58 | 9.99E-03 | 0.021 |
| *KLC1* | 0.052 | 2.58 | 1.00E-02 | 0.021 |
| *TCFL5* | 0.052 | 2.58 | 1.00E-02 | 0.021 |
| *LOC100506725* | 0.052 | 2.56 | 1.04E-02 | 0.021 |
| *NUDT1* | 0.052 | 2.56 | 1.05E-02 | 0.021 |
| *ADAMTS14* | 0.052 | 2.56 | 1.06E-02 | 0.021 |
| *ADGRE5* | 0.052 | 2.56 | 1.06E-02 | 0.021 |
| *ZNF664* | 0.052 | 2.56 | 1.06E-02 | 0.021 |
| *SELENOP* | 0.052 | 2.55 | 1.08E-02 | 0.021 |
| *SFR1* | 0.052 | 2.54 | 1.12E-02 | 0.022 |
| *VKORC1* | 0.051 | 2.53 | 1.16E-02 | 0.022 |
| *PARP4* | 0.051 | 2.52 | 1.16E-02 | 0.022 |
| *LRP6* | 0.051 | 2.50 | 1.24E-02 | 0.023 |
| *LOC100507487* | 0.051 | 2.49 | 1.27E-02 | 0.023 |
| *ARHGEF37* | 0.050 | 2.47 | 1.37E-02 | 0.024 |
| *SAP30* | 0.050 | 2.46 | 1.38E-02 | 0.024 |
| *MYLK* | 0.050 | 2.46 | 1.40E-02 | 0.024 |
| *CTDSP1* | 0.050 | 2.45 | 1.45E-02 | 0.024 |
| *BCDIN3D* | 0.049 | 2.39 | 1.67E-02 | 0.027 |
| *CAPN3* | 0.049 | 2.39 | 1.68E-02 | 0.027 |
| *TNS2* | 0.048 | 2.37 | 1.80E-02 | 0.029 |
| *ADARB2.AS1* | 0.048 | 2.36 | 1.84E-02 | 0.029 |
| *B3GNT9* | 0.048 | 2.35 | 1.87E-02 | 0.029 |
| *GPRC5B* | 0.048 | 2.34 | 1.91E-02 | 0.029 |
| *SSFA2* | 0.047 | 2.33 | 1.98E-02 | 0.030 |
| *RASSF2* | 0.047 | 2.32 | 2.04E-02 | 0.030 |
| *HHATL* | 0.047 | 2.32 | 2.05E-02 | 0.030 |
| *FNTA* | 0.047 | 2.31 | 2.12E-02 | 0.031 |
| *CMTM5* | 0.045 | 2.21 | 2.71E-02 | 0.039 |
| *TMTC4* | 0.044 | 2.18 | 2.96E-02 | 0.042 |
| *SCRG1* | 0.044 | 2.16 | 3.06E-02 | 0.042 |
| *CRTAP* | 0.043 | 2.13 | 3.30E-02 | 0.045 |
| *CHST6* | 0.043 | 2.12 | 3.37E-02 | 0.045 |
| *QDPR* | 0.042 | 2.08 | 3.75E-02 | 0.0498 |
| *HIPK2* | 0.042 | 2.07 | 3.83E-02 | 0.0501 |
| *RDH11* | 0.042 | 2.07 | 3.87E-02 | 0.050 |
| *IGFLR1* | 0.041 | 2.00 | 4.58E-02 | 0.059 |
| *PAIP2B* | 0.040 | 1.99 | 4.69E-02 | 0.059 |

| | | | | |
|---|---|---|---|---|
| *MOBP* | 0.040 | 1.98 | 4.80E-02 | 0.060 |
| *S100B* | 0.039 | 1.90 | 5.81E-02 | 0.072 |
| *ARHGEF1* | 0.038 | 1.88 | 5.99E-02 | 0.073 |
| *PLEKHB1* | 0.037 | 1.84 | 6.65E-02 | 0.080 |
| *FRMD5* | 0.037 | 1.82 | 6.95E-02 | 0.083 |
| *CDH20* | 0.037 | 1.80 | 7.17E-02 | 0.084 |
| *PPIF* | 0.036 | 1.78 | 7.50E-02 | 0.087 |
| *AAMDC* | 0.036 | 1.78 | 7.56E-02 | 0.087 |
| *KATNA1* | 0.035 | 1.73 | 8.31E-02 | 0.094 |
| *SEPT4* | 0.035 | 1.72 | 8.64E-02 | 0.097 |
| *FAT1* | 0.035 | 1.71 | 8.72E-02 | 0.097 |
| *TST* | 0.032 | 1.58 | 1.15E-01 | 0.126 |
| *ICK* | 0.030 | 1.46 | 1.45E-01 | 0.158 |
| *BCAR3* | 0.029 | 1.43 | 1.53E-01 | 0.164 |
| *DENND5A* | 0.029 | 1.41 | 1.60E-01 | 0.170 |
| *LAP3* | 0.028 | 1.35 | 1.76E-01 | 0.185 |
| *RGS1* | 0.020 | 0.99 | 3.21E-01 | 0.334 |
| *SESTD1* | 0.010 | 0.50 | 6.20E-01 | 0.639 |
| *FAM19A4* | 0.009 | 0.43 | 6.69E-01 | 0.682 |
| *C9orf72* | 0.007 | 0.36 | 7.18E-01 | 0.725 |
| *RCOR2* | 0.005 | 0.26 | 7.93E-01 | 0.793 |

#### Supplementary Table 7: Spearman-rank correlations of *C9orf72* and the top 100 transcripts in Component 7.

| Transcript | rho | *p* | Adjusted *p* |
|---|---|---|---|
| *C9orf72* | 1 | 0 | 0 |
| *AAMDC* | 0.68 | 1.13E-06 | 5.68E-05 |
| *GPRC5B* | 0.66 | 2.29E-06 | 7.71E-05 |
| *PPIF* | 0.65 | 4.00E-06 | 1.01E-04 |
| *TST* | 0.61 | 1.49E-05 | 3.01E-04 |
| *BCDIN3D* | 0.58 | 5.32E-05 | 8.96E-04 |
| *LDLRAD4* | 0.57 | 7.22E-05 | 1.04E-03 |
| *SELENOP* | 0.57 | 8.47E-05 | 1.07E-03 |
| *KLC1* | 0.55 | 1.53E-04 | 1.55E-03 |
| *FNTA* | 0.55 | 1.54E-04 | 1.55E-03 |
| *CMTM5* | 0.54 | 1.69E-04 | 1.55E-03 |
| *TMEM98* | 0.54 | 1.92E-04 | 1.62E-03 |
| *LOC100506725* | 0.53 | 2.32E-04 | 1.72E-03 |
| *RASSF2* | 0.53 | 2.58E-04 | 1.72E-03 |
| *CAPN3* | 0.53 | 2.65E-04 | 1.72E-03 |

| | | | |
|---|---|---|---|
| *USP54* | 0.53 | 2.87E-04 | 1.72E-03 |
| *CARNS1* | 0.53 | 2.90E-04 | 1.72E-03 |
| *MYLK* | 0.53 | 3.06E-04 | 1.72E-03 |
| *IFIT2* | 0.51 | 4.29E-04 | 2.28E-03 |
| *SLC31A2* | 0.51 | 4.95E-04 | 2.50E-03 |
| *TNS2* | 0.50 | 5.66E-04 | 2.72E-03 |
| *ARHGEF37* | 0.50 | 6.35E-04 | 2.83E-03 |
| *NDE1* | 0.50 | 6.79E-04 | 2.83E-03 |
| *CTNNA3* | 0.50 | 7.01E-04 | 2.83E-03 |
| *MAG* | 0.50 | 7.01E-04 | 2.83E-03 |
| *CLDND1* | 0.49 | 8.29E-04 | 3.22E-03 |
| *ADAMTS4* | 0.48 | 1.01E-03 | 3.55E-03 |
| *PLEKHB1* | 0.48 | 1.01E-03 | 3.55E-03 |
| *TCFL5* | 0.48 | 1.02E-03 | 3.55E-03 |
| *ASPRV1* | 0.48 | 1.08E-03 | 3.63E-03 |
| *TSPAN8* | 0.48 | 1.12E-03 | 3.66E-03 |
| *SEPT4* | 0.48 | 1.22E-03 | 3.84E-03 |
| *DENND5A* | 0.46 | 1.87E-03 | 5.53E-03 |
| *SLC5A11* | 0.46 | 1.87E-03 | 5.53E-03 |
| *PCBP4* | 0.46 | 1.91E-03 | 5.53E-03 |
| *KHNYN* | 0.46 | 2.08E-03 | 5.83E-03 |
| *GALNT6* | 0.45 | 2.45E-03 | 6.68E-03 |
| *VKORC1* | 0.45 | 2.51E-03 | 6.68E-03 |
| *TMEM140* | 0.45 | 2.61E-03 | 6.76E-03 |
| *MOBP* | 0.44 | 2.71E-03 | 6.76E-03 |
| *DMBT1* | 0.44 | 2.78E-03 | 6.76E-03 |
| *HIPK2* | 0.44 | 2.81E-03 | 6.76E-03 |
| *IGFLR1* | 0.44 | 3.03E-03 | 7.12E-03 |
| *SAP30* | 0.43 | 3.77E-03 | 8.66E-03 |
| *SCRG1* | 0.43 | 3.92E-03 | 8.80E-03 |
| *FRMD5* | 0.43 | 4.03E-03 | 8.84E-03 |
| *BCAR3* | 0.43 | 4.13E-03 | 8.88E-03 |
| *PARP4* | 0.42 | 4.39E-03 | 9.11E-03 |
| *ZNF664* | 0.42 | 4.42E-03 | 9.11E-03 |
| *PCSK6* | 0.42 | 4.74E-03 | 9.58E-03 |
| *HHATL* | 0.42 | 4.90E-03 | 9.70E-03 |
| *FAM149B1* | 0.42 | 5.28E-03 | 1.03E-02 |
| *JPT2* | 0.41 | 5.86E-03 | 1.12E-02 |
| *ADGRE5* | 0.41 | 6.41E-03 | 1.20E-02 |
| *FEZ1* | 0.41 | 6.52E-03 | 1.20E-02 |
| *USH1C* | 0.40 | 6.93E-03 | 1.25E-02 |
| *BAMBI* | 0.40 | 7.50E-03 | 1.33E-02 |
| *QDPR* | 0.39 | 8.78E-03 | 1.53E-02 |

| | | | |
|---|---|---|---|
| *CTDSP1* | 0.39 | 9.48E-03 | 1.62E-02 |
| *S100B* | 0.38 | 1.06E-02 | 1.79E-02 |
| *HSD17B3* | 0.38 | 1.13E-02 | 1.88E-02 |
| *ADARB2.AS1* | 0.38 | 1.22E-02 | 1.99E-02 |
| *ARRDC2* | 0.37 | 1.30E-02 | 2.07E-02 |
| *SFR1* | 0.37 | 1.31E-02 | 2.07E-02 |
| *B3GNT9* | 0.36 | 1.82E-02 | 2.78E-02 |
| *RAB7B* | 0.36 | 1.82E-02 | 2.78E-02 |
| *CHST6* | 0.35 | 1.92E-02 | 2.90E-02 |
| *ICK* | 0.35 | 1.99E-02 | 2.96E-02 |
| *LLGL2* | 0.32 | 3.37E-02 | 4.94E-02 |
| *TMTC2* | 0.32 | 3.46E-02 | 5.00E-02 |
| *PRKCQ* | 0.31 | 3.92E-02 | 5.58E-02 |
| *FAM19A4* | 0.31 | 4.27E-02 | 5.99E-02 |
| *PAIP2B* | 0.30 | 4.50E-02 | 6.23E-02 |
| *LAP3* | 0.30 | 4.87E-02 | 6.64E-02 |
| *ENPP6* | 0.29 | 5.24E-02 | 7.06E-02 |
| *SSFA2* | 0.29 | 5.85E-02 | 7.78E-02 |
| *ANKRD65* | 0.27 | 8.10E-02 | 1.06E-01 |
| *ZBED3* | 0.26 | 8.35E-02 | 1.08E-01 |
| *NUDT1* | -0.25 | 1.04E-01 | 1.33E-01 |
| *ABCA8* | 0.25 | 1.06E-01 | 1.33E-01 |
| *FLNC* | 0.24 | 1.23E-01 | 1.53E-01 |
| *FAT1* | 0.22 | 1.52E-01 | 1.87E-01 |
| *CRTAP* | 0.21 | 1.70E-01 | 2.06E-01 |
| *RDH11* | 0.21 | 1.79E-01 | 2.15E-01 |
| *KATNA1* | 0.20 | 1.82E-01 | 2.16E-01 |
| *SESTD1* | 0.18 | 2.41E-01 | 2.83E-01 |
| *TMTC4* | 0.17 | 2.59E-01 | 3.01E-01 |
| *LOC101929384* | 0.17 | 2.82E-01 | 3.23E-01 |
| *CDH20* | 0.16 | 2.96E-01 | 3.35E-01 |
| *LOC100507487* | 0.16 | 3.09E-01 | 3.47E-01 |
| *KIAA1147* | 0.14 | 3.69E-01 | 4.09E-01 |
| *SMARCC1* | 0.13 | 3.82E-01 | 4.19E-01 |
| *C9orf3* | 0.10 | 5.16E-01 | 5.61E-01 |
| *ARHGAP21* | -0.09 | 5.48E-01 | 5.89E-01 |
| *LRP6* | -0.08 | 5.93E-01 | 6.30E-01 |
| *PHF10* | 0.08 | 5.99E-01 | 6.30E-01 |
| *ARHGEF1* | 0.08 | 6.21E-01 | 6.47E-01 |
| *RGS1* | 0.06 | 6.90E-01 | 7.12E-01 |
| *TSPAN15* | 0.03 | 8.24E-01 | 8.41E-01 |
| *RCOR2* | 0.03 | 8.71E-01 | 8.80E-01 |
| *ADAMTS14* | -0.01 | 9.45E-01 | 9.45E-01 |

### Supplementary Table 8: Limma linear model statistics for normative spatial expression of the FTD- and neurodevelopment-related transcripts of interest in the six AHBA donors.

| Transcript | *ß* | *t* | *p* | Adjusted *p* |
|---|---|---|---|---|
| *CALB1* | -0.063 | -3.64 | 5.90E-04 | 0.022 |
| *NES* | 0.045 | 2.62 | 0.011 | 0.213 |
| *SQSTM1* | 0.039 | 2.11 | 0.039 | 0.497 |
| *NLGN4X* | -0.032 | -1.89 | 0.064 | 0.604 |
| *GAD1* | -0.035 | -1.74 | 0.087 | 0.664 |
| *HES1* | 0.022 | 1.50 | 0.138 | 0.767 |
| *S100A6* | 0.033 | 1.47 | 0.148 | 0.767 |
| *CHMP2B* | 0.020 | 1.37 | 0.177 | 0.767 |
| *TIA1* | 0.021 | 1.30 | 0.198 | 0.767 |
| *SLC32A1* | -0.028 | -1.26 | 0.214 | 0.767 |
| *SLC24A5* | 0.018 | 1.17 | 0.249 | 0.767 |
| *NRGN* | -0.019 | -1.12 | 0.269 | 0.767 |
| *EWSR1* | 0.017 | 1.07 | 0.288 | 0.767 |
| *OPTN* | 0.017 | 0.99 | 0.324 | 0.767 |
| *TCF7L2* | -0.021 | -0.98 | 0.333 | 0.767 |
| *PVALB* | 0.016 | 0.90 | 0.371 | 0.767 |
| *FUS* | 0.014 | 0.90 | 0.373 | 0.767 |
| *MET* | 0.011 | 0.81 | 0.421 | 0.767 |
| *MAPT* | -0.012 | -0.79 | 0.433 | 0.767 |
| *TMEM106B* | -0.011 | -0.77 | 0.443 | 0.767 |
| *CHCHD10* | -0.014 | -0.76 | 0.449 | 0.767 |
| *TBK1* | -0.016 | -0.74 | 0.464 | 0.767 |
| *SLC17A6* | -0.016 | -0.68 | 0.501 | 0.767 |
| *GRN* | 0.009 | 0.68 | 0.502 | 0.767 |
| *LHX2* | -0.013 | -0.67 | 0.505 | 0.767 |
| *SOX2* | 0.011 | 0.64 | 0.526 | 0.768 |
| *DLX1* | 0.007 | 0.49 | 0.626 | 0.838 |
| *C9orf72* | 0.007 | 0.48 | 0.635 | 0.838 |
| *CALB2* | -0.009 | -0.47 | 0.639 | 0.838 |
| *TAF15* | 0.004 | 0.22 | 0.823 | 0.990 |
| *TARDBP* | 0.003 | 0.19 | 0.850 | 0.990 |
| *CNTNAP2* | -0.002 | -0.15 | 0.883 | 0.990 |
| *FOXP2* | 0.003 | 0.15 | 0.885 | 0.990 |
| *CEACAM21* | 0.002 | 0.10 | 0.923 | 0.990 |
| *VCP* | -0.001 | -0.09 | 0.933 | 0.990 |
| *RGS16* | -0.001 | -0.05 | 0.957 | 0.990 |
| *CCNF* | 0.001 | 0.04 | 0.972 | 0.990 |

| | | | | |
|---|---|---|---|---|
| *DLX2* | 1.43E-04 | 0.01 | 0.990 | 0.990 |

#### Supplementary Table 9: Spearman-rank correlations between *C9orf72* and the FTD- and neurodevelopment-related transcripts of interest in the six AHBA donors.

| Transcript | rho | *p* | Adjusted *p* |
|---|---|---|---|
| *C9orf72* | 1 | 0 | 0 |
| *DLX2* | -0.639 | 5.33E-06 | 1.01E-04 |
| *NES* | 0.611 | 1.61E-05 | 2.04E-04 |
| *DLX1* | -0.602 | 2.32E-05 | 2.21E-04 |
| *SQSTM1* | 0.583 | 4.60E-05 | 3.50E-04 |
| *FOXP2* | 0.570 | 7.33E-05 | 4.64E-04 |
| *MAPT* | 0.564 | 8.97E-05 | 4.87E-04 |
| *FUS* | 0.523 | 3.28E-04 | 1.56E-03 |
| *CNTNAP2* | 0.488 | 8.92E-04 | 3.77E-03 |
| *TCF7L2* | 0.470 | 1.46E-03 | 5.24E-03 |
| *PVALB* | 0.468 | 1.52E-03 | 5.24E-03 |
| *RGS16* | 0.460 | 1.85E-03 | 5.82E-03 |
| *CHCHD10* | 0.458 | 1.99E-03 | 5.82E-03 |
| *CHMP2B* | 0.445 | 2.72E-03 | 7.38E-03 |
| *SLC17A6* | 0.418 | 5.07E-03 | 1.28E-02 |
| *SOX2* | 0.389 | 9.45E-03 | 2.24E-02 |
| *TAF15* | 0.385 | 1.03E-02 | 2.29E-02 |
| *CCNF* | 0.370 | 1.40E-02 | 2.95E-02 |
| *MET* | -0.361 | 1.67E-02 | 3.28E-02 |
| *LHX2* | -0.359 | 1.72E-02 | 3.28E-02 |
| *CALB2* | 0.356 | 1.83E-02 | 3.30E-02 |
| *S100A6* | 0.353 | 1.91E-02 | 3.30E-02 |
| *TARDBP* | 0.321 | 3.41E-02 | 5.64E-02 |
| *TBK1* | 0.293 | 5.37E-02 | 8.50E-02 |
| *NLGN4X* | -0.282 | 6.35E-02 | 9.66E-02 |
| *SLC24A5* | 0.278 | 6.84E-02 | 1.00E-01 |
| *CEACAM21* | -0.268 | 7.85E-02 | 1.11E-01 |
| *TIA1* | -0.264 | 8.35E-02 | 1.13E-01 |
| *OPTN* | 0.251 | 1.01E-01 | 1.32E-01 |
| *NRGN* | -0.211 | 1.69E-01 | 2.14E-01 |
| *TMEM106B* | -0.208 | 1.75E-01 | 2.15E-01 |
| *HES1* | 0.173 | 2.62E-01 | 3.11E-01 |
| *CALB1* | 0.109 | 4.81E-01 | 5.54E-01 |
| *SLC32A1* | 0.094 | 5.43E-01 | 6.01E-01 |

| | | | |
|---|---|---|---|
| *GRN* | -0.091 | 5.54E-01 | 6.01E-01 |
| *VCP* | 0.072 | 6.41E-01 | 6.77E-01 |
| *EWSR1* | -0.067 | 6.67E-01 | 6.85E-01 |
| *GAD1* | -0.032 | 8.38E-01 | 8.38E-01 |

### Exploratory Gene Ontology (GO) Analyses for Data-Driven Transcripts

GO analysis of the top 100 data-driven transcripts revealed 144 terms (108 biological processes, 20 cellular components, and 16 molecular functions), though they did not survive Benjamini-Hochberg correction at $p<0.05$ and $q<0.20$. The top 25 biological process terms included 14 developmental, cell growth, and cell cycle processes **(Suppl. Fig. 1a)**. Some developmental terms included positive regulation of stem cell population maintenance ($p$=1.57e-3), cellular component assembly involved in morphogenesis ($p$=4.26e-3), cellular anatomical entity morphogenesis ($p$=4.26e-3), myelin assembly ($p$=5.61e-3), transforming growth factor beta receptor signaling pathway ($p$=6.44e-3), central nervous system myelination ($p$=7.07e-3), and axon ensheathment in central nervous system ($p$=7.07e-3). GO analysis with *C9orf72* and the 70 transcripts with which positively associations occurred consistently revealed 175 terms, many developmental terms in the top 25, though these also did not survive correction **(Suppl. Fig. 1b)**. See Supplementary Tables 7-8 for the full lists of GO terms, respectively.

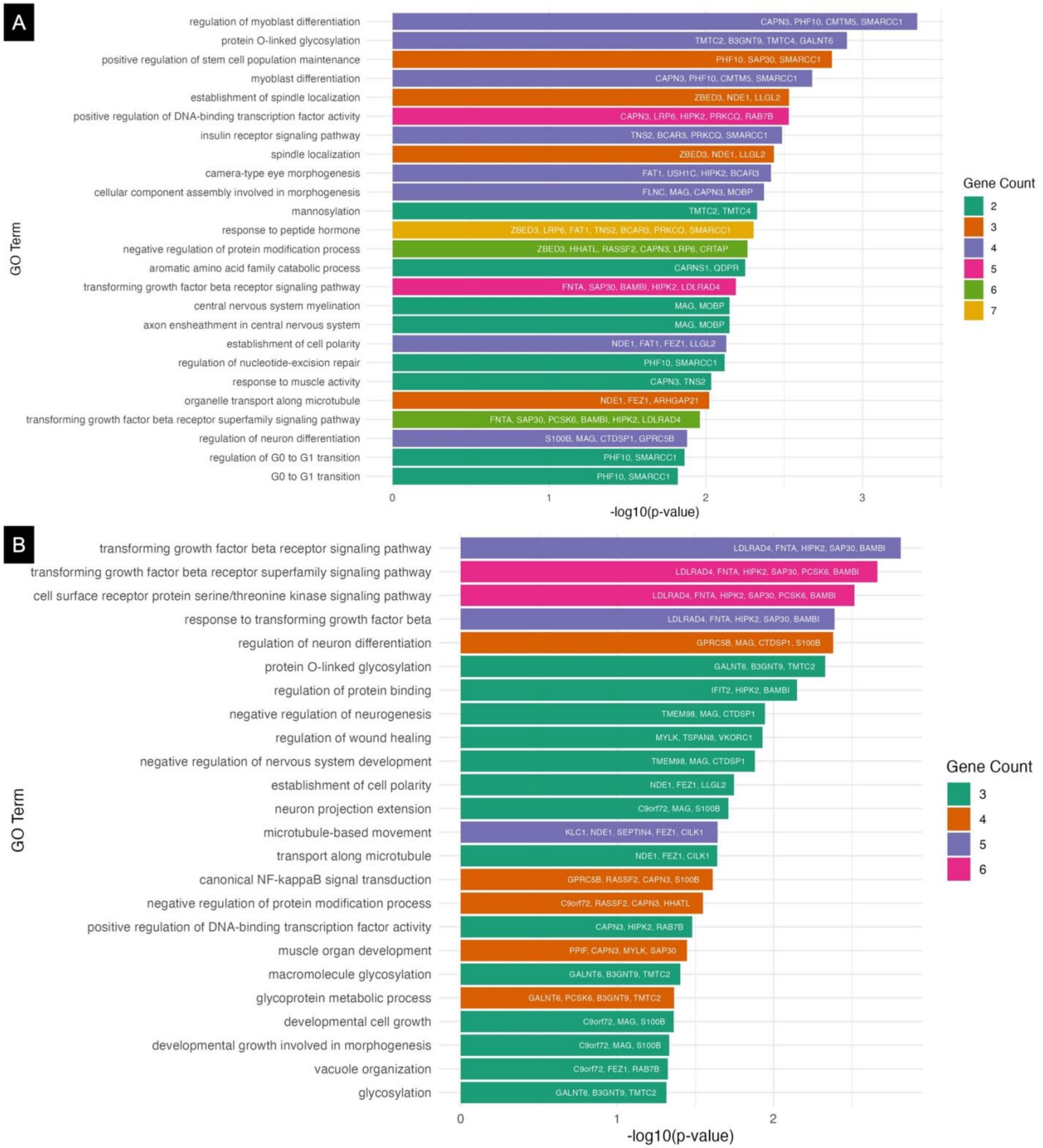

**Supplementary Figure 1. Exploratory gene ontology (GO) analyses of (A) the top 100 data-driven transcripts and (B) transcripts which also correlated significantly with *C9orf72* expression.** GO terms were primarily related to neurodevelopment, cell growth, and cell cycle maintenance. The top 25 biological process GO terms were significant at $p<0.05$ but did not survive correction for multiple comparisons. Where two highly similar terms were present, as determined using simplify() from clusterProfiler, the higher-ranked of the two was plotted. To facilitate understanding of potential relationships of the 70 transcripts with *C9orf72*, GO terms which involved three or more genes were plotted. The bar graph colours (gene counts) represent the number of genes that contributed to each GO term.

Supplementary Table 10: Exploratory gene ontology analysis of the top 100 gene transcripts that contribute to Component 7.

| Ontology | ID | Description | *p* | *q* | Contributory Genes | Gene Count |
|---|---|---|---|---|---|---|
| BP | GO:0045661 | regulation of myoblast differentiation | 4.47E-04 | 0.343 | CAPN3/PHF10/CMTM5/SMARCC1 | 4 |
| BP | GO:0006493 | protein O-linked glycosylation | 1.26E-03 | 0.343 | TMTC2/B3GNT9/TMTC4/GALNT6 | 4 |
| BP | GO:1902459 | positive regulation of stem cell population maintenance | 1.57E-03 | 0.343 | PHF10/SAP30/SMARCC1 | 3 |
| BP | GO:0045445 | myoblast differentiation | 2.10E-03 | 0.343 | CAPN3/PHF10/CMTM5/SMARCC1 | 4 |
| BP | GO:0051293 | establishment of spindle localization | 2.94E-03 | 0.343 | ZBED3/NDE1/LLGL2 | 3 |
| BP | GO:0051091 | positive regulation of DNA-binding transcription factor activity | 2.96E-03 | 0.343 | CAPN3/LRP6/HIPK2/PRKCQ/RAB7B | 5 |
| BP | GO:0008286 | insulin receptor signaling pathway | 3.27E-03 | 0.343 | TNS2/BCAR3/PRKCQ/SMARCC1 | 4 |
| BP | GO:0035269 | protein O-linked mannosylation | 3.53E-03 | 0.343 | TMTC2/TMTC4 | 2 |
| BP | GO:0051653 | spindle localization | 3.68E-03 | 0.343 | ZBED3/NDE1/LLGL2 | 3 |
| BP | GO:0048593 | camera-type eye morphogenesis | 3.84E-03 | 0.343 | FAT1/USH1C/HIPK2/BCAR3 | 4 |
| BP | GO:0010927 | cellular component assembly involved in morphogenesis | 4.26E-03 | 0.343 | FLNC/MAG/CAPN3/MOBP | 4 |
| BP | GO:0032989 | cellular anatomical entity morphogenesis | 4.26E-03 | 0.343 | FLNC/MAG/CAPN3/MOBP | 4 |
| BP | GO:0035268 | protein mannosylation | 4.31E-03 | 0.343 | TMTC2/TMTC4 | 2 |
| BP | GO:0002089 | lens morphogenesis in camera-type eye | 4.72E-03 | 0.343 | HIPK2/BCAR3 | 2 |
| BP | GO:0097502 | mannosylation | 4.72E-03 | 0.343 | TMTC2/TMTC4 | 2 |
| BP | GO:0043434 | response to peptide hormone | 4.96E-03 | 0.343 | ZBED3/LRP6/FAT1/TNS2/BCAR3/PRKCQ/SMARCC1 | 7 |
| BP | GO:2000036 | regulation of stem cell population maintenance | 5.08E-03 | 0.343 | PHF10/SAP30/SMARCC1 | 3 |
| BP | GO:0031400 | negative regulation of protein modification process | 5.43E-03 | 0.343 | ZBED3/HHATL/RASSF2/CAPN3/LRP6/CRTAP | 6 |
| BP | GO:0009074 | aromatic amino acid family catabolic process | 5.61E-03 | 0.343 | CARNS1/QDPR | 2 |
| BP | GO:0032288 | myelin assembly | 5.61E-03 | 0.343 | MAG/MOBP | 2 |

| | | | | | | |
|---|---|---|---|---|---|---|
| BP | GO:0007179 | transforming growth factor beta receptor signaling pathway | 6.44E-03 | 0.371 | FNTA/SAP30/BAMBI/HIPK2/LDLRAD4 | 5 |
| BP | GO:0022010 | central nervous system myelination | 7.07E-03 | 0.371 | MAG/MOBP | 2 |
| BP | GO:0032291 | axon ensheathment in central nervous system | 7.07E-03 | 0.371 | MAG/MOBP | 2 |
| BP | GO:0030010 | establishment of cell polarity | 7.42E-03 | 0.371 | NDE1/FAT1/FEZ1/LLGL2 | 4 |
| BP | GO:2000819 | regulation of nucleotide-excision repair | 7.59E-03 | 0.371 | PHF10/SMARCC1 | 2 |
| BP | GO:0048592 | eye morphogenesis | 7.90E-03 | 0.371 | FAT1/USH1C/HIPK2/BCAR3 | 4 |
| BP | GO:0002088 | lens development in camera-type eye | 8.99E-03 | 0.390 | FAT1/HIPK2/BCAR3 | 3 |
| BP | GO:0014850 | response to muscle activity | 9.25E-03 | 0.390 | CAPN3/TNS2 | 2 |
| BP | GO:0072384 | organelle transport along microtubule | 9.54E-03 | 0.390 | NDE1/FEZ1/ARHGAP21 | 3 |
| BP | GO:0032868 | response to insulin | 9.64E-03 | 0.390 | ZBED3/TNS2/BCAR3/PRKCQ/SMARCC1 | 5 |
| BP | GO:0010970 | transport along microtubule | 1.02E-02 | 0.390 | NDE1/FEZ1/ARHGAP21/CILK1 | 4 |
| BP | GO:0017015 | regulation of transforming growth factor beta receptor signaling pathway | 1.02E-02 | 0.390 | SAP30/BAMBI/HIPK2/LDLRAD4 | 4 |
| BP | GO:1903844 | regulation of cellular response to transforming growth factor beta stimulus | 1.08E-02 | 0.392 | SAP30/BAMBI/HIPK2/LDLRAD4 | 4 |
| BP | GO:0141091 | transforming growth factor beta receptor superfamily signaling pathway | 1.09E-02 | 0.392 | FNTA/SAP30/PCSK6/BAMBI/HIPK2/LDLRAD4 | 6 |
| BP | GO:0045664 | regulation of neuron differentiation | 1.31E-02 | 0.423 | S100B/MAG/CTDSP1/GPRC5B | 4 |
| BP | GO:0070316 | regulation of G0 to G1 transition | 1.37E-02 | 0.423 | PHF10/SMARCC1 | 2 |
| BP | GO:0071560 | cellular response to transforming growth factor beta stimulus | 1.45E-02 | 0.423 | FNTA/SAP30/BAMBI/HIPK2/LDLRAD4 | 5 |
| BP | GO:0007178 | cell surface receptor protein serine/threonine kinase signaling pathway | 1.48E-02 | 0.423 | FNTA/SAP30/PCSK6/BAMBI/HIPK2/LDLRAD4 | 6 |
| BP | GO:0048709 | oligodendrocyte differentiation | 1.50E-02 | 0.423 | TMEM98/MAG/MOBP | 3 |
| BP | GO:0045023 | G0 to G1 transition | 1.51E-02 | 0.423 | PHF10/SMARCC1 | 2 |
| BP | GO:0090101 | negative regulation of transmembrane receptor protein | 1.58E-02 | 0.423 | SAP30/BAMBI/HIPK2/LDLRAD4 | 4 |

| | | | | | | |
|---|---|---|---|---|---|---|
| | | serine/threonine kinase signaling pathway | | | | |
| BP | GO:0009072 | aromatic amino acid metabolic process | 1.58E-02 | 0.423 | CARNS1/QDPR | 2 |
| BP | GO:0071559 | response to transforming growth factor beta | 1.59E-02 | 0.423 | FNTA/SAP30/BAMBI/HIPK2/LDLRAD4 | 5 |
| BP | GO:0030705 | cytoskeleton-dependent intracellular transport | 1.66E-02 | 0.423 | NDE1/FEZ1/ARHGAP21/CILK1 | 4 |
| BP | GO:0043393 | regulation of protein binding | 1.73E-02 | 0.423 | IFIT2/BAMBI/HIPK2 | 3 |
| BP | GO:0090263 | positive regulation of canonical Wnt signaling pathway | 1.73E-02 | 0.423 | ZBED3/BAMBI/GPRC5B | 3 |
| BP | GO:0071375 | cellular response to peptide hormone stimulus | 1.75E-02 | 0.423 | FAT1/TNS2/BCAR3/PRKCQ/SMARCC1 | 5 |
| BP | GO:0032869 | cellular response to insulin stimulus | 1.79E-02 | 0.423 | TNS2/BCAR3/PRKCQ/SMARCC1 | 4 |
| BP | GO:0046627 | negative regulation of insulin receptor signaling pathway | 1.81E-02 | 0.423 | TNS2/PRKCQ | 2 |
| BP | GO:0090092 | regulation of transmembrane receptor protein serine/threonine kinase signaling pathway | 1.86E-02 | 0.423 | SAP30/PCSK6/BAMBI/HIPK2/LDLRAD4 | 5 |
| BP | GO:1900077 | negative regulation of cellular response to insulin stimulus | 1.89E-02 | 0.423 | TNS2/PRKCQ | 2 |
| BP | GO:0045663 | positive regulation of myoblast differentiation | 1.97E-02 | 0.423 | PHF10/SMARCC1 | 2 |
| BP | GO:0051294 | establishment of spindle orientation | 1.97E-02 | 0.423 | NDE1/LLGL2 | 2 |
| BP | GO:0051092 | positive regulation of NF-kappaB transcription factor activity | 1.97E-02 | 0.423 | CAPN3/PRKCQ/RAB7B | 3 |
| BP | GO:0031641 | regulation of myelination | 2.05E-02 | 0.423 | TMEM98/MAG | 2 |
| BP | GO:0030510 | regulation of BMP signaling pathway | 2.06E-02 | 0.423 | PCSK6/BAMBI/HIPK2 | 3 |
| BP | GO:0031644 | regulation of nervous system process | 2.06E-02 | 0.423 | TMEM98/MAG/TAFA4 | 3 |
| BP | GO:0006486 | protein glycosylation | 2.08E-02 | 0.423 | TMTC2/B3GNT9/TMTC4/GALNT6 | 4 |
| BP | GO:0043413 | macromolecule glycosylation | 2.08E-02 | 0.423 | TMTC2/B3GNT9/TMTC4/GALNT6 | 4 |
| BP | GO:0099111 | microtubule-based transport | 2.14E-02 | 0.423 | NDE1/FEZ1/ARHGAP21/CILK1 | 4 |
| BP | GO:0007163 | establishment or maintenance of cell polarity | 2.21E-02 | 0.423 | NDE1/FAT1/FEZ1/LLGL2 | 4 |

| | | | | | | |
|---|---|---|---|---|---|---|
| BP | GO:0006509 | membrane protein ectodomain proteolysis | 2.22E-02 | 0.423 | TSPAN15/PRKCQ | 2 |
| BP | GO:0032092 | positive regulation of protein binding | 2.22E-02 | 0.423 | BAMBI/HIPK2 | 2 |
| BP | GO:0045197 | establishment or maintenance of epithelial cell apical/basal polarity | 2.22E-02 | 0.423 | FAT1/LLGL2 | 2 |
| BP | GO:0014003 | oligodendrocyte development | 2.30E-02 | 0.433 | MAG/MOBP | 2 |
| BP | GO:0030512 | negative regulation of transforming growth factor beta receptor signaling pathway | 2.42E-02 | 0.442 | SAP30/BAMBI/LDLRAD4 | 3 |
| BP | GO:0045214 | sarcomere organization | 2.48E-02 | 0.442 | FLNC/CAPN3 | 2 |
| BP | GO:0007018 | microtubule-based movement | 2.53E-02 | 0.442 | KLC1/NDE1/FEZ1/ARHGAP21/SEPTIN4/CILK1 | 6 |
| BP | GO:0035088 | establishment or maintenance of apical/basal cell polarity | 2.57E-02 | 0.442 | FAT1/LLGL2 | 2 |
| BP | GO:0051646 | mitochondrion localization | 2.57E-02 | 0.442 | ZBED3/FEZ1 | 2 |
| BP | GO:0061245 | establishment or maintenance of bipolar cell polarity | 2.57E-02 | 0.442 | FAT1/LLGL2 | 2 |
| BP | GO:0070085 | glycosylation | 2.71E-02 | 0.453 | TMTC2/B3GNT9/TMTC4/GALNT6 | 4 |
| BP | GO:0050768 | negative regulation of neurogenesis | 2.71E-02 | 0.453 | TMEM98/MAG/CTDSP1 | 3 |
| BP | GO:0051090 | regulation of DNA-binding transcription factor activity | 2.80E-02 | 0.453 | CAPN3/LRP6/HIPK2/PRKCQ/RAB7B | 5 |
| BP | GO:0061041 | regulation of wound healing | 2.81E-02 | 0.453 | MYLK/TSPAN8/VKORC1 | 3 |
| BP | GO:2000059 | negative regulation of ubiquitin-dependent protein catabolic process | 2.84E-02 | 0.453 | HIPK2/SMARCC1 | 2 |
| BP | GO:0071470 | cellular response to osmotic stress | 2.94E-02 | 0.453 | CAPN3/MYLK | 2 |
| BP | GO:0051961 | negative regulation of nervous system development | 3.13E-02 | 0.453 | TMEM98/MAG/CTDSP1 | 3 |
| BP | GO:0090287 | regulation of cellular response to growth factor stimulus | 3.35E-02 | 0.453 | SAP30/PCSK6/BAMBI/HIPK2/LDLRAD4 | 5 |
| BP | GO:0030177 | positive regulation of Wnt signaling pathway | 3.47E-02 | 0.453 | ZBED3/BAMBI/GPRC5B | 3 |
| BP | GO:0033619 | membrane protein proteolysis | 3.63E-02 | 0.453 | TSPAN15/PRKCQ | 2 |
| BP | GO:0009100 | glycoprotein metabolic process | 3.71E-02 | 0.453 | TMTC2/B3GNT9/TMTC4/PCSK6/GALNT6 | 5 |
| BP | GO:0042552 | myelination | 3.82E-02 | 0.453 | TMEM98/MAG/MOBP | 3 |

| | | | | | | |
|---|---|---|---|---|---|---|
| BP | GO:0007272 | ensheathment of neurons | 3.94E-02 | 0.453 | TMEM98/MAG/MOBP | 3 |
| BP | GO:0008366 | axon ensheathment | 3.94E-02 | 0.453 | TMEM98/MAG/MOBP | 3 |
| BP | GO:0060042 | retina morphogenesis in camera-type eye | 3.95E-02 | 0.453 | USH1C/HIPK2 | 2 |
| BP | GO:0030193 | regulation of blood coagulation | 4.16E-02 | 0.453 | TSPAN8/VKORC1 | 2 |
| BP | GO:0170040 | proteinogenic amino acid catabolic process | 4.38E-02 | 0.453 | CARNS1/QDPR | 2 |
| BP | GO:1900046 | regulation of hemostasis | 4.38E-02 | 0.453 | TSPAN8/VKORC1 | 2 |
| BP | GO:0030199 | collagen fibril organization | 4.49E-02 | 0.453 | ADAMTS14/CRTAP | 2 |
| BP | GO:0046626 | regulation of insulin receptor signaling pathway | 4.49E-02 | 0.453 | TNS2/PRKCQ | 2 |
| BP | GO:0090596 | sensory organ morphogenesis | 4.51E-02 | 0.453 | FAT1/USH1C/HIPK2/BCAR3 | 4 |
| BP | GO:0006702 | androgen biosynthetic process | 4.59E-02 | 0.453 | HSD17B3 | 1 |
| BP | GO:0014820 | tonic smooth muscle contraction | 4.59E-02 | 0.453 | MYLK | 1 |
| BP | GO:0015820 | L-leucine transport | 4.59E-02 | 0.453 | LLGL2 | 1 |
| BP | GO:0018243 | protein O-linked glycosylation via threonine | 4.59E-02 | 0.453 | GALNT6 | 1 |
| BP | GO:1901856 | negative regulation of cellular respiration | 4.59E-02 | 0.453 | PPIF | 1 |
| BP | GO:1902513 | regulation of organelle transport along microtubule | 4.59E-02 | 0.453 | FEZ1 | 1 |
| BP | GO:1903799 | negative regulation of miRNA processing | 4.59E-02 | 0.453 | BCDIN3D | 1 |
| BP | GO:1903911 | positive regulation of receptor clustering | 4.59E-02 | 0.453 | FNTA | 1 |
| BP | GO:2000563 | positive regulation of CD4-positive, alpha-beta T cell proliferation | 4.59E-02 | 0.453 | PRKCQ | 1 |
| BP | GO:0045665 | negative regulation of neuron differentiation | 4.61E-02 | 0.453 | MAG/CTDSP1 | 2 |
| BP | GO:0030509 | BMP signaling pathway | 4.71E-02 | 0.453 | PCSK6/BAMBI/HIPK2 | 3 |
| BP | GO:0030514 | negative regulation of BMP signaling pathway | 4.72E-02 | 0.453 | BAMBI/HIPK2 | 2 |
| BP | GO:1903051 | negative regulation of proteolysis involved in protein catabolic process | 4.72E-02 | 0.453 | HIPK2/SMARCC1 | 2 |
| BP | GO:0006023 | aminoglycan biosynthetic process | 4.84E-02 | 0.453 | B3GNT9/CHST6 | 2 |

| | | | | | | |
|---|---|---|---|---|---|---|
| BP | GO:0170035 | L-amino acid catabolic process | 4.84E-02 | 0.453 | CARNS1/QDPR | 2 |
| BP | GO:0050818 | regulation of coagulation | 4.95E-02 | 0.453 | TSPAN8/VKORC1 | 2 |
| CC | GO:0071564 | npBAF complex | 1.92E-03 | 0.170 | PHF10/SMARCC1 | 2 |
| CC | GO:0016586 | RSC-type complex | 2.21E-03 | 0.170 | PHF10/SMARCC1 | 2 |
| CC | GO:0032153 | cell division site | 4.06E-03 | 0.208 | NDE1/MYLK/SEPTIN4 | 3 |
| CC | GO:0005875 | microtubule associated complex | 6.49E-03 | 0.233 | KLC1/NDE1/FNTA/KATNA1 | 4 |
| CC | GO:0016514 | SWI/SNF complex | 8.76E-03 | 0.233 | PHF10/SMARCC1 | 2 |
| CC | GO:0000776 | kinetochore | 9.08E-03 | 0.233 | RASSF2/NDE1/PHF10/SMARCC1 | 4 |
| CC | GO:0000779 | condensed chromosome, centromeric region | 1.14E-02 | 0.236 | RASSF2/NDE1/PHF10/SMARCC1 | 4 |
| CC | GO:0005912 | adherens junction | 1.23E-02 | 0.236 | CTNNA3/CDH20/LLGL2/FRMD5 | 4 |
| CC | GO:0005925 | focal adhesion | 1.51E-02 | 0.257 | FLNC/ADGRE5/FAT1/TNS2/BCAR3/LAP3 | 6 |
| CC | GO:0030055 | cell-substrate junction | 1.67E-02 | 0.257 | FLNC/ADGRE5/FAT1/TNS2/BCAR3/LAP3 | 6 |
| CC | GO:0005871 | kinesin complex | 2.15E-02 | 0.287 | KLC1/NDE1 | 2 |
| CC | GO:0001673 | male germ cell nucleus | 2.50E-02 | 0.287 | TCFL5/SMARCC1 | 2 |
| CC | GO:0032154 | cleavage furrow | 2.68E-02 | 0.287 | NDE1/MYLK | 2 |
| CC | GO:0000775 | chromosome, centromeric region | 3.53E-02 | 0.287 | RASSF2/NDE1/PHF10/SMARCC1 | 4 |
| CC | GO:0005802 | trans-Golgi network | 3.83E-02 | 0.287 | CHST6/RAB7B/DENND5A/LAP3 | 4 |
| CC | GO:0043073 | germ cell nucleus | 4.09E-02 | 0.287 | TCFL5/SMARCC1 | 2 |
| CC | GO:0000793 | condensed chromosome | 4.53E-02 | 0.287 | RASSF2/NDE1/PHF10/SMARCC1 | 4 |
| CC | GO:0001917 | photoreceptor inner segment | 4.53E-02 | 0.287 | RDH11/USH1C | 2 |
| CC | GO:0097197 | tetraspanin-enriched microdomain | 4.61E-02 | 0.287 | TSPAN15 | 1 |
| CC | GO:0031902 | late endosome membrane | 4.98E-02 | 0.287 | SLC31A2/TSPAN15/RAB7B | 3 |
| MF | GO:0030695 | GTPase regulator activity | 3.48E-03 | 0.360 | DENND11/ARHGEF1/ARHGAP21/BCAR3/RGS1/ARHGEF37/DENND5A/LLGL2 | 8 |
| MF | GO:0060589 | nucleoside-triphosphatase regulator activity | 3.48E-03 | 0.360 | DENND11/ARHGEF1/ARHGAP21/BCAR3/RGS1/ARHGEF37/DENND5A/LLGL2 | 8 |
| MF | GO:0005085 | guanyl-nucleotide exchange factor activity | 6.32E-03 | 0.360 | DENND11/ARHGEF1/BCAR3/ARHGEF37/DENND5A | 5 |
| MF | GO:0000030 | mannosyltransferase activity | 7.07E-03 | 0.360 | TMTC2/TMTC4 | 2 |
| MF | GO:0070006 | metalloaminopeptidase activity | 7.64E-03 | 0.360 | AOPEP/LAP3 | 2 |

| | | | | | | |
|---|---|---|---|---|---|---|
| MF | GO:0016757 | glycosyltransferase activity | 1.16E-02 | 0.384 | TMTC2/B3GNT9/TMTC4/PARP4/GALNT6 | 5 |
| MF | GO:0008237 | metallopeptidase activity | 1.46E-02 | 0.384 | AOPEP/ADAMTS4/ADAMTS14/LAP3 | 4 |
| MF | GO:0005109 | frizzled binding | 1.59E-02 | 0.384 | LRP6/BAMBI | 2 |
| MF | GO:0004177 | aminopeptidase activity | 1.75E-02 | 0.384 | AOPEP/LAP3 | 2 |
| MF | GO:0016758 | hexosyltransferase activity | 1.80E-02 | 0.384 | TMTC2/B3GNT9/TMTC4/GALNT6 | 4 |
| MF | GO:0038024 | cargo receptor activity | 4.30E-02 | 0.384 | LRP6/DMBT1 | 2 |
| MF | GO:0008235 | metalloexopeptidase activity | 4.55E-02 | 0.384 | AOPEP/LAP3 | 2 |
| MF | GO:0043121 | neurotrophin binding | 4.96E-02 | 0.384 | PCSK6 | 1 |
| MF | GO:0043208 | glycosphingolipid binding | 4.96E-02 | 0.384 | MAG | 1 |
| MF | GO:0047429 | nucleoside triphosphate diphosphatase activity | 4.96E-02 | 0.384 | NUDT1 | 1 |
| MF | GO:0005096 | GTPase activator activity | 4.96E-02 | 0.384 | ARHGEF1/ARHGAP21/RGS1/LLGL2 | 4 |

Abbreviations: BP=biological process; CC=cellular component; MF=molecular function.

#### Supplementary Table 11: Exploratory gene ontology analysis of the 70 transcripts that contribute heavily to Component 7 and correlate significantly with normative C9orf72 expression.

| **Ontology** | **ID** | **Description** | ***p*** | ***q*** | **Contributory Genes** | **Gene Count** |
|---|---|---|---|---|---|---|
| BP | GO:0007179 | transforming growth factor beta receptor signaling pathway | 1.54E-03 | 0.277 | LDLRAD4/FNTA/HIPK2/SAP30/BAMBI | 5 |
| BP | GO:0141091 | transforming growth factor beta receptor superfamily signaling pathway | 2.17E-03 | 0.277 | LDLRAD4/FNTA/HIPK2/SAP30/PCSK6/BAMBI | 6 |
| BP | GO:0002089 | lens morphogenesis in camera-type eye | 2.47E-03 | 0.277 | HIPK2/BCAR3 | 2 |
| BP | GO:0009074 | aromatic amino acid family catabolic process | 2.94E-03 | 0.277 | CARNS1/QDPR | 2 |
| BP | GO:0032288 | myelin assembly | 2.94E-03 | 0.277 | MAG/MOBP | 2 |
| BP | GO:0007178 | cell surface receptor protein serine/threonine kinase signaling pathway | 3.05E-03 | 0.277 | LDLRAD4/FNTA/HIPK2/SAP30/PCSK6/BAMBI | 6 |

| | | | | | | |
|---|---|---|---|---|---|---|
| BP | GO:0017015 | regulation of transforming growth factor beta receptor signaling pathway | 3.19E-03 | 0.277 | LDLRAD4/HIPK2/SAP30/BAMBI | 4 |
| BP | GO:1903844 | regulation of cellular response to transforming growth factor beta stimulus | 3.38E-03 | 0.277 | LDLRAD4/HIPK2/SAP30/BAMBI | 4 |
| BP | GO:0071560 | cellular response to transforming growth factor beta stimulus | 3.69E-03 | 0.277 | LDLRAD4/FNTA/HIPK2/SAP30/BAMBI | 5 |
| BP | GO:0022010 | central nervous system myelination | 3.72E-03 | 0.277 | MAG/MOBP | 2 |
| BP | GO:0032291 | axon ensheathment in central nervous system | 3.72E-03 | 0.277 | MAG/MOBP | 2 |
| BP | GO:0071559 | response to transforming growth factor beta | 4.06E-03 | 0.277 | LDLRAD4/FNTA/HIPK2/SAP30/BAMBI | 5 |
| BP | GO:0045664 | regulation of neuron differentiation | 4.16E-03 | 0.277 | GPRC5B/MAG/CTDSP1/S100B | 4 |
| BP | GO:0006493 | protein O-linked glycosylation | 4.68E-03 | 0.277 | GALNT6/B3GNT9/TMTC2 | 3 |
| BP | GO:0090092 | regulation of transmembrane receptor protein serine/threonine kinase signaling pathway | 4.82E-03 | 0.277 | LDLRAD4/HIPK2/SAP30/PCSK6/BAMBI | 5 |
| BP | GO:0014850 | response to muscle activity | 4.88E-03 | 0.277 | CAPN3/TNS2 | 2 |
| BP | GO:0090101 | negative regulation of transmembrane receptor protein serine/threonine kinase signaling pathway | 5.06E-03 | 0.277 | LDLRAD4/HIPK2/SAP30/BAMBI | 4 |
| BP | GO:0048709 | oligodendrocyte differentiation | 6.10E-03 | 0.315 | TMEM98/MAG/MOBP | 3 |
| BP | GO:0043393 | regulation of protein binding | 7.06E-03 | 0.331 | IFIT2/HIPK2/BAMBI | 3 |
| BP | GO:0009072 | aromatic amino acid metabolic process | 8.43E-03 | 0.331 | CARNS1/QDPR | 2 |
| BP | GO:0030510 | regulation of BMP signaling pathway | 8.48E-03 | 0.331 | HIPK2/PCSK6/BAMBI | 3 |
| BP | GO:0090287 | regulation of cellular response to growth factor stimulus | 9.18E-03 | 0.331 | LDLRAD4/HIPK2/SAP30/PCSK6/BAMBI | 5 |
| BP | GO:0030512 | negative regulation of transforming growth factor beta receptor signaling pathway | 1.01E-02 | 0.331 | LDLRAD4/SAP30/BAMBI | 3 |
| BP | GO:0051294 | establishment of spindle orientation | 1.05E-02 | 0.331 | NDE1/LLGL2 | 2 |
| BP | GO:0048593 | camera-type eye morphogenesis | 1.09E-02 | 0.331 | HIPK2/BCAR3/USH1C | 3 |

| | | | | | | |
|---|---|---|---|---|---|---|
| BP | GO:0031641 | regulation of myelination | 1.10E-02 | 0.331 | TMEM98/MAG | 2 |
| BP | GO:0050768 | negative regulation of neurogenesis | 1.13E-02 | 0.331 | TMEM98/MAG/CTDSP1 | 3 |
| BP | GO:0010927 | cellular component assembly involved in morphogenesis | 1.18E-02 | 0.331 | CAPN3/MAG/MOBP | 3 |
| BP | GO:0032989 | cellular anatomical entity morphogenesis | 1.18E-02 | 0.331 | CAPN3/MAG/MOBP | 3 |
| BP | GO:0061041 | regulation of wound healing | 1.18E-02 | 0.331 | MYLK/TSPAN8/VKORC1 | 3 |
| BP | GO:0032092 | positive regulation of protein binding | 1.19E-02 | 0.331 | HIPK2/BAMBI | 2 |
| BP | GO:2000785 | regulation of autophagosome assembly | 1.19E-02 | 0.331 | C9orf72/FEZ1 | 2 |
| BP | GO:0014003 | oligodendrocyte development | 1.23E-02 | 0.331 | MAG/MOBP | 2 |
| BP | GO:0051961 | negative regulation of nervous system development | 1.32E-02 | 0.331 | TMEM98/MAG/CTDSP1 | 3 |
| BP | GO:0071470 | cellular response to osmotic stress | 1.58E-02 | 0.331 | CAPN3/MYLK | 2 |
| BP | GO:0042552 | myelination | 1.63E-02 | 0.331 | TMEM98/MAG/MOBP | 3 |
| BP | GO:0007272 | ensheathment of neurons | 1.68E-02 | 0.331 | TMEM98/MAG/MOBP | 3 |
| BP | GO:0008366 | axon ensheathment | 1.68E-02 | 0.331 | TMEM98/MAG/MOBP | 3 |
| BP | GO:0044088 | regulation of vacuole organization | 1.75E-02 | 0.331 | C9orf72/FEZ1 | 2 |
| BP | GO:0030010 | establishment of cell polarity | 1.79E-02 | 0.331 | NDE1/FEZ1/LLGL2 | 3 |
| BP | GO:0051293 | establishment of spindle localization | 1.80E-02 | 0.331 | NDE1/LLGL2 | 2 |
| BP | GO:0048592 | eye morphogenesis | 1.88E-02 | 0.331 | HIPK2/BCAR3/USH1C | 3 |
| BP | GO:1990138 | neuron projection extension | 1.94E-02 | 0.331 | C9orf72/MAG/S100B | 3 |
| BP | GO:0030509 | BMP signaling pathway | 2.03E-02 | 0.331 | HIPK2/PCSK6/BAMBI | 3 |
| BP | GO:0051653 | spindle localization | 2.09E-02 | 0.331 | NDE1/LLGL2 | 2 |
| BP | GO:0060042 | retina morphogenesis in camera-type eye | 2.15E-02 | 0.331 | HIPK2/USH1C | 2 |
| BP | GO:0030193 | regulation of blood coagulation | 2.27E-02 | 0.331 | TSPAN8/VKORC1 | 2 |
| BP | GO:0007018 | microtubule-based movement | 2.28E-02 | 0.331 | KLC1/NDE1/SEPTIN4/FEZ1/CILK1 | 5 |
| BP | GO:0010970 | transport along microtubule | 2.29E-02 | 0.331 | NDE1/FEZ1/CILK1 | 3 |
| BP | GO:1903034 | regulation of response to wounding | 2.32E-02 | 0.331 | MYLK/TSPAN8/VKORC1 | 3 |
| BP | GO:0170040 | proteinogenic amino acid catabolic process | 2.39E-02 | 0.331 | CARNS1/QDPR | 2 |
| BP | GO:1900046 | regulation of hemostasis | 2.39E-02 | 0.331 | TSPAN8/VKORC1 | 2 |

| | | | | | | |
|---|---|---|---|---|---|---|
| BP | GO:0071772 | response to BMP | 2.42E-02 | 0.331 | HIPK2/PCSK6/BAMBI | 3 |
| BP | GO:0071773 | cellular response to BMP stimulus | 2.42E-02 | 0.331 | HIPK2/PCSK6/BAMBI | 3 |
| BP | GO:0007249 | canonical NF-kappaB signal transduction | 2.45E-02 | 0.331 | GPRC5B/RASSF2/CAPN3/S100B | 4 |
| BP | GO:0045665 | negative regulation of neuron differentiation | 2.52E-02 | 0.331 | MAG/CTDSP1 | 2 |
| BP | GO:0030514 | negative regulation of BMP signaling pathway | 2.59E-02 | 0.331 | HIPK2/BAMBI | 2 |
| BP | GO:0006023 | aminoglycan biosynthetic process | 2.65E-02 | 0.331 | B3GNT9/CHST6 | 2 |
| BP | GO:0170035 | L-amino acid catabolic process | 2.65E-02 | 0.331 | CARNS1/QDPR | 2 |
| BP | GO:0045661 | regulation of myoblast differentiation | 2.72E-02 | 0.331 | CMTM5/CAPN3 | 2 |
| BP | GO:0050818 | regulation of coagulation | 2.72E-02 | 0.331 | TSPAN8/VKORC1 | 2 |
| BP | GO:0031400 | negative regulation of protein modification process | 2.82E-02 | 0.331 | C9orf72/RASSF2/CAPN3/HHATL | 4 |
| BP | GO:0071277 | cellular response to calcium ion | 2.85E-02 | 0.331 | PPIF/CAPN3 | 2 |
| BP | GO:0035914 | skeletal muscle cell differentiation | 2.99E-02 | 0.331 | PPIF/SAP30 | 2 |
| BP | GO:0045685 | regulation of glial cell differentiation | 2.99E-02 | 0.331 | TMEM98/MAG | 2 |
| BP | GO:0014823 | response to activity | 3.13E-02 | 0.331 | CAPN3/TNS2 | 2 |
| BP | GO:0030705 | cytoskeleton-dependent intracellular transport | 3.31E-02 | 0.331 | NDE1/FEZ1/CILK1 | 3 |
| BP | GO:0051091 | positive regulation of DNA-binding transcription factor activity | 3.31E-02 | 0.331 | CAPN3/HIPK2/RAB7B | 3 |
| BP | GO:0006702 | androgen biosynthetic process | 3.32E-02 | 0.331 | HSD17B3 | 1 |
| BP | GO:0014820 | tonic smooth muscle contraction | 3.32E-02 | 0.331 | MYLK | 1 |
| BP | GO:0015820 | L-leucine transport | 3.32E-02 | 0.331 | LLGL2 | 1 |
| BP | GO:0018243 | protein O-linked glycosylation via threonine | 3.32E-02 | 0.331 | GALNT6 | 1 |
| BP | GO:1901856 | negative regulation of cellular respiration | 3.32E-02 | 0.331 | PPIF | 1 |
| BP | GO:1902513 | regulation of organelle transport along microtubule | 3.32E-02 | 0.331 | FEZ1 | 1 |
| BP | GO:1903799 | negative regulation of miRNA processing | 3.32E-02 | 0.331 | BCDIN3D | 1 |

| | | | | | | |
|---|---|---|---|---|---|---|
| BP | GO:1903911 | positive regulation of receptor clustering | 3.32E-02 | 0.331 | FNTA | 1 |
| BP | GO:0006970 | response to osmotic stress | 3.34E-02 | 0.331 | CAPN3/MYLK | 2 |
| BP | GO:0003407 | neural retina development | 3.56E-02 | 0.331 | HIPK2/USH1C | 2 |
| BP | GO:0007517 | muscle organ development | 3.58E-02 | 0.331 | PPIF/CAPN3/MYLK/SAP30 | 4 |
| BP | GO:0018210 | peptidyl-threonine modification | 3.63E-02 | 0.331 | GALNT6/HIPK2 | 2 |
| BP | GO:0000098 | sulfur amino acid catabolic process | 3.65E-02 | 0.331 | TST | 1 |
| BP | GO:0032060 | bleb assembly | 3.65E-02 | 0.331 | MYLK | 1 |
| BP | GO:0033234 | negative regulation of protein sumoylation | 3.65E-02 | 0.331 | CAPN3 | 1 |
| BP | GO:0042373 | vitamin K metabolic process | 3.65E-02 | 0.331 | VKORC1 | 1 |
| BP | GO:0045654 | positive regulation of megakaryocyte differentiation | 3.65E-02 | 0.331 | RAB7B | 1 |
| BP | GO:0046549 | retinal cone cell development | 3.65E-02 | 0.331 | USH1C | 1 |
| BP | GO:1903059 | regulation of protein lipidation | 3.65E-02 | 0.331 | HHATL | 1 |
| BP | GO:0051098 | regulation of binding | 3.77E-02 | 0.331 | IFIT2/HIPK2/BAMBI | 3 |
| BP | GO:0002088 | lens development in camera-type eye | 3.78E-02 | 0.331 | HIPK2/BCAR3 | 2 |
| BP | GO:0072384 | organelle transport along microtubule | 3.93E-02 | 0.331 | NDE1/FEZ1 | 2 |
| BP | GO:0006486 | protein glycosylation | 3.95E-02 | 0.331 | GALNT6/B3GNT9/TMTC2 | 3 |
| BP | GO:0043413 | macromolecule glycosylation | 3.95E-02 | 0.331 | GALNT6/B3GNT9/TMTC2 | 3 |
| BP | GO:0015791 | polyol transmembrane transport | 3.97E-02 | 0.331 | SLC5A11 | 1 |
| BP | GO:0018342 | protein prenylation | 3.97E-02 | 0.331 | FNTA | 1 |
| BP | GO:0034163 | regulation of toll-like receptor 9 signaling pathway | 3.97E-02 | 0.331 | RAB7B | 1 |
| BP | GO:0035022 | positive regulation of Rac protein signal transduction | 3.97E-02 | 0.331 | FNTA | 1 |
| BP | GO:0061370 | testosterone biosynthetic process | 3.97E-02 | 0.331 | HSD17B3 | 1 |
| BP | GO:0071476 | cellular hypotonic response | 3.97E-02 | 0.331 | MYLK | 1 |
| BP | GO:0086100 | endothelin receptor signaling pathway | 3.97E-02 | 0.331 | BCAR3 | 1 |
| BP | GO:0097264 | self proteolysis | 3.97E-02 | 0.331 | CAPN3 | 1 |
| BP | GO:0097354 | prenylation | 3.97E-02 | 0.331 | FNTA | 1 |

| | | | | | | |
|---|---|---|---|---|---|---|
| BP | GO:0098528 | skeletal muscle fiber differentiation | 3.97E-02 | 0.331 | PPIF | 1 |
| BP | GO:0098911 | regulation of ventricular cardiac muscle cell action potential | 3.97E-02 | 0.331 | CTNNA3 | 1 |
| BP | GO:0099111 | microtubule-based transport | 4.04E-02 | 0.331 | NDE1/FEZ1/CILK1 | 3 |
| BP | GO:0007163 | establishment or maintenance of cell polarity | 4.13E-02 | 0.331 | NDE1/FEZ1/LLGL2 | 3 |
| BP | GO:0045666 | positive regulation of neuron differentiation | 4.17E-02 | 0.331 | GPRC5B/S100B | 2 |
| BP | GO:0046330 | positive regulation of JNK cascade | 4.24E-02 | 0.331 | RASSF2/HIPK2 | 2 |
| BP | GO:0060395 | SMAD protein signal transduction | 4.24E-02 | 0.331 | LDLRAD4/HIPK2 | 2 |
| BP | GO:0010269 | response to selenium ion | 4.30E-02 | 0.331 | SELENOP | 1 |
| BP | GO:0032530 | regulation of microvillus organization | 4.30E-02 | 0.331 | USH1C | 1 |
| BP | GO:0048711 | positive regulation of astrocyte differentiation | 4.30E-02 | 0.331 | MAG | 1 |
| BP | GO:0052803 | imidazole-containing compound metabolic process | 4.30E-02 | 0.331 | CARNS1 | 1 |
| BP | GO:0071340 | skeletal muscle acetylcholine-gated channel clustering | 4.30E-02 | 0.331 | FNTA | 1 |
| BP | GO:0075509 | endocytosis involved in viral entry into host cell | 4.30E-02 | 0.331 | JPT2 | 1 |
| BP | GO:1902902 | negative regulation of autophagosome assembly | 4.30E-02 | 0.331 | FEZ1 | 1 |
| BP | GO:0009100 | glycoprotein metabolic process | 4.32E-02 | 0.331 | GALNT6/PCSK6/B3GNT9/TMTC2 | 4 |
| BP | GO:1901606 | alpha-amino acid catabolic process | 4.32E-02 | 0.331 | CARNS1/QDPR | 2 |
| BP | GO:0048588 | developmental cell growth | 4.35E-02 | 0.331 | C9orf72/MAG/S100B | 3 |
| BP | GO:0006971 | hypotonic response | 4.62E-02 | 0.331 | MYLK | 1 |
| BP | GO:0007100 | mitotic centrosome separation | 4.62E-02 | 0.331 | NDE1 | 1 |
| BP | GO:0010155 | regulation of proton transport | 4.62E-02 | 0.331 | PPIF | 1 |
| BP | GO:0015803 | branched-chain amino acid transport | 4.62E-02 | 0.331 | LLGL2 | 1 |
| BP | GO:0032536 | regulation of cell projection size | 4.62E-02 | 0.331 | USH1C | 1 |
| BP | GO:0035721 | intraciliary retrograde transport | 4.62E-02 | 0.331 | CILK1 | 1 |

| | | | | | | |
|---|---|---|---|---|---|---|
| BP | GO:0042559 | pteridine-containing compound biosynthetic process | 4.62E-02 | 0.331 | QDPR | 1 |
| BP | GO:0042670 | retinal cone cell differentiation | 4.62E-02 | 0.331 | USH1C | 1 |
| BP | GO:0048715 | negative regulation of oligodendrocyte differentiation | 4.62E-02 | 0.331 | TMEM98 | 1 |
| BP | GO:0071472 | cellular response to salt stress | 4.62E-02 | 0.331 | CAPN3 | 1 |
| BP | GO:0090385 | phagosome-lysosome fusion | 4.62E-02 | 0.331 | RAB7B | 1 |
| BP | GO:0060560 | developmental growth involved in morphogenesis | 4.64E-02 | 0.331 | C9orf72/MAG/S100B | 3 |
| BP | GO:0042147 | retrograde transport, endosome to Golgi | 4.73E-02 | 0.331 | DENND5A/RAB7B | 2 |
| BP | GO:0007033 | vacuole organization | 4.73E-02 | 0.331 | C9orf72/FEZ1/RAB7B | 3 |
| BP | GO:0070085 | glycosylation | 4.83E-02 | 0.331 | GALNT6/B3GNT9/TMTC2 | 3 |
| BP | GO:0086069 | bundle of His cell to Purkinje myocyte communication | 4.94E-02 | 0.331 | CTNNA3 | 1 |
| BP | GO:0098693 | regulation of synaptic vesicle cycle | 4.94E-02 | 0.331 | C9orf72 | 1 |
| CC | GO:0032153 | cell division site | 1.55E-03 | 0.188 | MYLK/NDE1/SEPTIN4 | 3 |
| CC | GO:0005871 | kinesin complex | 1.13E-02 | 0.286 | KLC1/NDE1 | 2 |
| CC | GO:0032154 | cleavage furrow | 1.42E-02 | 0.286 | MYLK/NDE1 | 2 |
| CC | GO:0005875 | microtubule associated complex | 1.58E-02 | 0.286 | KLC1/FNTA/NDE1 | 3 |
| CC | GO:0030426 | growth cone | 1.89E-02 | 0.286 | C9orf72/KLC1/FEZ1 | 3 |
| CC | GO:0030427 | site of polarized growth | 2.04E-02 | 0.286 | C9orf72/KLC1/FEZ1 | 3 |
| CC | GO:0044304 | main axon | 2.50E-02 | 0.286 | C9orf72/MAG | 2 |
| CC | GO:0005912 | adherens junction | 2.57E-02 | 0.286 | CTNNA3/FRMD5/LLGL2 | 3 |
| CC | GO:0030670 | phagocytic vesicle membrane | 3.10E-02 | 0.286 | DMBT1/RAB7B | 2 |
| CC | GO:0032476 | polyprenyl diphosphate synthase complex | 3.63E-02 | 0.286 | FNTA | 1 |
| CC | GO:0033270 | paranode region of axon | 3.96E-02 | 0.286 | MAG | 1 |
| CC | GO:0043220 | Schmidt-Lanterman incisure | 3.96E-02 | 0.286 | MAG | 1 |
| CC | GO:0010494 | cytoplasmic stress granule | 3.98E-02 | 0.286 | C9orf72/HIPK2 | 2 |
| CC | GO:0031616 | spindle pole centrosome | 4.28E-02 | 0.286 | NDE1 | 1 |
| CC | GO:0005940 | septin ring | 4.60E-02 | 0.286 | SEPTIN4 | 1 |
| CC | GO:0031089 | platelet dense granule lumen | 4.60E-02 | 0.286 | SELENOP | 1 |

| | | | | | | |
|---|---|---|---|---|---|---|
| CC | GO:0031105 | septin complex | 4.60E-02 | 0.286 | SEPTIN4 | 1 |
| CC | GO:0043218 | compact myelin | 4.92E-02 | 0.286 | MAG | 1 |
| CC | GO:0098691 | dopaminergic synapse | 4.92E-02 | 0.286 | SEPTIN4 | 1 |
| MF | GO:0005085 | guanyl-nucleotide exchange factor activity | 9.76E-03 | 0.306 | C9orf72/ARHGEF37/DENND5A/BCAR3 | 4 |
| MF | GO:0016757 | glycosyltransferase activity | 1.61E-02 | 0.306 | GALNT6/PARP4/B3GNT9/TMTC2 | 4 |
| MF | GO:0016782 | transferase activity, transferring sulphur-containing groups | 2.75E-02 | 0.306 | TST/CHST6 | 2 |
| MF | GO:0030695 | GTPase regulator activity | 3.28E-02 | 0.306 | C9orf72/ARHGEF37/DENND5A/BCAR3/LLGL2 | 5 |
| MF | GO:0060589 | nucleoside-triphosphatase regulator activity | 3.28E-02 | 0.306 | C9orf72/ARHGEF37/DENND5A/BCAR3/LLGL2 | 5 |
| MF | GO:0046332 | SMAD binding | 3.34E-02 | 0.306 | LDLRAD4/HIPK2 | 2 |
| MF | GO:0048306 | calcium-dependent protein binding | 3.34E-02 | 0.306 | DMBT1/S100B | 2 |
| MF | GO:0016758 | hexosyltransferase activity | 3.48E-02 | 0.306 | GALNT6/B3GNT9/TMTC2 | 3 |
| MF | GO:0043121 | neurotrophin binding | 3.57E-02 | 0.306 | PCSK6 | 1 |
| MF | GO:0043208 | glycosphingolipid binding | 3.57E-02 | 0.306 | MAG | 1 |
| MF | GO:0005114 | type II transforming growth factor beta receptor binding | 3.92E-02 | 0.306 | BAMBI | 1 |
| MF | GO:0016725 | oxidoreductase activity, acting on CH or CH2 groups | 3.92E-02 | 0.306 | VKORC1 | 1 |
| MF | GO:0016783 | sulfurtransferase activity | 3.92E-02 | 0.306 | TST | 1 |
| MF | GO:0019911 | structural constituent of myelin sheath | 3.92E-02 | 0.306 | MOBP | 1 |
| MF | GO:0030283 | testosterone dehydrogenase [NAD(P)+] activity | 3.92E-02 | 0.306 | HSD17B3 | 1 |
| MF | GO:0050786 | RAGE receptor binding | 3.92E-02 | 0.306 | S100B | 1 |
| MF | GO:0008420 | RNA polymerase II CTD heptapeptide repeat phosphatase activity | 4.62E-02 | 0.306 | CTDSP1 | 1 |
| MF | GO:0015166 | polyol transmembrane transporter activity | 4.62E-02 | 0.306 | SLC5A11 | 1 |
| MF | GO:0031432 | titin binding | 4.62E-02 | 0.306 | CAPN3 | 1 |
| MF | GO:0045159 | myosin II binding | 4.62E-02 | 0.306 | LLGL2 | 1 |
| MF | GO:0044548 | S100 protein binding | 4.96E-02 | 0.306 | S100B | 1 |

Abbreviations: BP=biological process; CC=cellular component; MF=molecular function.

### Exploratory Gene Ontology (GO) Analyses for Hypothesis-Driven Transcripts

GO analysis of the 21 hypothesis-driven transcripts and *C9orf72* revealed 84 terms (72 biological processes and 12 cellular components), all of which survived Benjamini-Hochberg correction **(Supplementary Figure 2; Supplementary Table 12)**.

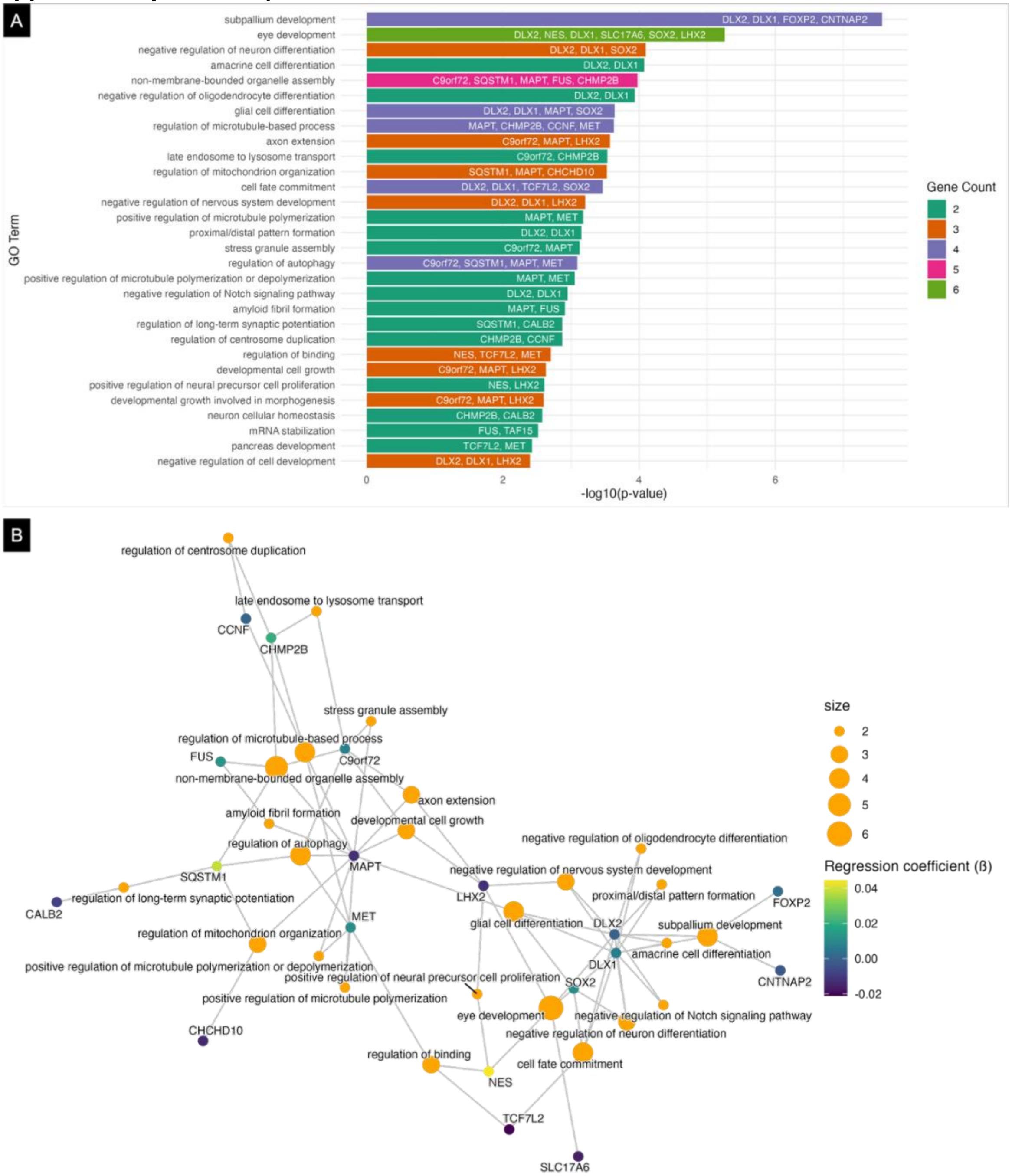

**Supplementary Figure 2. Exploratory gene ontology (GO) findings of C9orf72 and the identified 21 transcripts in a (A) bar graph and (B) gene-concept network plot.** All 84 GO terms survived corrections for multiple comparisons, and only the top 30 and 25 are visualized. Where two highly similar terms were present, as determined using simplify() from clusterProfiler, the higher-ranked of the two was plotted. To facilitate understanding of potential relationships of the 37 transcripts with *C9orf72*, GO terms which involved two or more genes were plotted. The bar graph colours (gene counts) represent the number of genes which contributed to each GO term. For the gene-concept legend items, size refers to the number of genes that contribute to that GO term, and regression coefficient (ß) reflects the strength and direction of the association between a transcript's normative expression pattern and the thalamic nuclei volume phenotype.

#### Supplementary Table 12: Exploratory gene ontology analysis of the FTD- and neurodevelopment-related transcripts of interest.

| Ontology | ID | Description | *p* | Adjusted *p* | *q* | Contributory Genes | Gene Count |
|---|---|---|---|---|---|---|---|
| BP | GO:0021544 | subpallium development | 2.72E-08 | 2.17E-05 | 1.45E-05 | DLX2/DLX1/FOXP2/ CNTNAP2 | 4 |
| BP | GO:0021543 | pallium development | 2.48E-06 | 8.54E-04 | 5.71E-04 | DLX2/DLX1/FOXP2/ CNTNAP2/LHX2 | 5 |
| BP | GO:0001654 | eye development | 5.52E-06 | 8.54E-04 | 5.71E-04 | DLX2/NES/DLX1/SLC 17A6/SOX2/LHX2 | 6 |
| BP | GO:0150063 | visual system development | 5.83E-06 | 8.54E-04 | 5.71E-04 | DLX2/NES/DLX1/SLC 17A6/SOX2/LHX2 | 6 |
| BP | GO:0030900 | forebrain development | 5.91E-06 | 8.54E-04 | 5.71E-04 | DLX2/DLX1/FOXP2/ CNTNAP2/SOX2/LH X2 | 6 |
| BP | GO:0048880 | sensory system development | 6.42E-06 | 8.54E-04 | 5.71E-04 | DLX2/NES/DLX1/SLC 17A6/SOX2/LHX2 | 6 |
| BP | GO:0021537 | telencephalon development | 1.47E-05 | 1.67E-03 | 1.12E-03 | DLX2/DLX1/FOXP2/ CNTNAP2/LHX2 | 5 |
| BP | GO:0043010 | camera-type eye development | 4.94E-05 | 4.93E-03 | 3.30E-03 | DLX2/NES/DLX1/SLC 17A6/LHX2 | 5 |
| BP | GO:0021892 | cerebral cortex GABAergic interneuron differentiation | 7.01E-05 | 6.09E-03 | 4.08E-03 | DLX2/DLX1 | 2 |
| BP | GO:0045665 | negative regulation of neuron differentiation | 7.97E-05 | 6.09E-03 | 4.08E-03 | DLX2/DLX1/SOX2 | 3 |
| BP | GO:0035881 | amacrine cell differentiation | 8.40E-05 | 6.09E-03 | 4.08E-03 | DLX2/DLX1 | 2 |
| BP | GO:0140694 | non-membrane-bounded organelle assembly | 1.06E-04 | 7.02E-03 | 4.69E-03 | C9orf72/SQSTM1/M APT/FUS/CHMP2B | 5 |
| BP | GO:0048715 | negative regulation of oligodendrocyte differentiation | 1.16E-04 | 7.10E-03 | 4.75E-03 | DLX2/DLX1 | 2 |
| BP | GO:0010001 | glial cell differentiation | 2.29E-04 | 1.19E-02 | 7.97E-03 | DLX2/DLX1/MAPT/S OX2 | 4 |
| BP | GO:0032886 | regulation of microtubule-based process | 2.36E-04 | 1.19E-02 | 7.97E-03 | MAPT/CHMP2B/CC NF/MET | 4 |

| | | | | | | | |
|---|---|---|---|---|---|---|---|
| BP | GO:0097154 | GABAergic neuron differentiation | 2.40E-04 | 1.19E-02 | 7.97E-03 | DLX2/DLX1 | 2 |
| BP | GO:0048675 | axon extension | 2.69E-04 | 1.19E-02 | 7.97E-03 | C9orf72/MAPT/LHX2 | 3 |
| BP | GO:0021756 | striatum development | 2.92E-04 | 1.19E-02 | 7.97E-03 | FOXP2/CNTNAP2 | 2 |
| BP | GO:1902774 | late endosome to lysosome transport | 2.92E-04 | 1.19E-02 | 7.97E-03 | C9orf72/CHMP2B | 2 |
| BP | GO:0010821 | regulation of mitochondrion organization | 2.99E-04 | 1.19E-02 | 7.97E-03 | SQSTM1/MAPT/CHCHD10 | 3 |
| BP | GO:0045165 | cell fate commitment | 3.44E-04 | 1.31E-02 | 8.73E-03 | DLX2/DLX1/TCF7L2/SOX2 | 4 |
| BP | GO:0021761 | limbic system development | 4.01E-04 | 1.45E-02 | 9.72E-03 | DLX2/DLX1/CNTNAP2 | 3 |
| BP | GO:0021987 | cerebral cortex development | 4.39E-04 | 1.47E-02 | 9.83E-03 | FOXP2/CNTNAP2/LHX2 | 3 |
| BP | GO:0021895 | cerebral cortex neuron differentiation | 4.42E-04 | 1.47E-02 | 9.83E-03 | DLX2/DLX1 | 2 |
| BP | GO:0045686 | negative regulation of glial cell differentiation | 4.76E-04 | 1.52E-02 | 1.02E-02 | DLX2/DLX1 | 2 |
| BP | GO:0050768 | negative regulation of neurogenesis | 5.23E-04 | 1.60E-02 | 1.07E-02 | DLX2/DLX1/LHX2 | 3 |
| BP | GO:0051961 | negative regulation of nervous system development | 6.16E-04 | 1.82E-02 | 1.22E-02 | DLX2/DLX1/LHX2 | 3 |
| BP | GO:0031116 | positive regulation of microtubule polymerization | 6.62E-04 | 1.85E-02 | 1.24E-02 | MAPT/MET | 2 |
| BP | GO:0042063 | gliogenesis | 6.73E-04 | 1.85E-02 | 1.24E-02 | DLX2/DLX1/MAPT/SOX2 | 4 |
| BP | GO:0009954 | proximal/distal pattern formation | 7.03E-04 | 1.86E-02 | 1.24E-02 | DLX2/DLX1 | 2 |
| BP | GO:0021772 | olfactory bulb development | 7.45E-04 | 1.86E-02 | 1.24E-02 | DLX2/LHX2 | 2 |
| BP | GO:0034063 | stress granule assembly | 7.45E-04 | 1.86E-02 | 1.24E-02 | C9orf72/MAPT | 2 |
| BP | GO:0010506 | regulation of autophagy | 8.10E-04 | 1.93E-02 | 1.29E-02 | C9orf72/SQSTM1/MAPT/MET | 4 |
| BP | GO:0021988 | olfactory lobe development | 8.33E-04 | 1.93E-02 | 1.29E-02 | DLX2/LHX2 | 2 |

| | | | | | | | |
|---|---|---|---|---|---|---|---|
| BP | GO:0031112 | positive regulation of microtubule polymerization or depolymerization | 8.79E-04 | 1.93E-02 | 1.29E-02 | MAPT/MET | 2 |
| BP | GO:0050767 | regulation of neurogenesis | 8.94E-04 | 1.93E-02 | 1.29E-02 | DLX2/DLX1/MAPT/LHX2 | 4 |
| BP | GO:0070507 | regulation of microtubule cytoskeleton organization | 8.95E-04 | 1.93E-02 | 1.29E-02 | MAPT/CHMP2B/MET | 3 |
| BP | GO:1990138 | neuron projection extension | 9.42E-04 | 1.98E-02 | 1.32E-02 | C9orf72/MAPT/LHX2 | 3 |
| BP | GO:0007034 | vacuolar transport | 1.01E-03 | 2.04E-02 | 1.37E-02 | C9orf72/SQSTM1/CHMP2B | 3 |
| BP | GO:0060041 | retina development in camera-type eye | 1.02E-03 | 2.04E-02 | 1.37E-02 | DLX2/DLX1/LHX2 | 3 |
| BP | GO:0045746 | negative regulation of Notch signaling pathway | 1.12E-03 | 2.19E-02 | 1.46E-02 | DLX2/DLX1 | 2 |
| BP | GO:0014014 | negative regulation of gliogenesis | 1.23E-03 | 2.20E-02 | 1.47E-02 | DLX2/DLX1 | 2 |
| BP | GO:0021879 | forebrain neuron differentiation | 1.23E-03 | 2.20E-02 | 1.47E-02 | DLX2/DLX1 | 2 |
| BP | GO:1990000 | amyloid fibril formation | 1.23E-03 | 2.20E-02 | 1.47E-02 | MAPT/FUS | 2 |
| BP | GO:0051495 | positive regulation of cytoskeleton organization | 1.24E-03 | 2.20E-02 | 1.47E-02 | NES/MAPT/MET | 3 |
| BP | GO:0048713 | regulation of oligodendrocyte differentiation | 1.29E-03 | 2.23E-02 | 1.49E-02 | DLX2/DLX1 | 2 |
| BP | GO:0010824 | regulation of centrosome duplication | 1.34E-03 | 2.23E-02 | 1.49E-02 | CHMP2B/CCNF | 2 |
| BP | GO:1900271 | regulation of long-term synaptic potentiation | 1.34E-03 | 2.23E-02 | 1.49E-02 | SQSTM1/CALB2 | 2 |
| BP | GO:0045664 | regulation of neuron differentiation | 1.40E-03 | 2.28E-02 | 1.53E-02 | DLX2/DLX1/SOX2 | 3 |
| BP | GO:0021953 | central nervous system neuron differentiation | 1.60E-03 | 2.55E-02 | 1.70E-02 | DLX2/DLX1/MAPT | 3 |
| BP | GO:0007409 | axonogenesis | 1.75E-03 | 2.73E-02 | 1.83E-02 | C9orf72/MAPT/LHX2/S100A6 | 4 |
| BP | GO:0051960 | regulation of nervous system development | 1.80E-03 | 2.76E-02 | 1.85E-02 | DLX2/DLX1/MAPT/LHX2 | 4 |

| | | | | | | | |
|---|---|---|---|---|---|---|---|
| BP | GO:0021872 | forebrain generation of neurons | 1.83E-03 | 2.76E-02 | 1.85E-02 | DLX2/DLX1 | 2 |
| BP | GO:0046605 | regulation of centrosome cycle | 1.90E-03 | 2.80E-02 | 1.88E-02 | CHMP2B/CCNF | 2 |
| BP | GO:0031329 | regulation of cellular catabolic process | 1.93E-03 | 2.80E-02 | 1.88E-02 | C9orf72/SQSTM1/MAPT/MET | 4 |
| BP | GO:0051098 | regulation of binding | 1.98E-03 | 2.81E-02 | 1.88E-02 | NES/TCF7L2/MET | 3 |
| BP | GO:0048167 | regulation of synaptic plasticity | 2.01E-03 | 2.81E-02 | 1.88E-02 | SQSTM1/MAPT/CALB2 | 3 |
| BP | GO:0048588 | developmental cell growth | 2.34E-03 | 3.21E-02 | 2.15E-02 | C9orf72/MAPT/LHX2 | 3 |
| BP | GO:2000179 | positive regulation of neural precursor cell proliferation | 2.47E-03 | 3.34E-02 | 2.23E-02 | NES/LHX2 | 2 |
| BP | GO:0060560 | developmental growth involved in morphogenesis | 2.51E-03 | 3.34E-02 | 2.23E-02 | C9orf72/MAPT/LHX2 | 3 |
| BP | GO:0031113 | regulation of microtubule polymerization | 2.55E-03 | 3.34E-02 | 2.23E-02 | MAPT/MET | 2 |
| BP | GO:0070050 | neuron cellular homeostasis | 2.63E-03 | 3.38E-02 | 2.26E-02 | CHMP2B/CALB2 | 2 |
| BP | GO:0046530 | photoreceptor cell differentiation | 2.95E-03 | 3.74E-02 | 2.50E-02 | DLX2/DLX1 | 2 |
| BP | GO:0048255 | mRNA stabilization | 3.03E-03 | 3.78E-02 | 2.53E-02 | FUS/TAF15 | 2 |
| BP | GO:0048663 | neuron fate commitment | 3.38E-03 | 4.15E-02 | 2.77E-02 | DLX2/DLX1 | 2 |
| BP | GO:0051298 | centrosome duplication | 3.47E-03 | 4.19E-02 | 2.80E-02 | CHMP2B/CCNF | 2 |
| BP | GO:0031016 | pancreas development | 3.74E-03 | 4.37E-02 | 2.92E-02 | TCF7L2/MET | 2 |
| BP | GO:0043489 | RNA stabilization | 3.83E-03 | 4.37E-02 | 2.92E-02 | FUS/TAF15 | 2 |
| BP | GO:0045685 | regulation of glial cell differentiation | 3.83E-03 | 4.37E-02 | 2.92E-02 | DLX2/DLX1 | 2 |
| BP | GO:1902373 | negative regulation of mRNA catabolic process | 3.83E-03 | 4.37E-02 | 2.92E-02 | FUS/TAF15 | 2 |
| BP | GO:0021536 | diencephalon development | 3.93E-03 | 4.42E-02 | 2.95E-02 | CNTNAP2/SOX2 | 2 |
| BP | GO:0010721 | negative regulation of cell development | 4.01E-03 | 4.45E-02 | 2.97E-02 | DLX2/DLX1/LHX2 | 3 |
| CC | GO:0044304 | main axon | 6.88E-05 | 4.91E-03 | 3.22E-03 | C9orf72/MAPT/CNTNAP2 | 3 |
| CC | GO:0044753 | amphisome | 1.05E-04 | 4.91E-03 | 3.22E-03 | SQSTM1/CHMP2B | 2 |
| CC | GO:0030673 | axolemma | 1.21E-04 | 4.91E-03 | 3.22E-03 | MAPT/CNTNAP2 | 2 |

| | | | | | | | |
|---|---|---|---|---|---|---|---|
| CC | GO:0005776 | autophagosome | 2.86E-04 | 7.70E-03 | 5.05E-03 | C9orf72/SQSTM1/CHMP2B | 3 |
| CC | GO:0099523 | presynaptic cytosol | 3.15E-04 | 7.70E-03 | 5.05E-03 | C9orf72/FUS | 2 |
| CC | GO:0099522 | cytosolic region | 6.75E-04 | 1.37E-02 | 9.00E-03 | C9orf72/FUS | 2 |
| CC | GO:0032589 | neuron projection membrane | 2.11E-03 | 3.55E-02 | 2.32E-02 | MAPT/CNTNAP2 | 2 |
| CC | GO:0060076 | excitatory synapse | 2.68E-03 | 3.55E-02 | 2.32E-02 | SLC17A6/CALB2 | 2 |
| CC | GO:0036464 | cytoplasmic ribonucleoprotein granule | 2.74E-03 | 3.55E-02 | 2.32E-02 | C9orf72/SQSTM1/MAPT | 3 |
| CC | GO:0016234 | inclusion body | 2.99E-03 | 3.55E-02 | 2.32E-02 | SQSTM1/MAPT | 2 |
| CC | GO:0035770 | ribonucleoprotein granule | 3.35E-03 | 3.55E-02 | 2.32E-02 | C9orf72/SQSTM1/MAPT | 3 |
| CC | GO:0150034 | distal axon | 3.49E-03 | 3.55E-02 | 2.32E-02 | C9orf72/MAPT/CALB2 | 3 |

Abbreviations: BP=biological process; CC=cellular component.

### Sensitivity analyses for thalamic nuclei volumes in ALLFTD participants

Sensitivity analyses of thalamic nuclei volumes in ALLFTD participants revealed findings generally consistent with the primary analyses **(Supplementary Table 13)**. In ALLFTD, *C9orf72* repeat expansion carriers had smaller volumes in the right and left medial pulvinar (PuM): $F(1,33)=13.2$, $p=9.54e-4$, and $F(1,33)=4.28$, $p=0.046$, respectively; right anterior pulvinar (PuA): $F(1,33)=8.77$, $p=0.0056$, and left PuA trending towards significance: $F(1,33)=3.77$, $p=0.061$; right and left mediodorsal medial magnocellular nucleus (MDm): $F(1,33)=12.5$, $p=1.22e-3$, and $F(1,33)=4.34$, $p=0.045$, respectively; right and left mediodorsal lateral parvocellular nucleus (MDl): $F(1,33)=12.9$, $p=1.05e-3$ and $F(1,33)=5.59$, $p=0.024$, respectively. The left lateral posterior (LP) nucleus trended towards being smaller in young adult *C9orf72* expansion carriers in ALLFTD: $F(1,33)=3.96$, $p=0.055$. Of note, volumes of the right and left inferior pulvinar (PuI) and the left laterodorsal (LD) nuclei were significantly smaller in young adult *C9orf72* carriers in the primary analyses consisting of all FPI participants ($p=8.90e-4$, $p=0.029$, and $p=0.040$, respectively), but not significant in ALLFTD participants alone ($p>0.05$ for all).

#### Supplementary Table 13: Thalamic nuclei volume differences between young adult C9orf72 repeat expansion carriers and non-carriers in the ALLFTD cohort.

| Thalamic Nucleus | *ß* | SE | *t* | *t(-1)* | *p* |
|---|---|---|---|---|---|
| Left AV | -2.36 | 8.53 | -0.28 | 0.28 | 0.783 |
| Right AV | -0.31 | 7.81 | -0.04 | 0.04 | 0.968 |
| Left LD | -2.53 | 2.75 | -0.92 | 0.92 | 0.365 |
| Right LD | -2.81 | 3.01 | -0.93 | 0.93 | 0.357 |
| Left LP | -11.64 | 5.85 | -1.99 | 1.99 | 0.055† |
| Right LP | -9.95 | 5.84 | -1.70 | 1.70 | 0.098† |
| Left VA | -8.99 | 14.08 | -0.64 | 0.64 | 0.528 |
| Right VA | -13.00 | 14.76 | -0.88 | 0.88 | 0.385 |
| Left VAmc | -0.27 | 1.17 | -0.23 | 0.23 | 0.816 |
| Right VAmc | -0.81 | 1.14 | -0.71 | 0.71 | 0.483 |
| Left VLa | -18.71 | 18.15 | -1.03 | 1.03 | 0.310 |
| Right VLa | -23.90 | 18.43 | -1.30 | 1.30 | 0.204 |
| Left VLp | -20.80 | 24.49 | -0.85 | 0.85 | 0.402 |
| Right VLp | -32.02 | 24.37 | -1.31 | 1.31 | 0.198 |
| Left VPL | -8.02 | 35.50 | -0.23 | 0.23 | 0.823 |
| Right VPL | -30.59 | 34.04 | -0.90 | 0.90 | 0.375 |
| Left CeM | -0.72 | 3.01 | -0.24 | 0.24 | 0.812 |
| Right CeM | -1.86 | 3.16 | -0.59 | 0.59 | 0.559 |
| Left CL | 1.46 | 2.74 | 0.53 | -0.53 | 0.598 |
| Right CL | 2.32 | 2.86 | 0.81 | -0.81 | 0.422 |
| Left CM | 3.17 | 7.77 | 0.41 | -0.41 | 0.686 |
| Right CM | -1.18 | 8.29 | -0.14 | 0.14 | 0.888 |
| Left Pf | 1.66 | 2.39 | 0.70 | -0.70 | 0.491 |

| | | | | | |
|---|---|---|---|---|---|
| Right Pf | -0.31 | 2.04 | -0.15 | 0.15 | 0.881 |
| Left MVRe | -0.63 | 0.87 | -0.72 | 0.72 | 0.478 |
| Right MVRe | -0.23 | 0.93 | -0.24 | 0.24 | 0.808 |
| **Left MDm** | **-59.49** | **28.54** | **-2.08** | **2.08** | **0.045*** |
| **Right MDm** | **-95.82** | **27.09** | **-3.54** | **3.54** | **0.0012**** |
| **Left MDl** | **-25.45** | **10.76** | **-2.36** | **2.36** | **0.024**** |
| **Right MDl** | **-37.89** | **10.55** | **-3.59** | **3.59** | **0.0011**** |
| Left LGN | -1.43 | 13.12 | -0.11 | 0.11 | 0.914 |
| Right LGN | -6.07 | 8.79 | -0.69 | 0.69 | 0.494 |
| Left MGN | -2.72 | 5.25 | -0.52 | 0.52 | 0.607 |
| Right MGN | -6.19 | 5.99 | -1.03 | 1.03 | 0.309 |
| Left LSg | -1.97 | 2.00 | -0.98 | 0.98 | 0.333 |
| Right LSg | -0.12 | 1.96 | -0.06 | 0.06 | 0.950 |
| Left PuA | -18.06 | 9.30 | -1.94 | 1.94 | 0.061† |
| **Right PuA** | **-26.12** | **8.82** | **-2.96** | **2.96** | **0.0056**** |
| **Left PuM** | **-93.23** | **45.07** | **-2.07** | **2.07** | **0.046*** |
| **Right PuM** | **-133.18** | **36.71** | **-3.63** | **3.63** | **0.00095***** |
| Left PuL | -14.95 | 9.57 | -1.56 | 1.56 | 0.128 |
| Right PuL | -5.38 | 8.66 | -0.62 | 0.62 | 0.539 |
| Left PuI | -8.72 | 14.23 | -0.61 | 0.61 | 0.544 |
| Right PuI | -11.80 | 10.56 | -1.12 | 1.12 | 0.272 |

Only the main effect of group is shown for each thalamic nuclei model. A negative t-statistic reflects a smaller volume in the carrier group, and vice versa. To ease interpretation of downstream correlations with transcriptomic expression, imaging model t-statistics were multiplied by -1, such that larger values corresponded to larger effects of smaller nucleus volume in the carrier group. Thalamic nuclei with significantly smaller volumes in *C9orf72* repeat expansion carriers are bolded. *$p$<0.05, **$p$<0.01, ***$p$<0.001. †$p$<0.10 (trending towards significance). Abbreviations: AV=anteroventral; LD=laterodorsal; LP=lateral posterior; VA=ventral anterior; VAmc=ventral anterior magnocellular; VLa=ventral lateral anterior; VLp=ventral lateral posterior; VPL=ventral posterolateral; CeM=central medial; CL=central lateral; CM=centromedian; Pf=parafascicular; MVRe=reuniens (medial ventral); MDm=mediodorsal medial magnocellular; MDl=mediodorsal lateral parvocellular; LGN=lateral geniculate; MGN=medial geniculate; LSg=limitans (suprageniculate); PuA=pulvinar anterior; PuM=pulvinar medial; PuL=pulvinar lateral; PuI=pulvinar inferior.

### Sensitivity analyses for normative spatial transcriptomic data

Sensitivity analyses were computed to better delineate sex- and age-related findings without removing the single female AHBA donor brain from the primary analysis. This included examination of each individual donor separately, as well as the two younger donors in aggregate, and the remaining older donors in aggregate, to verify the expression patterns of the top 100 data-driven transcripts derived from the NMF of the primary analyses, and of the 37 hypothesis-driven transcripts of interest. For sensitivity analyses of the individual donors, when multiple probes indexed the expression of the same gene, we selected and used the probe with the highest mean intensity across samples instead of applying the differential stability calculation meant for selecting a probe across multiple donors. Microarray data from the late-fifties male donor were missing these seven transcripts: *ABCA8, FLNC, LOC100507433, ADAMTS14, ENPP6, HSD17B3, TSPAN8*.

Similarly, microarray data from the early-20s male donor were missing these nine transcripts: *FLNC, LOC100507487, ADAMTS14, LOC101929384, HSD17B3, TSPAN8, RGS1, DMBT1, GALNT6*. Therefore, seven transcripts were not available in the sensitivity analysis which combined the two younger donors: *FLNC, LOC100507487, ADAMTS14, LOC101929384, HSD17B3, RGS1, GALNT6*.

Sensitivity analyses of normative spatial expression of *C9orf72* and the top 100 data-driven transcripts in individual donors generally did not yield adjusted *p*-values that survived correction for multiple comparisons, except for in the late-30s male and sole female **(Supplementary Table 14).** The directions of the regression coefficients (*ß*) were generally consistent across all donors, although effect magnitudes differed depending on the donor, where the youngest and oldest male donors showed relatively smaller *ß*, the remaining male donors showed relatively similar *ß*, and the sole female donor showed relatively larger *ß* for the top 100 transcripts. This suggests a generally reproducible spatial normative expression pattern of the top 100 transcripts, though with inter-individual variability also present, likely due to demographic differences in age and sex, as well as differences in sampling density and measurement noise across donors. When sensitivity analyses were computed in aggregate (the two younger donors in aggregate and four older donors in aggregate), similar t-values were observed, with relatively larger *ß* observed in the aggregate of four donors.

Sensitivity analyses of the hypothesis-driven transcripts demonstrated similar findings of consistent directions of regression coefficients per transcript, though individual *p*-values did not survive correction for multiple comparisons, except for in the late-30s male and sole female **(Supplementary Table 15).** When sensitivity analyses were computed in aggregate (the two younger donors in aggregate and four older donors in aggregate), similar t-values were observed, with relatively larger *ß* observed in the aggregate of four donors. In the analysis of the two younger donors in aggregate, none of the transcripts survived corrections, though *CALB1* (which remained significant after correction in the primary analyses) appeared as a top 5 transcript whose spatial normative expression was most associated with thalamic nuclei volumes. In the analysis of the four older donors in aggregate, *CALB1* was the sole transcript which remained significant after correction, consistent with the primary analyses.

#### Supplementary Table 14a: Sensitivity analyses comprising Limma linear model statistics for normative spatial expression of *C9orf72* and the top 100 data-driven transcripts in the early-20s male donor.

| **Transcript** | ***ß*** | ***t*** | ***p*** | **Adjusted *p*** |
|---|---|---|---|---|
| *ARRDC2* | 0.052 | 2.49 | 1.64E-02 | 4.02E-01 |
| *ENPP6* | 0.041 | 2.25 | 2.91E-02 | 4.02E-01 |
| *PRKCQ* | 0.047 | 2.17 | 3.48E-02 | 4.02E-01 |
| *KIAA1147* | 0.035 | 2.14 | 3.77E-02 | 4.02E-01 |
| *RAB7B* | 0.041 | 2.09 | 4.20E-02 | 4.02E-01 |
| *MYLK* | 0.044 | 1.96 | 5.62E-02 | 4.02E-01 |
| *ADAMTS4* | 0.025 | 1.95 | 5.73E-02 | 4.02E-01 |
| *NDE1* | 0.023 | 1.94 | 5.85E-02 | 4.02E-01 |
| *JPT2* | 0.025 | 1.91 | 6.26E-02 | 4.02E-01 |
| *CTNNA3* | 0.030 | 1.87 | 6.79E-02 | 4.02E-01 |
| *USH1C* | 0.035 | 1.86 | 6.97E-02 | 4.02E-01 |
| *ZBED3* | 0.019 | 1.85 | 7.02E-02 | 4.02E-01 |
| *HIPK2* | 0.026 | 1.85 | 7.11E-02 | 4.02E-01 |
| *SAP30* | 0.037 | 1.81 | 7.70E-02 | 4.02E-01 |
| *ABCA8* | 0.031 | 1.74 | 8.86E-02 | 4.02E-01 |

| | | | | |
|---|---|---|---|---|
| *ARHGEF37* | 0.024 | 1.73 | 8.91E-02 | 4.02E-01 |
| *MAG* | 0.019 | 1.68 | 9.96E-02 | 4.02E-01 |
| *IGFLR1* | 0.028 | 1.67 | 1.02E-01 | 4.02E-01 |
| *MOBP* | 0.020 | 1.66 | 1.02E-01 | 4.02E-01 |
| *CLDND1* | 0.022 | 1.66 | 1.04E-01 | 4.02E-01 |
| *SSFA2* | 0.025 | 1.64 | 1.07E-01 | 4.02E-01 |
| *CMTM5* | 0.030 | 1.63 | 1.10E-01 | 4.02E-01 |
| *RASSF2* | 0.018 | 1.62 | 1.11E-01 | 4.02E-01 |
| *TMEM98* | 0.022 | 1.61 | 1.14E-01 | 4.02E-01 |
| *IFIT2* | 0.023 | 1.60 | 1.16E-01 | 4.02E-01 |
| *SLC31A2* | 0.026 | 1.58 | 1.20E-01 | 4.02E-01 |
| *FAM149B1* | 0.020 | 1.57 | 1.23E-01 | 4.02E-01 |
| *CARNS1* | 0.024 | 1.53 | 1.32E-01 | 4.02E-01 |
| *SLC5A11* | 0.019 | 1.52 | 1.35E-01 | 4.02E-01 |
| *PAIP2B* | 0.018 | 1.52 | 1.36E-01 | 4.02E-01 |
| *TMTC2* | 0.021 | 1.50 | 1.39E-01 | 4.02E-01 |
| *SELENOP* | 0.019 | 1.50 | 1.40E-01 | 4.02E-01 |
| *PHF10* | 0.022 | 1.41 | 1.66E-01 | 4.45E-01 |
| *CAPN3* | 0.015 | 1.38 | 1.74E-01 | 4.45E-01 |
| *FNTA* | 0.026 | 1.35 | 1.83E-01 | 4.45E-01 |
| *KATNA1* | 0.023 | 1.34 | 1.85E-01 | 4.45E-01 |
| *USP54* | 0.024 | 1.34 | 1.87E-01 | 4.45E-01 |
| *TMTC4* | 0.028 | 1.34 | 1.87E-01 | 4.45E-01 |
| *FAM19A4* | -0.013 | -1.32 | 1.93E-01 | 4.45E-01 |
| *LOC100506725* | 0.016 | 1.32 | 1.94E-01 | 4.45E-01 |
| *RDH11* | 0.020 | 1.30 | 1.98E-01 | 4.45E-01 |
| *CHST6* | 0.012 | 1.29 | 2.04E-01 | 4.46E-01 |
| *TCFL5* | 0.021 | 1.20 | 2.35E-01 | 4.92E-01 |
| *SMARCC1* | 0.018 | 1.19 | 2.39E-01 | 4.92E-01 |
| *ARHGEF1* | 0.030 | 1.18 | 2.44E-01 | 4.92E-01 |
| *S100B* | 0.013 | 1.17 | 2.46E-01 | 4.92E-01 |
| *C9orf3* | 0.017 | 1.14 | 2.59E-01 | 5.05E-01 |
| *NUDT1* | 0.028 | 1.13 | 2.63E-01 | 5.05E-01 |
| *ANKRD65* | 0.019 | 1.07 | 2.88E-01 | 5.23E-01 |
| *FEZ1* | 0.026 | 1.06 | 2.93E-01 | 5.23E-01 |
| *TMEM140* | -0.020 | -1.06 | 2.95E-01 | 5.23E-01 |
| *CTDSP1* | 0.009 | 1.06 | 2.97E-01 | 5.23E-01 |
| *ADGRE5* | 0.014 | 1.04 | 3.01E-01 | 5.23E-01 |
| *PLEKHB1* | 0.012 | 1.02 | 3.11E-01 | 5.29E-01 |
| *PPIF* | 0.016 | 1.00 | 3.23E-01 | 5.40E-01 |
| *SCRG1* | 0.026 | 0.97 | 3.37E-01 | 5.54E-01 |
| *PCBP4* | 0.019 | 0.95 | 3.47E-01 | 5.58E-01 |
| *QDPR* | 0.013 | 0.94 | 3.52E-01 | 5.58E-01 |

| | | | | |
|---|---|---|---|---|
| *PCSK6* | 0.015 | 0.90 | 3.70E-01 | 5.78E-01 |
| *ZNF664* | 0.010 | 0.89 | 3.77E-01 | 5.79E-01 |
| *LLGL2* | 0.015 | 0.87 | 3.86E-01 | 5.82E-01 |
| *CRTAP* | 0.014 | 0.85 | 4.00E-01 | 5.89E-01 |
| *LRP6* | 0.012 | 0.84 | 4.07E-01 | 5.89E-01 |
| *BAMBI* | 0.009 | 0.82 | 4.14E-01 | 5.89E-01 |
| *ASPRV1* | 0.012 | 0.81 | 4.22E-01 | 5.89E-01 |
| *LDLRAD4* | 0.013 | 0.81 | 4.23E-01 | 5.89E-01 |
| *TNS2* | 0.011 | 0.78 | 4.39E-01 | 6.03E-01 |
| *FAT1* | 0.005 | 0.75 | 4.54E-01 | 6.08E-01 |
| *VKORC1* | 0.015 | 0.75 | 4.56E-01 | 6.08E-01 |
| *KHNYN* | 0.013 | 0.74 | 4.62E-01 | 6.08E-01 |
| *RCOR2* | -0.009 | -0.65 | 5.19E-01 | 6.72E-01 |
| *SESTD1* | -0.006 | -0.64 | 5.26E-01 | 6.72E-01 |
| *LAP3* | 0.009 | 0.53 | 5.96E-01 | 7.50E-01 |
| *SEPT4* | 0.006 | 0.52 | 6.03E-01 | 7.50E-01 |
| *TST* | 0.010 | 0.49 | 6.29E-01 | 7.72E-01 |
| *GPRC5B* | 0.008 | 0.45 | 6.55E-01 | 7.74E-01 |
| *CDH20* | 0.007 | 0.45 | 6.58E-01 | 7.74E-01 |
| *C9orf72* | -0.008 | -0.44 | 6.63E-01 | 7.74E-01 |
| *FRMD5* | -0.008 | -0.43 | 6.68E-01 | 7.74E-01 |
| *BCAR3* | 0.008 | 0.41 | 6.82E-01 | 7.74E-01 |
| *ADARB2.AS1* | 0.009 | 0.40 | 6.91E-01 | 7.74E-01 |
| *PARP4* | 0.007 | 0.39 | 6.96E-01 | 7.74E-01 |
| *HHATL* | 0.007 | 0.39 | 6.98E-01 | 7.74E-01 |
| *B3GNT9* | 0.008 | 0.36 | 7.18E-01 | 7.79E-01 |
| *TSPAN15* | 0.006 | 0.36 | 7.19E-01 | 7.79E-01 |
| *AAMDC* | 0.006 | 0.32 | 7.51E-01 | 8.03E-01 |
| *KLC1* | -0.003 | -0.24 | 8.13E-01 | 8.60E-01 |
| *ARHGAP21* | -0.002 | -0.14 | 8.90E-01 | 9.30E-01 |
| *DENND5A* | 0.001 | 0.12 | 9.07E-01 | 9.38E-01 |
| *BCDIN3D* | -0.002 | -0.09 | 9.26E-01 | 9.46E-01 |
| *ICK* | -0.0003 | -0.02 | 9.87E-01 | 9.89E-01 |
| *SFR1* | 0.0003 | 0.01 | 9.89E-01 | 9.89E-01 |

#### Supplementary Table 14b: Sensitivity analyses comprising Limma linear model statistics for normative spatial expression of *C9orf72* and the top 100 data-driven transcripts in the early-30s male donor.

| **Transcript** | **ß** | ***t*** | ***p*** | **Adjusted *p*** |
|---|---|---|---|---|
| *SSFA2* | 0.129 | 2.82 | 7.29E-03 | 2.71E-01 |
| *ZNF664* | 0.119 | 2.75 | 8.78E-03 | 2.71E-01 |
| *VKORC1* | 0.118 | 2.59 | 1.34E-02 | 2.71E-01 |

| | | | | |
|---|---|---|---|---|
| *IGFLR1* | 0.104 | 2.41 | 2.04E-02 | 2.71E-01 |
| *ASPRV1* | 0.097 | 2.36 | 2.31E-02 | 2.71E-01 |
| *PAIP2B* | 0.083 | 2.24 | 3.07E-02 | 2.71E-01 |
| *BAMBI* | 0.086 | 2.21 | 3.26E-02 | 2.71E-01 |
| *ZBED3* | 0.085 | 2.14 | 3.81E-02 | 2.71E-01 |
| *SCRG1* | 0.087 | 2.09 | 4.25E-02 | 2.71E-01 |
| *ARRDC2* | 0.086 | 2.08 | 4.43E-02 | 2.71E-01 |
| *CRTAP* | 0.074 | 2.05 | 4.67E-02 | 2.71E-01 |
| *RAB7B* | 0.083 | 2.00 | 5.27E-02 | 2.71E-01 |
| *ANKRD65* | 0.072 | 1.95 | 5.76E-02 | 2.71E-01 |
| *CHST6* | 0.073 | 1.95 | 5.78E-02 | 2.71E-01 |
| *SMARCC1* | 0.071 | 1.92 | 6.14E-02 | 2.71E-01 |
| *FAM149B1* | 0.080 | 1.92 | 6.20E-02 | 2.71E-01 |
| *ADGRE5* | 0.079 | 1.92 | 6.22E-02 | 2.71E-01 |
| *ARHGEF37* | 0.073 | 1.91 | 6.34E-02 | 2.71E-01 |
| *B3GNT9* | 0.067 | 1.86 | 7.00E-02 | 2.71E-01 |
| *FAT1* | 0.070 | 1.86 | 7.00E-02 | 2.71E-01 |
| *CTDSP1* | 0.070 | 1.85 | 7.14E-02 | 2.71E-01 |
| *TMTC2* | 0.077 | 1.85 | 7.21E-02 | 2.71E-01 |
| *CDH20* | 0.062 | 1.83 | 7.48E-02 | 2.71E-01 |
| *IFIT2* | 0.075 | 1.81 | 7.71E-02 | 2.71E-01 |
| *MYLK* | 0.063 | 1.79 | 8.09E-02 | 2.71E-01 |
| *BCAR3* | 0.090 | 1.78 | 8.21E-02 | 2.71E-01 |
| *MOBP* | 0.060 | 1.78 | 8.26E-02 | 2.71E-01 |
| *LRP6* | 0.073 | 1.77 | 8.37E-02 | 2.71E-01 |
| *PLEKHB1* | 0.067 | 1.77 | 8.38E-02 | 2.71E-01 |
| *SAP30* | 0.073 | 1.77 | 8.47E-02 | 2.71E-01 |
| *TCFL5* | 0.067 | 1.76 | 8.55E-02 | 2.71E-01 |
| *S100B* | 0.069 | 1.76 | 8.58E-02 | 2.71E-01 |
| *CMTM5* | 0.067 | 1.70 | 9.65E-02 | 2.76E-01 |
| *TNS2* | 0.062 | 1.68 | 1.00E-01 | 2.76E-01 |
| *RASSF2* | 0.062 | 1.68 | 1.00E-01 | 2.76E-01 |
| *FAM19A4* | -0.075 | -1.67 | 1.02E-01 | 2.76E-01 |
| *KHNYN* | 0.065 | 1.66 | 1.04E-01 | 2.76E-01 |
| *HIPK2* | 0.057 | 1.65 | 1.06E-01 | 2.76E-01 |
| *SESTD1* | -0.082 | -1.65 | 1.07E-01 | 2.76E-01 |
| *FNTA* | 0.061 | 1.64 | 1.09E-01 | 2.76E-01 |
| *USP54* | 0.060 | 1.62 | 1.12E-01 | 2.76E-01 |
| *PCBP4* | 0.055 | 1.58 | 1.21E-01 | 2.92E-01 |
| *BCDIN3D* | 0.060 | 1.56 | 1.26E-01 | 2.96E-01 |
| *TSPAN8* | 0.064 | 1.51 | 1.39E-01 | 3.12E-01 |
| *CLDND1* | 0.058 | 1.49 | 1.43E-01 | 3.12E-01 |
| *NDE1* | 0.055 | 1.49 | 1.44E-01 | 3.12E-01 |

| | | | | |
|---|---|---|---|---|
| *USH1C* | 0.066 | 1.48 | 1.45E-01 | 3.12E-01 |
| *AAMDC* | 0.054 | 1.46 | 1.51E-01 | 3.18E-01 |
| *CTNNA3* | 0.052 | 1.41 | 1.67E-01 | 3.26E-01 |
| *FEZ1* | 0.057 | 1.41 | 1.67E-01 | 3.26E-01 |
| *JPT2* | 0.061 | 1.40 | 1.69E-01 | 3.26E-01 |
| *ARHGEF1* | 0.048 | 1.40 | 1.70E-01 | 3.26E-01 |
| *LOC101929384* | 0.067 | 1.39 | 1.71E-01 | 3.26E-01 |
| *SELENOP* | 0.053 | 1.35 | 1.85E-01 | 3.43E-01 |
| *QDPR* | 0.049 | 1.34 | 1.87E-01 | 3.43E-01 |
| *HHATL* | 0.053 | 1.29 | 2.04E-01 | 3.67E-01 |
| *ICK* | -0.054 | -1.27 | 2.11E-01 | 3.70E-01 |
| *DMBT1* | 0.052 | 1.25 | 2.17E-01 | 3.70E-01 |
| *SLC31A2* | 0.050 | 1.25 | 2.19E-01 | 3.70E-01 |
| *KATNA1* | 0.049 | 1.24 | 2.21E-01 | 3.70E-01 |
| *MAG* | 0.048 | 1.22 | 2.31E-01 | 3.70E-01 |
| *TMTC4* | 0.046 | 1.21 | 2.32E-01 | 3.70E-01 |
| *CAPN3* | 0.045 | 1.21 | 2.33E-01 | 3.70E-01 |
| *CARNS1* | 0.046 | 1.20 | 2.37E-01 | 3.70E-01 |
| *RDH11* | 0.045 | 1.20 | 2.38E-01 | 3.70E-01 |
| *KLC1* | -0.039 | -1.14 | 2.61E-01 | 3.94E-01 |
| *ADAMTS4* | 0.043 | 1.12 | 2.68E-01 | 3.94E-01 |
| *ADAMTS14* | 0.049 | 1.12 | 2.69E-01 | 3.94E-01 |
| *ABCA8* | 0.042 | 1.12 | 2.69E-01 | 3.94E-01 |
| *SLC5A11* | 0.039 | 1.10 | 2.79E-01 | 3.98E-01 |
| *KIAA1147* | 0.045 | 1.08 | 2.85E-01 | 3.98E-01 |
| *LLGL2* | 0.042 | 1.08 | 2.87E-01 | 3.98E-01 |
| *PARP4* | 0.044 | 1.07 | 2.90E-01 | 3.98E-01 |
| *GPRC5B* | 0.039 | 1.07 | 2.92E-01 | 3.98E-01 |
| *PHF10* | 0.042 | 1.04 | 3.04E-01 | 4.10E-01 |
| *TMEM98* | 0.041 | 1.02 | 3.12E-01 | 4.15E-01 |
| *LOC100507487* | 0.052 | 1.01 | 3.20E-01 | 4.19E-01 |
| *HSD17B3* | 0.039 | 0.99 | 3.27E-01 | 4.24E-01 |
| *ENPP6* | 0.036 | 0.96 | 3.42E-01 | 4.37E-01 |
| *C9orf3* | 0.039 | 0.92 | 3.63E-01 | 4.58E-01 |
| *NUDT1* | 0.042 | 0.88 | 3.82E-01 | 4.76E-01 |
| *FLNC* | 0.032 | 0.86 | 3.94E-01 | 4.77E-01 |
| *GALNT6* | 0.033 | 0.86 | 3.95E-01 | 4.77E-01 |
| *PPIF* | 0.032 | 0.86 | 3.96E-01 | 4.77E-01 |
| *DENND5A* | 0.030 | 0.84 | 4.08E-01 | 4.81E-01 |
| *TSPAN15* | 0.036 | 0.83 | 4.10E-01 | 4.81E-01 |
| *PRKCQ* | 0.027 | 0.81 | 4.20E-01 | 4.88E-01 |
| *TST* | 0.030 | 0.79 | 4.33E-01 | 4.97E-01 |
| *ARHGAP21* | 0.033 | 0.77 | 4.47E-01 | 5.08E-01 |

| | | | | |
|---|---|---|---|---|
| *SFR1* | 0.029 | 0.71 | 4.84E-01 | 5.43E-01 |
| *TMEM140* | 0.028 | 0.70 | 4.90E-01 | 5.44E-01 |
| *LOC100506725* | 0.024 | 0.58 | 5.65E-01 | 6.20E-01 |
| *SEPT4* | 0.017 | 0.55 | 5.86E-01 | 6.34E-01 |
| *RGS1* | 0.016 | 0.54 | 5.90E-01 | 6.34E-01 |
| *FRMD5* | 0.019 | 0.51 | 6.13E-01 | 6.52E-01 |
| *C9orf72* | 0.015 | 0.49 | 6.27E-01 | 6.60E-01 |
| *ADARB2.AS1* | -0.017 | -0.43 | 6.72E-01 | 7.00E-01 |
| *PCSK6* | 0.013 | 0.36 | 7.21E-01 | 7.43E-01 |
| *RCOR2* | -0.010 | -0.33 | 7.45E-01 | 7.60E-01 |
| *LDLRAD4* | 0.008 | 0.25 | 8.04E-01 | 8.12E-01 |
| *LAP3* | 0.007 | 0.18 | 8.61E-01 | 8.61E-01 |

#### Supplementary Table 14c: Sensitivity analyses comprising Limma linear model statistics for normative spatial expression of *C9orf72* and the top 100 data-driven transcripts in the late-30s male donor.

| **Transcript** | ***ß*** | ***t*** | ***p*** | **Adjusted *p*** |
|---|---|---|---|---|
| *CRTAP* | 0.120 | 3.84 | 2.34E-04 | 1.07E-02 |
| *SSFA2* | 0.103 | 3.60 | 5.22E-04 | 1.07E-02 |
| *TSPAN8* | 0.110 | 3.56 | 6.08E-04 | 1.07E-02 |
| *USH1C* | 0.102 | 3.55 | 6.14E-04 | 1.07E-02 |
| *SCRG1* | 0.105 | 3.54 | 6.46E-04 | 1.07E-02 |
| *ASPRV1* | 0.107 | 3.51 | 7.12E-04 | 1.07E-02 |
| *ZBED3* | 0.108 | 3.49 | 7.60E-04 | 1.07E-02 |
| *RAB7B* | 0.102 | 3.41 | 9.79E-04 | 1.07E-02 |
| *CTDSP1* | 0.103 | 3.38 | 1.08E-03 | 1.07E-02 |
| *HHATL* | 0.103 | 3.30 | 1.42E-03 | 1.07E-02 |
| *GALNT6* | 0.094 | 3.29 | 1.46E-03 | 1.07E-02 |
| *SMARCC1* | 0.101 | 3.29 | 1.46E-03 | 1.07E-02 |
| *USP54* | 0.099 | 3.27 | 1.53E-03 | 1.07E-02 |
| *PCBP4* | 0.096 | 3.27 | 1.55E-03 | 1.07E-02 |
| *PRKCQ* | 0.091 | 3.22 | 1.78E-03 | 1.07E-02 |
| *PHF10* | 0.083 | 3.22 | 1.81E-03 | 1.07E-02 |
| *DMBT1* | 0.094 | 3.20 | 1.93E-03 | 1.07E-02 |
| *ZNF664* | 0.097 | 3.19 | 1.97E-03 | 1.07E-02 |
| *JPT2* | 0.093 | 3.18 | 2.02E-03 | 1.07E-02 |
| *KIAA1147* | 0.094 | 3.12 | 2.44E-03 | 1.17E-02 |
| *MYLK* | 0.095 | 3.11 | 2.54E-03 | 1.17E-02 |
| *BAMBI* | 0.089 | 3.11 | 2.54E-03 | 1.17E-02 |
| *PAIP2B* | 0.098 | 3.09 | 2.72E-03 | 1.19E-02 |
| *ADGRE5* | 0.093 | 3.04 | 3.14E-03 | 1.23E-02 |
| *CHST6* | 0.087 | 3.04 | 3.14E-03 | 1.23E-02 |

| | | | | |
|---|---|---|---|---|
| *LOC101929384* | 0.087 | 3.03 | 3.21E-03 | 1.23E-02 |
| *ARRDC2* | 0.090 | 3.02 | 3.28E-03 | 1.23E-02 |
| *ARHGEF37* | 0.089 | 2.99 | 3.65E-03 | 1.27E-02 |
| *ABCA8* | 0.090 | 2.99 | 3.65E-03 | 1.27E-02 |
| *CDH20* | 0.086 | 2.98 | 3.76E-03 | 1.27E-02 |
| *TSPAN15* | 0.087 | 2.93 | 4.32E-03 | 1.31E-02 |
| *FLNC* | 0.089 | 2.92 | 4.51E-03 | 1.31E-02 |
| *TMTC2* | 0.088 | 2.90 | 4.70E-03 | 1.31E-02 |
| *HSD17B3* | 0.084 | 2.90 | 4.78E-03 | 1.31E-02 |
| *NDE1* | 0.089 | 2.89 | 4.81E-03 | 1.31E-02 |
| *FAT1* | 0.081 | 2.89 | 4.82E-03 | 1.31E-02 |
| *IFIT2* | 0.082 | 2.89 | 4.91E-03 | 1.31E-02 |
| *B3GNT9* | 0.081 | 2.88 | 4.94E-03 | 1.31E-02 |
| *CLDND1* | 0.088 | 2.86 | 5.28E-03 | 1.37E-02 |
| *LRP6* | 0.081 | 2.82 | 5.88E-03 | 1.46E-02 |
| *FAM149B1* | 0.080 | 2.82 | 5.93E-03 | 1.46E-02 |
| *RASSF2* | 0.083 | 2.80 | 6.26E-03 | 1.48E-02 |
| *CTNNA3* | 0.080 | 2.79 | 6.54E-03 | 1.48E-02 |
| *IGFLR1* | 0.076 | 2.78 | 6.75E-03 | 1.48E-02 |
| *FEZ1* | 0.086 | 2.76 | 6.95E-03 | 1.48E-02 |
| *PCSK6* | 0.079 | 2.76 | 7.00E-03 | 1.48E-02 |
| *VKORC1* | 0.090 | 2.75 | 7.20E-03 | 1.48E-02 |
| *GPRC5B* | 0.086 | 2.75 | 7.24E-03 | 1.48E-02 |
| *NUDT1* | 0.085 | 2.75 | 7.33E-03 | 1.48E-02 |
| *ARHGAP21* | 0.083 | 2.74 | 7.37E-03 | 1.48E-02 |
| *CARNS1* | 0.082 | 2.74 | 7.50E-03 | 1.48E-02 |
| *MAG* | 0.082 | 2.73 | 7.70E-03 | 1.50E-02 |
| *KATNA1* | 0.072 | 2.72 | 7.87E-03 | 1.50E-02 |
| *S100B* | 0.081 | 2.69 | 8.64E-03 | 1.60E-02 |
| *CMTM5* | 0.078 | 2.68 | 8.79E-03 | 1.60E-02 |
| *TNS2* | 0.085 | 2.68 | 8.87E-03 | 1.60E-02 |
| *SLC5A11* | 0.081 | 2.67 | 9.15E-03 | 1.62E-02 |
| *ENPP6* | 0.071 | 2.53 | 1.31E-02 | 2.27E-02 |
| *ADAMTS4* | 0.076 | 2.52 | 1.34E-02 | 2.29E-02 |
| *MOBP* | 0.076 | 2.52 | 1.37E-02 | 2.31E-02 |
| *ARHGEF1* | 0.071 | 2.51 | 1.40E-02 | 2.32E-02 |
| *PARP4* | 0.077 | 2.50 | 1.44E-02 | 2.35E-02 |
| *AAMDC* | 0.075 | 2.45 | 1.62E-02 | 2.59E-02 |
| *TMEM98* | 0.072 | 2.43 | 1.71E-02 | 2.69E-02 |
| *SFR1* | 0.065 | 2.42 | 1.77E-02 | 2.69E-02 |
| *SLC31A2* | 0.070 | 2.42 | 1.77E-02 | 2.69E-02 |
| *FNTA* | 0.073 | 2.41 | 1.80E-02 | 2.69E-02 |
| *KHNYN* | 0.070 | 2.40 | 1.83E-02 | 2.69E-02 |

| | | | | |
|---|---|---|---|---|
| *SAP30* | 0.072 | 2.40 | 1.84E-02 | 2.69E-02 |
| *SELENOP* | 0.067 | 2.39 | 1.91E-02 | 2.75E-02 |
| *HIPK2* | 0.064 | 2.38 | 1.97E-02 | 2.80E-02 |
| *KLC1* | -0.063 | -2.35 | 2.10E-02 | 2.95E-02 |
| *ADAMTS14* | 0.058 | 2.33 | 2.21E-02 | 3.05E-02 |
| *TST* | 0.070 | 2.32 | 2.25E-02 | 3.06E-02 |
| *DENND5A* | 0.074 | 2.32 | 2.27E-02 | 3.06E-02 |
| *CAPN3* | 0.068 | 2.29 | 2.47E-02 | 3.26E-02 |
| *PLEKHB1* | 0.071 | 2.28 | 2.49E-02 | 3.26E-02 |
| *LOC100506725* | 0.065 | 2.21 | 2.94E-02 | 3.81E-02 |
| *SEPT4* | 0.064 | 2.13 | 3.58E-02 | 4.58E-02 |
| *TCFL5* | 0.064 | 2.11 | 3.74E-02 | 4.72E-02 |
| *QDPR* | 0.063 | 2.05 | 4.38E-02 | 5.45E-02 |
| *LOC100507487* | 0.062 | 2.04 | 4.42E-02 | 5.45E-02 |
| *PPIF* | 0.061 | 2.03 | 4.50E-02 | 5.47E-02 |
| *TMTC4* | 0.054 | 1.98 | 5.12E-02 | 6.15E-02 |
| *BCAR3* | 0.055 | 1.84 | 6.91E-02 | 8.21E-02 |
| *TMEM140* | 0.049 | 1.83 | 7.12E-02 | 8.36E-02 |
| *FRMD5* | 0.048 | 1.62 | 1.09E-01 | 1.26E-01 |
| *RDH11* | 0.046 | 1.57 | 1.21E-01 | 1.39E-01 |
| *RGS1* | 0.039 | 1.31 | 1.94E-01 | 2.20E-01 |
| *LLGL2* | 0.032 | 1.26 | 2.13E-01 | 2.39E-01 |
| *C9orf72* | 0.028 | 1.11 | 2.68E-01 | 2.98E-01 |
| *ANKRD65* | 0.030 | 1.07 | 2.88E-01 | 3.16E-01 |
| *C9orf3* | 0.025 | 1.05 | 2.95E-01 | 3.20E-01 |
| *LDLRAD4* | 0.023 | 0.95 | 3.44E-01 | 3.69E-01 |
| *RCOR2* | 0.022 | 0.75 | 4.55E-01 | 4.84E-01 |
| *ICK* | 0.013 | 0.48 | 6.29E-01 | 6.62E-01 |
| *ADARB2.AS1* | 0.011 | 0.45 | 6.57E-01 | 6.83E-01 |
| *LAP3* | 0.012 | 0.44 | 6.63E-01 | 6.83E-01 |
| *BCDIN3D* | -0.009 | -0.36 | 7.19E-01 | 7.34E-01 |
| *SESTD1* | 0.004 | 0.14 | 8.90E-01 | 8.99E-01 |
| *FAM19A4* | 0.004 | 0.12 | 9.06E-01 | 9.06E-01 |

#### Supplementary Table 14d: Sensitivity analyses comprising Limma linear model statistics for normative spatial expression of *C9orf72* and the top 100 data-driven transcripts in the female donor.

| **Transcript** | **ß** | ***t*** | ***p*** | **Adjusted *p*** |
|---|---|---|---|---|
| *BAMBI* | 0.207 | 3.25 | 2.50E-03 | 1.81E-02 |
| *PPIF* | 0.203 | 3.22 | 2.70E-03 | 1.81E-02 |
| *TSPAN8* | 0.193 | 3.22 | 2.73E-03 | 1.81E-02 |

| | | | | |
|---|---|---|---|---|
| *RCOR2* | 0.170 | 3.18 | 3.05E-03 | 1.81E-02 |
| *DENND5A* | 0.207 | 3.15 | 3.26E-03 | 1.81E-02 |
| *PCBP4* | 0.187 | 3.12 | 3.53E-03 | 1.81E-02 |
| *TST* | 0.203 | 3.11 | 3.71E-03 | 1.81E-02 |
| *S100B* | 0.187 | 3.08 | 4.01E-03 | 1.81E-02 |
| *CLDND1* | 0.192 | 3.07 | 4.06E-03 | 1.81E-02 |
| *PLEKHB1* | 0.177 | 3.07 | 4.13E-03 | 1.81E-02 |
| *TNS2* | 0.197 | 3.06 | 4.20E-03 | 1.81E-02 |
| *MOBP* | 0.186 | 3.06 | 4.21E-03 | 1.81E-02 |
| *PAIP2B* | 0.201 | 3.05 | 4.34E-03 | 1.81E-02 |
| *AAMDC* | 0.179 | 3.03 | 4.57E-03 | 1.81E-02 |
| *IGFLR1* | 0.179 | 3.00 | 4.85E-03 | 1.81E-02 |
| *TCFL5* | 0.173 | 2.99 | 5.01E-03 | 1.81E-02 |
| *BCAR3* | 0.179 | 2.99 | 5.08E-03 | 1.81E-02 |
| *MAG* | 0.185 | 2.98 | 5.14E-03 | 1.81E-02 |
| *QDPR* | 0.182 | 2.98 | 5.16E-03 | 1.81E-02 |
| *CTDSP1* | 0.177 | 2.97 | 5.24E-03 | 1.81E-02 |
| *ADGRE5* | 0.186 | 2.97 | 5.31E-03 | 1.81E-02 |
| *RASSF2* | 0.187 | 2.96 | 5.39E-03 | 1.81E-02 |
| *ADAMTS4* | 0.179 | 2.96 | 5.42E-03 | 1.81E-02 |
| *SEPT4* | 0.168 | 2.96 | 5.51E-03 | 1.81E-02 |
| *SELENOP* | 0.181 | 2.95 | 5.53E-03 | 1.81E-02 |
| *USP54* | 0.183 | 2.95 | 5.59E-03 | 1.81E-02 |
| *TMEM140* | 0.175 | 2.94 | 5.75E-03 | 1.81E-02 |
| *SCRG1* | 0.177 | 2.93 | 5.82E-03 | 1.81E-02 |
| *PRKCQ* | 0.169 | 2.92 | 5.99E-03 | 1.81E-02 |
| *SLC5A11* | 0.175 | 2.92 | 6.06E-03 | 1.81E-02 |
| *FNTA* | 0.175 | 2.92 | 6.07E-03 | 1.81E-02 |
| *IFIT2* | 0.176 | 2.91 | 6.16E-03 | 1.81E-02 |
| *SLC31A2* | 0.173 | 2.91 | 6.26E-03 | 1.81E-02 |
| *MYLK* | 0.166 | 2.90 | 6.30E-03 | 1.81E-02 |
| *CTNNA3* | 0.164 | 2.88 | 6.68E-03 | 1.81E-02 |
| *TMEM98* | 0.176 | 2.87 | 6.87E-03 | 1.81E-02 |
| *SSFA2* | 0.168 | 2.86 | 7.01E-03 | 1.81E-02 |
| *FRMD5* | 0.153 | 2.86 | 7.02E-03 | 1.81E-02 |
| *LOC100506725* | 0.167 | 2.84 | 7.37E-03 | 1.81E-02 |
| *CAPN3* | 0.165 | 2.84 | 7.40E-03 | 1.81E-02 |
| *CRTAP* | 0.155 | 2.84 | 7.45E-03 | 1.81E-02 |
| *ARRDC2* | 0.162 | 2.83 | 7.63E-03 | 1.81E-02 |
| *NDE1* | 0.170 | 2.82 | 7.77E-03 | 1.81E-02 |
| *FEZ1* | 0.168 | 2.81 | 7.90E-03 | 1.81E-02 |
| *HHATL* | 0.161 | 2.79 | 8.37E-03 | 1.86E-02 |
| *ARHGEF37* | 0.169 | 2.79 | 8.48E-03 | 1.86E-02 |

| | | | | |
|---|---|---|---|---|
| *CARNS1* | 0.163 | 2.76 | 9.06E-03 | 1.92E-02 |
| *JPT2* | 0.153 | 2.75 | 9.23E-03 | 1.92E-02 |
| *KHNYN* | 0.167 | 2.74 | 9.46E-03 | 1.92E-02 |
| *HIPK2* | 0.155 | 2.74 | 9.52E-03 | 1.92E-02 |
| *CMTM5* | 0.147 | 2.72 | 9.93E-03 | 1.97E-02 |
| *SFR1* | 0.151 | 2.71 | 1.03E-02 | 1.98E-02 |
| *TMTC2* | 0.152 | 2.70 | 1.04E-02 | 1.98E-02 |
| *FAM149B1* | 0.141 | 2.69 | 1.07E-02 | 1.98E-02 |
| *SAP30* | 0.147 | 2.68 | 1.10E-02 | 1.98E-02 |
| *PCSK6* | 0.152 | 2.68 | 1.11E-02 | 1.98E-02 |
| *GALNT6* | 0.153 | 2.68 | 1.12E-02 | 1.98E-02 |
| *ABCA8* | 0.151 | 2.67 | 1.15E-02 | 2.00E-02 |
| *PARP4* | 0.156 | 2.63 | 1.24E-02 | 2.12E-02 |
| *DMBT1* | 0.142 | 2.62 | 1.29E-02 | 2.14E-02 |
| *CHST6* | 0.141 | 2.62 | 1.30E-02 | 2.14E-02 |
| *GPRC5B* | 0.141 | 2.58 | 1.43E-02 | 2.32E-02 |
| *CDH20* | 0.139 | 2.53 | 1.60E-02 | 2.56E-02 |
| *HSD17B3* | 0.143 | 2.46 | 1.90E-02 | 3.00E-02 |
| *RAB7B* | 0.125 | 2.42 | 2.07E-02 | 3.21E-02 |
| *LOC101929384* | 0.137 | 2.40 | 2.17E-02 | 3.33E-02 |
| *FAM19A4* | 0.161 | 2.38 | 2.25E-02 | 3.35E-02 |
| *ICK* | 0.160 | 2.38 | 2.26E-02 | 3.35E-02 |
| *ADAMTS14* | 0.120 | 2.33 | 2.59E-02 | 3.79E-02 |
| *KIAA1147* | 0.134 | 2.31 | 2.69E-02 | 3.88E-02 |
| *ADARB2.AS1* | 0.116 | 2.23 | 3.25E-02 | 4.62E-02 |
| *RDH11* | 0.112 | 2.20 | 3.46E-02 | 4.86E-02 |
| *C9orf3* | -0.118 | -2.16 | 3.80E-02 | 5.25E-02 |
| *ANKRD65* | 0.110 | 2.14 | 3.95E-02 | 5.39E-02 |
| *ARHGEF1* | 0.097 | 1.97 | 5.61E-02 | 7.50E-02 |
| *KATNA1* | 0.087 | 1.97 | 5.64E-02 | 7.50E-02 |
| *C9orf72* | 0.087 | 1.86 | 7.14E-02 | 9.36E-02 |
| *ZNF664* | 0.093 | 1.83 | 7.55E-02 | 9.77E-02 |
| *SESTD1* | 0.120 | 1.82 | 7.79E-02 | 9.96E-02 |
| *ARHGAP21* | -0.097 | -1.79 | 8.26E-02 | 1.04E-01 |
| *ASPRV1* | 0.091 | 1.77 | 8.59E-02 | 1.07E-01 |
| *TMTC4* | 0.081 | 1.72 | 9.48E-02 | 1.17E-01 |
| *RGS1* | 0.097 | 1.68 | 1.02E-01 | 1.24E-01 |
| *BCDIN3D* | 0.083 | 1.66 | 1.06E-01 | 1.25E-01 |
| *LRP6* | -0.094 | -1.66 | 1.06E-01 | 1.25E-01 |
| *USH1C* | -0.080 | -1.46 | 1.54E-01 | 1.80E-01 |
| *ZBED3* | 0.063 | 1.34 | 1.89E-01 | 2.19E-01 |
| *PHF10* | 0.065 | 1.30 | 2.01E-01 | 2.31E-01 |
| *FLNC* | 0.051 | 1.29 | 2.06E-01 | 2.33E-01 |

| | | | | |
|---|---|---|---|---|
| *LLGL2* | 0.059 | 1.25 | 2.18E-01 | 2.45E-01 |
| *B3GNT9* | 0.062 | 1.23 | 2.25E-01 | 2.50E-01 |
| *ENPP6* | -0.051 | -0.96 | 3.42E-01 | 3.75E-01 |
| *LDLRAD4* | 0.047 | 0.79 | 4.34E-01 | 4.72E-01 |
| *SMARCC1* | 0.036 | 0.72 | 4.74E-01 | 5.03E-01 |
| *FAT1* | 0.031 | 0.72 | 4.77E-01 | 5.03E-01 |
| *VKORC1* | 0.032 | 0.72 | 4.78E-01 | 5.03E-01 |
| *TSPAN15* | 0.027 | 0.61 | 5.45E-01 | 5.68E-01 |
| *NUDT1* | -0.032 | -0.54 | 5.92E-01 | 6.11E-01 |
| *LAP3* | 0.021 | 0.51 | 6.16E-01 | 6.28E-01 |
| *KLC1* | -0.019 | -0.42 | 6.74E-01 | 6.81E-01 |
| *LOC100507487* | 0.014 | 0.36 | 7.25E-01 | 7.25E-01 |

#### Supplementary Table 14e: Sensitivity analyses comprising Limma linear model statistics for normative spatial expression of *C9orf72* and the top 100 data-driven transcripts in the first mid-to-late fifties male donor.

| **Transcript** | ***ß*** | ***t*** | ***p*** | **Adjusted *p*** |
|---|---|---|---|---|
| *LLGL2* | 0.100 | 2.05 | 4.50E-02 | 8.90E-01 |
| *ICK* | 0.108 | 1.77 | 8.12E-02 | 8.90E-01 |
| *PHF10* | 0.082 | 1.69 | 9.66E-02 | 8.90E-01 |
| *ENPP6* | 0.073 | 1.47 | 1.46E-01 | 8.90E-01 |
| *SLC5A11* | 0.077 | 1.45 | 1.52E-01 | 8.90E-01 |
| *RGS1* | 0.075 | 1.44 | 1.55E-01 | 8.90E-01 |
| *USP54* | 0.068 | 1.37 | 1.75E-01 | 8.90E-01 |
| *ANKRD65* | 0.065 | 1.35 | 1.81E-01 | 8.90E-01 |
| *PCSK6* | 0.068 | 1.34 | 1.85E-01 | 8.90E-01 |
| *BCAR3* | -0.071 | -1.33 | 1.90E-01 | 8.90E-01 |
| *TMTC4* | 0.075 | 1.32 | 1.93E-01 | 8.90E-01 |
| *TMEM140* | 0.060 | 1.31 | 1.96E-01 | 8.90E-01 |
| *SLC31A2* | 0.067 | 1.29 | 2.03E-01 | 8.90E-01 |
| *ADAMTS4* | 0.066 | 1.26 | 2.11E-01 | 8.90E-01 |
| *ADAMTS14* | 0.062 | 1.26 | 2.13E-01 | 8.90E-01 |
| *LOC100506725* | 0.067 | 1.25 | 2.18E-01 | 8.90E-01 |
| *FRMD5* | 0.065 | 1.24 | 2.18E-01 | 8.90E-01 |
| *GALNT6* | 0.064 | 1.23 | 2.24E-01 | 8.90E-01 |
| *TSPAN15* | 0.063 | 1.21 | 2.29E-01 | 8.90E-01 |
| *CARNS1* | 0.063 | 1.21 | 2.30E-01 | 8.90E-01 |
| *DMBT1* | 0.066 | 1.20 | 2.36E-01 | 8.90E-01 |
| *SFR1* | 0.065 | 1.19 | 2.38E-01 | 8.90E-01 |
| *RDH11* | 0.057 | 1.17 | 2.47E-01 | 8.90E-01 |
| *TMEM98* | 0.062 | 1.16 | 2.52E-01 | 8.90E-01 |
| *MAG* | 0.059 | 1.13 | 2.64E-01 | 8.90E-01 |

| | | | | |
|---|---|---|---|---|
| *LAP3* | 0.061 | 1.10 | 2.75E-01 | 8.90E-01 |
| *BCDIN3D* | 0.058 | 1.07 | 2.88E-01 | 8.90E-01 |
| *ABCA8* | 0.054 | 1.02 | 3.10E-01 | 8.90E-01 |
| *PARP4* | 0.052 | 1.01 | 3.17E-01 | 8.90E-01 |
| *JPT2* | 0.055 | 1.00 | 3.20E-01 | 8.90E-01 |
| *FAM19A4* | 0.061 | 0.99 | 3.25E-01 | 8.90E-01 |
| *SESTD1* | 0.053 | 0.96 | 3.39E-01 | 8.90E-01 |
| *C9orf3* | 0.049 | 0.96 | 3.42E-01 | 8.90E-01 |
| *SAP30* | 0.047 | 0.95 | 3.45E-01 | 8.90E-01 |
| *HSD17B3* | 0.047 | 0.94 | 3.50E-01 | 8.90E-01 |
| *NUDT1* | 0.056 | 0.94 | 3.53E-01 | 8.90E-01 |
| *CAPN3* | 0.050 | 0.94 | 3.53E-01 | 8.90E-01 |
| *FEZ1* | 0.047 | 0.92 | 3.59E-01 | 8.90E-01 |
| *ADARB2.AS1* | 0.045 | 0.90 | 3.69E-01 | 8.90E-01 |
| *NDE1* | 0.046 | 0.88 | 3.85E-01 | 8.90E-01 |
| *PCBP4* | 0.045 | 0.85 | 4.00E-01 | 8.90E-01 |
| *KIAA1147* | 0.041 | 0.84 | 4.05E-01 | 8.90E-01 |
| *SSFA2* | -0.044 | -0.83 | 4.08E-01 | 8.90E-01 |
| *SELENOP* | 0.044 | 0.80 | 4.27E-01 | 8.90E-01 |
| *FAM149B1* | 0.044 | 0.80 | 4.28E-01 | 8.90E-01 |
| *TMTC2* | 0.044 | 0.79 | 4.31E-01 | 8.90E-01 |
| *CLDND1* | 0.042 | 0.79 | 4.35E-01 | 8.90E-01 |
| *CRTAP* | -0.044 | -0.78 | 4.37E-01 | 8.90E-01 |
| *FAT1* | -0.041 | -0.77 | 4.44E-01 | 8.90E-01 |
| *RAB7B* | 0.040 | 0.76 | 4.49E-01 | 8.90E-01 |
| *LOC101929384* | 0.039 | 0.74 | 4.64E-01 | 8.90E-01 |
| *ARRDC2* | 0.036 | 0.72 | 4.72E-01 | 8.90E-01 |
| *HHATL* | 0.039 | 0.72 | 4.74E-01 | 8.90E-01 |
| *ZBED3* | 0.039 | 0.72 | 4.76E-01 | 8.90E-01 |
| *KATNA1* | 0.036 | 0.69 | 4.91E-01 | 8.99E-01 |
| *SCRG1* | -0.035 | -0.68 | 5.01E-01 | 8.99E-01 |
| *ARHGAP21* | 0.034 | 0.66 | 5.11E-01 | 8.99E-01 |
| *QDPR* | 0.036 | 0.65 | 5.16E-01 | 8.99E-01 |
| *LDLRAD4* | 0.033 | 0.64 | 5.26E-01 | 9.00E-01 |
| *IGFLR1* | -0.034 | -0.60 | 5.50E-01 | 9.26E-01 |
| *USH1C* | 0.030 | 0.57 | 5.71E-01 | 9.45E-01 |
| *FNTA* | 0.031 | 0.56 | 5.80E-01 | 9.45E-01 |
| *SEPT4* | 0.029 | 0.54 | 5.89E-01 | 9.45E-01 |
| *ZNF664* | -0.027 | -0.51 | 6.09E-01 | 9.50E-01 |
| *LOC100507487* | 0.027 | 0.51 | 6.12E-01 | 9.50E-01 |
| *KLC1* | 0.020 | 0.41 | 6.82E-01 | 9.80E-01 |
| *C9orf72* | 0.021 | 0.39 | 7.01E-01 | 9.80E-01 |
| *CDH20* | -0.020 | -0.38 | 7.05E-01 | 9.80E-01 |

| | | | | |
|---|---|---|---|---|
| *CTNNA3* | 0.020 | 0.38 | 7.05E-01 | 9.80E-01 |
| *AAMDC* | 0.022 | 0.38 | 7.06E-01 | 9.80E-01 |
| *PAIP2B* | 0.019 | 0.34 | 7.37E-01 | 9.80E-01 |
| *GPRC5B* | 0.018 | 0.33 | 7.40E-01 | 9.80E-01 |
| *CMTM5* | 0.017 | 0.32 | 7.51E-01 | 9.80E-01 |
| *ARHGEF1* | 0.017 | 0.32 | 7.51E-01 | 9.80E-01 |
| *TNS2* | -0.016 | -0.29 | 7.71E-01 | 9.80E-01 |
| *BAMBI* | 0.014 | 0.28 | 7.81E-01 | 9.80E-01 |
| *RASSF2* | 0.015 | 0.28 | 7.83E-01 | 9.80E-01 |
| *MYLK* | 0.013 | 0.26 | 7.97E-01 | 9.80E-01 |
| *ADGRE5* | 0.013 | 0.25 | 8.02E-01 | 9.80E-01 |
| *FLNC* | -0.011 | -0.25 | 8.03E-01 | 9.80E-01 |
| *B3GNT9* | -0.012 | -0.24 | 8.11E-01 | 9.80E-01 |
| *LRP6* | -0.011 | -0.23 | 8.20E-01 | 9.80E-01 |
| *RCOR2* | 0.010 | 0.22 | 8.26E-01 | 9.80E-01 |
| *CHST6* | -0.011 | -0.20 | 8.46E-01 | 9.80E-01 |
| *SMARCC1* | -0.011 | -0.19 | 8.46E-01 | 9.80E-01 |
| *TSPAN8* | 0.011 | 0.19 | 8.49E-01 | 9.80E-01 |
| *HIPK2* | 0.009 | 0.17 | 8.66E-01 | 9.80E-01 |
| *ASPRV1* | -0.009 | -0.17 | 8.67E-01 | 9.80E-01 |
| *KHNYN* | -0.008 | -0.15 | 8.78E-01 | 9.80E-01 |
| *DENND5A* | 0.009 | 0.15 | 8.82E-01 | 9.80E-01 |
| *ARHGEF37* | 0.006 | 0.11 | 9.13E-01 | 9.80E-01 |
| *CTDSP1* | -0.006 | -0.11 | 9.16E-01 | 9.80E-01 |
| *TST* | -0.006 | -0.11 | 9.16E-01 | 9.80E-01 |
| *PLEKHB1* | -0.005 | -0.09 | 9.27E-01 | 9.80E-01 |
| *VKORC1* | 0.005 | 0.09 | 9.29E-01 | 9.80E-01 |
| *PPIF* | -0.005 | -0.09 | 9.31E-01 | 9.80E-01 |
| *MOBP* | 0.004 | 0.07 | 9.47E-01 | 9.86E-01 |
| *IFIT2* | 0.002 | 0.04 | 9.67E-01 | 9.86E-01 |
| *S100B* | 0.002 | 0.04 | 9.72E-01 | 9.86E-01 |
| *TCFL5* | 0.002 | 0.03 | 9.76E-01 | 9.86E-01 |
| *PRKCQ* | 0.000 | 0.00 | 9.97E-01 | 9.97E-01 |

#### Supplementary Table 14f: Sensitivity analyses comprising Limma linear model statistics for normative spatial expression of *C9orf72* and the top 100 data-driven transcripts in the second mid-to-late fifties male donor.

| Transcript | *ß* | *t* | *p* | Adjusted *p* |
|---|---|---|---|---|
| *BAMBI* | 0.069 | 2.49 | 2.02E-02 | 5.01E-01 |
| *C9orf72* | -0.071 | -2.46 | 2.19E-02 | 5.01E-01 |
| *SMARCC1* | 0.089 | 2.30 | 3.08E-02 | 5.01E-01 |
| *USH1C* | 0.100 | 2.23 | 3.58E-02 | 5.01E-01 |

| | | | | |
|---|---|---|---|---|
| *SSFA2* | 0.042 | 2.19 | 3.85E-02 | 5.01E-01 |
| *RAB7B* | 0.058 | 2.16 | 4.11E-02 | 5.01E-01 |
| *IGFLR1* | 0.046 | 2.11 | 4.54E-02 | 5.01E-01 |
| *FAM19A4* | -0.030 | -2.10 | 4.67E-02 | 5.01E-01 |
| *ARHGEF1* | 0.073 | 2.09 | 4.80E-02 | 5.01E-01 |
| *HIPK2* | 0.050 | 2.00 | 5.71E-02 | 5.37E-01 |
| *AAMDC* | 0.048 | 1.91 | 6.79E-02 | 5.50E-01 |
| *RCOR2* | -0.078 | -1.90 | 7.05E-02 | 5.50E-01 |
| *KHNYN* | -0.041 | -1.83 | 8.05E-02 | 5.50E-01 |
| *CRTAP* | 0.070 | 1.82 | 8.19E-02 | 5.50E-01 |
| *RGS1* | -0.031 | -1.57 | 1.29E-01 | 7.87E-01 |
| *LRP6* | 0.040 | 1.54 | 1.37E-01 | 7.87E-01 |
| *CHST6* | 0.021 | 1.52 | 1.43E-01 | 7.87E-01 |
| *LLGL2* | 0.066 | 1.44 | 1.63E-01 | 7.87E-01 |
| *SCRG1* | 0.041 | 1.44 | 1.63E-01 | 7.87E-01 |
| *IFIT2* | 0.043 | 1.42 | 1.67E-01 | 7.87E-01 |
| *ICK* | -0.032 | -1.38 | 1.82E-01 | 8.15E-01 |
| *MOBP* | 0.039 | 1.34 | 1.94E-01 | 8.23E-01 |
| *ARRDC2* | 0.049 | 1.31 | 2.01E-01 | 8.23E-01 |
| *LDLRAD4* | -0.044 | -1.28 | 2.15E-01 | 8.41E-01 |
| *CTDSP1* | 0.018 | 1.14 | 2.64E-01 | 9.62E-01 |
| *C9orf3* | 0.035 | 1.11 | 2.77E-01 | 9.62E-01 |
| *KIAA1147* | 0.029 | 1.04 | 3.08E-01 | 9.62E-01 |
| *PAIP2B* | 0.031 | 1.03 | 3.12E-01 | 9.62E-01 |
| *JPT2* | 0.034 | 1.02 | 3.16E-01 | 9.62E-01 |
| *FAT1* | 0.016 | 0.99 | 3.32E-01 | 9.62E-01 |
| *ARHGEF37* | 0.032 | 0.96 | 3.48E-01 | 9.62E-01 |
| *ANKRD65* | -0.045 | -0.96 | 3.49E-01 | 9.62E-01 |
| *SFR1* | -0.026 | -0.94 | 3.56E-01 | 9.62E-01 |
| *ADARB2.AS1* | -0.038 | -0.93 | 3.61E-01 | 9.62E-01 |
| *ADGRE5* | 0.026 | 0.92 | 3.66E-01 | 9.62E-01 |
| *PARP4* | -0.029 | -0.89 | 3.80E-01 | 9.62E-01 |
| *TNS2* | 0.041 | 0.88 | 3.87E-01 | 9.62E-01 |
| *GPRC5B* | -0.030 | -0.85 | 4.02E-01 | 9.62E-01 |
| *CTNNA3* | 0.033 | 0.85 | 4.05E-01 | 9.62E-01 |
| *PLEKHB1* | 0.020 | 0.82 | 4.22E-01 | 9.62E-01 |
| *SESTD1* | -0.035 | -0.80 | 4.30E-01 | 9.62E-01 |
| *MYLK* | 0.029 | 0.79 | 4.36E-01 | 9.62E-01 |
| *ZBED3* | 0.023 | 0.78 | 4.44E-01 | 9.62E-01 |
| *TMTC2* | -0.031 | -0.77 | 4.50E-01 | 9.62E-01 |
| *ZNF664* | 0.024 | 0.73 | 4.71E-01 | 9.76E-01 |
| *LAP3* | -0.029 | -0.71 | 4.83E-01 | 9.76E-01 |
| *FRMD5* | -0.019 | -0.70 | 4.88E-01 | 9.76E-01 |

| | | | | |
|---|---|---|---|---|
| *LOC100506725* | -0.037 | -0.67 | 5.08E-01 | 9.95E-01 |
| *HHATL* | -0.024 | -0.65 | 5.23E-01 | 9.98E-01 |
| *B3GNT9* | -0.017 | -0.61 | 5.45E-01 | 9.98E-01 |
| *TMEM140* | -0.033 | -0.53 | 6.03E-01 | 9.98E-01 |
| *PPIF* | -0.007 | -0.50 | 6.23E-01 | 9.98E-01 |
| *SAP30* | -0.017 | -0.49 | 6.31E-01 | 9.98E-01 |
| *FNTA* | 0.013 | 0.47 | 6.40E-01 | 9.98E-01 |
| *TCFL5* | 0.012 | 0.46 | 6.47E-01 | 9.98E-01 |
| *RDH11* | 0.015 | 0.43 | 6.68E-01 | 9.98E-01 |
| *TMEM98* | -0.019 | -0.41 | 6.82E-01 | 9.98E-01 |
| *PRKCQ* | -0.007 | -0.39 | 6.97E-01 | 9.98E-01 |
| *DENND5A* | -0.005 | -0.39 | 7.00E-01 | 9.98E-01 |
| *TST* | -0.004 | -0.39 | 7.03E-01 | 9.98E-01 |
| *SELENOP* | -0.017 | -0.37 | 7.16E-01 | 9.98E-01 |
| *TSPAN15* | -0.016 | -0.35 | 7.28E-01 | 9.98E-01 |
| *ASPRV1* | 0.011 | 0.35 | 7.32E-01 | 9.98E-01 |
| *KLC1* | 0.006 | 0.34 | 7.38E-01 | 9.98E-01 |
| *ARHGAP21* | 0.014 | 0.34 | 7.40E-01 | 9.98E-01 |
| *S100B* | 0.007 | 0.33 | 7.48E-01 | 9.98E-01 |
| *RASSF2* | 0.008 | 0.32 | 7.49E-01 | 9.98E-01 |
| *CARNS1* | -0.016 | -0.31 | 7.59E-01 | 9.98E-01 |
| *NDE1* | 0.009 | 0.31 | 7.62E-01 | 9.98E-01 |
| *CMTM5* | -0.007 | -0.28 | 7.79E-01 | 9.98E-01 |
| *SLC31A2* | -0.011 | -0.25 | 8.04E-01 | 9.98E-01 |
| *SLC5A11* | 0.008 | 0.23 | 8.18E-01 | 9.98E-01 |
| *GALNT6* | 0.009 | 0.18 | 8.59E-01 | 9.98E-01 |
| *VKORC1* | 0.007 | 0.18 | 8.62E-01 | 9.98E-01 |
| *QDPR* | -0.009 | -0.17 | 8.69E-01 | 9.98E-01 |
| *BCAR3* | -0.005 | -0.15 | 8.80E-01 | 9.98E-01 |
| *LOC101929384* | -0.005 | -0.15 | 8.83E-01 | 9.98E-01 |
| *PHF10* | 0.004 | 0.11 | 9.13E-01 | 9.98E-01 |
| *PCBP4* | 0.002 | 0.11 | 9.16E-01 | 9.98E-01 |
| *PCSK6* | -0.004 | -0.11 | 9.17E-01 | 9.98E-01 |
| *FAM149B1* | 0.002 | 0.09 | 9.33E-01 | 9.98E-01 |
| *MAG* | -0.004 | -0.08 | 9.35E-01 | 9.98E-01 |
| *SEPT4* | 0.003 | 0.08 | 9.40E-01 | 9.98E-01 |
| *TMTC4* | 0.003 | 0.07 | 9.47E-01 | 9.98E-01 |
| *CLDND1* | -0.002 | -0.05 | 9.60E-01 | 9.98E-01 |
| *BCDIN3D* | 0.002 | 0.05 | 9.62E-01 | 9.98E-01 |
| *CDH20* | 0.002 | 0.04 | 9.69E-01 | 9.98E-01 |
| *NUDT1* | -0.001 | -0.04 | 9.71E-01 | 9.98E-01 |
| *KATNA1* | -0.001 | -0.03 | 9.76E-01 | 9.98E-01 |
| *ADAMTS4* | 0.001 | 0.01 | 9.88E-01 | 9.98E-01 |

| | | | | |
|---|---|---|---|---|
| *DMBT1* | 0.000 | 0.01 | 9.93E-01 | 9.98E-01 |
| *CAPN3* | 0.000 | 0.01 | 9.94E-01 | 9.98E-01 |
| *USP54* | 0.000 | -0.01 | 9.96E-01 | 9.98E-01 |
| *FEZ1* | 0.000 | 0.00 | 9.98E-01 | 9.98E-01 |

#### Supplementary Table 14g: Sensitivity analyses comprising Limma linear model statistics for normative spatial expression of *C9orf72* and the top 100 data-driven transcripts in the two younger AHBA donors in aggregate.

| **Transcript** | ***ß*** | ***t*** | ***p*** | **Adjusted *p*** |
|---|---|---|---|---|
| *ARRDC2* | 0.058 | 3.37 | 1.40E-03 | 1.22E-01 |
| *ENPP6* | 0.044 | 2.78 | 7.54E-03 | 1.22E-01 |
| *RAB7B* | 0.046 | 2.77 | 7.75E-03 | 1.22E-01 |
| *PRKCQ* | 0.049 | 2.76 | 7.94E-03 | 1.22E-01 |
| *TSPAN8* | 0.045 | 2.61 | 1.18E-02 | 1.22E-01 |
| *MYLK* | 0.050 | 2.59 | 1.23E-02 | 1.22E-01 |
| *ZBED3* | 0.031 | 2.57 | 1.31E-02 | 1.22E-01 |
| *ARHGEF37* | 0.031 | 2.55 | 1.37E-02 | 1.22E-01 |
| *PAIP2B* | 0.028 | 2.55 | 1.38E-02 | 1.22E-01 |
| *CTNNA3* | 0.035 | 2.52 | 1.49E-02 | 1.22E-01 |
| *ANKRD65* | 0.041 | 2.51 | 1.51E-02 | 1.22E-01 |
| *NDE1* | 0.027 | 2.48 | 1.62E-02 | 1.22E-01 |
| *IGFLR1* | 0.042 | 2.43 | 1.86E-02 | 1.22E-01 |
| *HIPK2* | 0.024 | 2.43 | 1.88E-02 | 1.22E-01 |
| *LDLRAD4* | 0.036 | 2.41 | 1.94E-02 | 1.22E-01 |
| *MOBP* | 0.026 | 2.38 | 2.09E-02 | 1.23E-01 |
| *SAP30* | 0.039 | 2.34 | 2.31E-02 | 1.28E-01 |
| *ADAMTS4* | 0.028 | 2.29 | 2.57E-02 | 1.28E-01 |
| *QDPR* | 0.025 | 2.27 | 2.71E-02 | 1.28E-01 |
| *JPT2* | 0.031 | 2.26 | 2.83E-02 | 1.28E-01 |
| *ASPRV1* | 0.028 | 2.25 | 2.87E-02 | 1.28E-01 |
| *CLDND1* | 0.029 | 2.20 | 3.22E-02 | 1.38E-01 |
| *USH1C* | 0.041 | 2.16 | 3.56E-02 | 1.46E-01 |
| *TMEM140* | 0.032 | 2.11 | 3.98E-02 | 1.51E-01 |
| *SSFA2* | 0.031 | 2.09 | 4.15E-02 | 1.51E-01 |
| *CMTM5* | 0.036 | 2.08 | 4.26E-02 | 1.51E-01 |
| *MAG* | 0.023 | 2.05 | 4.57E-02 | 1.51E-01 |
| *IFIT2* | 0.033 | 2.03 | 4.75E-02 | 1.51E-01 |
| *KATNA1* | 0.030 | 2.02 | 4.86E-02 | 1.51E-01 |
| *SMARCC1* | 0.028 | 2.01 | 4.90E-02 | 1.51E-01 |
| *C9orf3* | 0.034 | 2.01 | 5.00E-02 | 1.51E-01 |
| *DMBT1* | 0.032 | 1.97 | 5.45E-02 | 1.58E-01 |
| *CARNS1* | 0.027 | 1.96 | 5.54E-02 | 1.58E-01 |

| | | | | |
|---|---|---|---|---|
| *SLC31A2* | 0.031 | 1.93 | 5.89E-02 | 1.61E-01 |
| *SELENOP* | 0.026 | 1.92 | 5.99E-02 | 1.61E-01 |
| *TMEM98* | 0.026 | 1.88 | 6.60E-02 | 1.68E-01 |
| *CHST6* | 0.022 | 1.88 | 6.61E-02 | 1.68E-01 |
| *USP54* | 0.031 | 1.81 | 7.67E-02 | 1.85E-01 |
| *VKORC1* | 0.035 | 1.80 | 7.71E-02 | 1.85E-01 |
| *CAPN3* | 0.020 | 1.79 | 7.89E-02 | 1.85E-01 |
| *ABCA8* | 0.031 | 1.77 | 8.22E-02 | 1.89E-01 |
| *ADGRE5* | 0.025 | 1.73 | 8.98E-02 | 2.01E-01 |
| *BAMBI* | 0.024 | 1.71 | 9.39E-02 | 2.05E-01 |
| *NUDT1* | 0.034 | 1.66 | 1.04E-01 | 2.19E-01 |
| *KLC1* | 0.034 | 1.64 | 1.07E-01 | 2.19E-01 |
| *TNS2* | 0.023 | 1.64 | 1.07E-01 | 2.19E-01 |
| *S100B* | 0.022 | 1.61 | 1.14E-01 | 2.28E-01 |
| *TMTC4* | 0.030 | 1.58 | 1.19E-01 | 2.30E-01 |
| *LOC100506725* | 0.019 | 1.58 | 1.20E-01 | 2.30E-01 |
| *PCBP4* | 0.028 | 1.53 | 1.31E-01 | 2.44E-01 |
| *SCRG1* | 0.038 | 1.53 | 1.32E-01 | 2.44E-01 |
| *ARHGAP21* | 0.026 | 1.52 | 1.35E-01 | 2.44E-01 |
| *FAM19A4* | -0.019 | -1.48 | 1.46E-01 | 2.59E-01 |
| *PHF10* | 0.028 | 1.43 | 1.58E-01 | 2.75E-01 |
| *RDH11* | 0.020 | 1.36 | 1.80E-01 | 3.07E-01 |
| *CRTAP* | 0.021 | 1.34 | 1.87E-01 | 3.14E-01 |
| *TSPAN15* | 0.022 | 1.24 | 2.19E-01 | 3.59E-01 |
| *B3GNT9* | 0.025 | 1.23 | 2.24E-01 | 3.59E-01 |
| *LLGL2* | 0.022 | 1.23 | 2.25E-01 | 3.59E-01 |
| *PPIF* | 0.021 | 1.20 | 2.37E-01 | 3.72E-01 |
| *SESTD1* | -0.014 | -1.18 | 2.42E-01 | 3.74E-01 |
| *FAM149B1* | -0.021 | -1.14 | 2.60E-01 | 3.86E-01 |
| *SEPT4* | 0.019 | 1.14 | 2.61E-01 | 3.86E-01 |
| *FNTA* | 0.022 | 1.13 | 2.65E-01 | 3.86E-01 |
| *PARP4* | 0.018 | 1.12 | 2.67E-01 | 3.86E-01 |
| *LAP3* | 0.025 | 1.11 | 2.74E-01 | 3.90E-01 |
| *ARHGEF1* | 0.024 | 1.08 | 2.85E-01 | 3.96E-01 |
| *FEZ1* | 0.020 | 1.08 | 2.87E-01 | 3.96E-01 |
| *FAT1* | 0.016 | 1.05 | 2.99E-01 | 4.08E-01 |
| *RASSF2* | 0.016 | 1.02 | 3.10E-01 | 4.17E-01 |
| *PLEKHB1* | 0.013 | 0.99 | 3.25E-01 | 4.26E-01 |
| *CDH20* | 0.016 | 0.99 | 3.26E-01 | 4.26E-01 |
| *RCOR2* | -0.014 | -0.94 | 3.52E-01 | 4.54E-01 |
| *GPRC5B* | 0.016 | 0.92 | 3.62E-01 | 4.60E-01 |
| *AAMDC* | 0.016 | 0.90 | 3.71E-01 | 4.65E-01 |
| *HHATL* | 0.016 | 0.88 | 3.84E-01 | 4.74E-01 |

| | | | | |
|---|---|---|---|---|
| *PCSK6* | 0.013 | 0.86 | 3.91E-01 | 4.77E-01 |
| *KHNYN* | 0.016 | 0.84 | 4.02E-01 | 4.85E-01 |
| *TST* | 0.017 | 0.80 | 4.29E-01 | 5.10E-01 |
| *C9orf72* | 0.011 | 0.72 | 4.74E-01 | 5.57E-01 |
| *LRP6* | 0.014 | 0.71 | 4.80E-01 | 5.57E-01 |
| *ZNF664* | 0.015 | 0.66 | 5.10E-01 | 5.84E-01 |
| *TCFL5* | 0.010 | 0.64 | 5.24E-01 | 5.94E-01 |
| *CTDSP1* | 0.012 | 0.60 | 5.54E-01 | 6.19E-01 |
| *BCAR3* | 0.009 | 0.54 | 5.94E-01 | 6.57E-01 |
| *TMTC2* | 0.008 | 0.48 | 6.32E-01 | 6.84E-01 |
| *SLC5A11* | 0.005 | 0.47 | 6.38E-01 | 6.84E-01 |
| *DENND5A* | 0.006 | 0.47 | 6.41E-01 | 6.84E-01 |
| *KIAA1147* | 0.007 | 0.43 | 6.70E-01 | 7.07E-01 |
| *ADARB2.AS1* | -0.007 | -0.37 | 7.14E-01 | 7.46E-01 |
| *ICK* | -0.006 | -0.34 | 7.39E-01 | 7.57E-01 |
| *SFR1* | 0.006 | 0.33 | 7.41E-01 | 7.57E-01 |
| *BCDIN3D* | -0.004 | -0.25 | 8.03E-01 | 8.12E-01 |
| *FRMD5* | 0.00022 | 0.01 | 9.90E-01 | 9.90E-01 |

#### Supplementary Table 14h: Sensitivity analyses comprising Limma linear model statistics for normative spatial expression of *C9orf72* and the top 100 data-driven transcripts in the four older AHBA donors in aggregate.

| **Transcript** | ***ß*** | ***t*** | ***p*** | **Adjusted *p*** |
|---|---|---|---|---|
| *C9orf3* | 0.105 | 3.81 | 1.38E-04 | 1.02E-02 |
| *DMBT1* | 0.101 | 3.66 | 2.52E-04 | 1.02E-02 |
| *ZBED3* | 0.096 | 3.49 | 4.89E-04 | 1.02E-02 |
| *RAB7B* | 0.095 | 3.46 | 5.36E-04 | 1.02E-02 |
| *KIAA1147* | 0.095 | 3.44 | 5.86E-04 | 1.02E-02 |
| *TSPAN15* | 0.092 | 3.35 | 8.24E-04 | 1.02E-02 |
| *PCSK6* | 0.090 | 3.26 | 1.13E-03 | 1.02E-02 |
| *JPT2* | 0.089 | 3.26 | 1.14E-03 | 1.02E-02 |
| *LOC101929384* | 0.088 | 3.21 | 1.34E-03 | 1.02E-02 |
| *ASPRV1* | 0.087 | 3.17 | 1.53E-03 | 1.02E-02 |
| *ENPP6* | 0.087 | 3.17 | 1.53E-03 | 1.02E-02 |
| *FEZ1* | 0.087 | 3.16 | 1.60E-03 | 1.02E-02 |
| *SLC5A11* | 0.086 | 3.14 | 1.67E-03 | 1.02E-02 |
| *ARHGAP21* | 0.086 | 3.12 | 1.85E-03 | 1.02E-02 |
| *GALNT6* | 0.085 | 3.10 | 1.95E-03 | 1.02E-02 |
| *PCBP4* | 0.085 | 3.10 | 1.97E-03 | 1.02E-02 |
| *USP54* | 0.085 | 3.09 | 2.04E-03 | 1.02E-02 |
| *TMTC2* | 0.084 | 3.07 | 2.13E-03 | 1.02E-02 |

| | | | | |
|---|---|---|---|---|
| *KHNYN* | 0.084 | 3.06 | 2.25E-03 | 1.02E-02 |
| *CTDSP1* | 0.083 | 3.02 | 2.51E-03 | 1.02E-02 |
| *FAM149B1* | 0.083 | 3.02 | 2.58E-03 | 1.02E-02 |
| *CLDND1* | 0.083 | 3.01 | 2.60E-03 | 1.02E-02 |
| *USH1C* | 0.083 | 3.01 | 2.60E-03 | 1.02E-02 |
| *TSPAN8* | 0.083 | 3.00 | 2.70E-03 | 1.02E-02 |
| *MAG* | 0.082 | 3.00 | 2.70E-03 | 1.02E-02 |
| *CARNS1* | 0.082 | 3.00 | 2.75E-03 | 1.02E-02 |
| *NDE1* | 0.082 | 2.98 | 2.86E-03 | 1.02E-02 |
| *HHATL* | 0.082 | 2.98 | 2.90E-03 | 1.02E-02 |
| *ABCA8* | 0.081 | 2.96 | 3.06E-03 | 1.02E-02 |
| *SFR1* | 0.081 | 2.95 | 3.21E-03 | 1.02E-02 |
| *BAMBI* | 0.081 | 2.95 | 3.24E-03 | 1.02E-02 |
| *HSD17B3* | 0.081 | 2.94 | 3.30E-03 | 1.02E-02 |
| *PARP4* | 0.081 | 2.94 | 3.34E-03 | 1.02E-02 |
| *ADGRE5* | 0.080 | 2.92 | 3.49E-03 | 1.04E-02 |
| *TMEM98* | 0.080 | 2.91 | 3.65E-03 | 1.05E-02 |
| *SLC31A2* | 0.079 | 2.89 | 3.90E-03 | 1.05E-02 |
| *LRP6* | 0.079 | 2.88 | 4.03E-03 | 1.05E-02 |
| *ADAMTS4* | 0.079 | 2.88 | 4.04E-03 | 1.05E-02 |
| *ADARB2.AS1* | 0.079 | 2.88 | 4.05E-03 | 1.05E-02 |
| *FLNC* | 0.078 | 2.84 | 4.51E-03 | 1.14E-02 |
| *CRTAP* | 0.078 | 2.83 | 4.70E-03 | 1.16E-02 |
| *SMARCC1* | 0.076 | 2.78 | 5.42E-03 | 1.30E-02 |
| *IFIT2* | 0.076 | 2.76 | 5.80E-03 | 1.36E-02 |
| *ARHGEF37* | 0.075 | 2.74 | 6.15E-03 | 1.41E-02 |
| *ZNF664* | 0.075 | 2.72 | 6.52E-03 | 1.46E-02 |
| *NUDT1* | 0.075 | 2.71 | 6.66E-03 | 1.46E-02 |
| *SAP30* | 0.074 | 2.70 | 7.05E-03 | 1.52E-02 |
| *ANKRD65* | 0.073 | 2.67 | 7.67E-03 | 1.61E-02 |
| *GPRC5B* | 0.073 | 2.66 | 7.82E-03 | 1.61E-02 |
| *SELENOP* | 0.073 | 2.65 | 8.06E-03 | 1.63E-02 |
| *RASSF2* | 0.073 | 2.64 | 8.22E-03 | 1.63E-02 |
| *CAPN3* | 0.072 | 2.62 | 8.72E-03 | 1.66E-02 |
| *LDLRAD4* | 0.072 | 2.62 | 8.79E-03 | 1.66E-02 |
| *LOC100506725* | 0.072 | 2.61 | 9.02E-03 | 1.66E-02 |
| *ARRDC2* | 0.072 | 2.61 | 9.12E-03 | 1.66E-02 |
| *ARHGEF1* | 0.072 | 2.60 | 9.31E-03 | 1.66E-02 |
| *ADAMTS14* | 0.071 | 2.60 | 9.37E-03 | 1.66E-02 |
| *TNS2* | 0.071 | 2.59 | 9.77E-03 | 1.70E-02 |
| *SCRG1* | 0.071 | 2.58 | 9.99E-03 | 1.71E-02 |
| *FNTA* | 0.070 | 2.56 | 1.04E-02 | 1.75E-02 |
| *CTNNA3* | 0.070 | 2.55 | 1.07E-02 | 1.78E-02 |

| | | | | |
|---|---|---|---|---|
| *HIPK2* | 0.069 | 2.49 | 1.27E-02 | 2.07E-02 |
| *PRKCQ* | 0.068 | 2.49 | 1.29E-02 | 2.07E-02 |
| *PAIP2B* | 0.068 | 2.46 | 1.39E-02 | 2.20E-02 |
| *SSFA2* | 0.067 | 2.45 | 1.44E-02 | 2.24E-02 |
| *CHST6* | 0.067 | 2.44 | 1.47E-02 | 2.25E-02 |
| *CDH20* | 0.067 | 2.43 | 1.51E-02 | 2.28E-02 |
| *TMEM140* | 0.067 | 2.42 | 1.54E-02 | 2.29E-02 |
| *TMTC4* | 0.066 | 2.40 | 1.63E-02 | 2.35E-02 |
| *CMTM5* | 0.066 | 2.40 | 1.65E-02 | 2.35E-02 |
| *PHF10* | 0.066 | 2.40 | 1.65E-02 | 2.35E-02 |
| *S100B* | 0.064 | 2.33 | 1.98E-02 | 2.77E-02 |
| *B3GNT9* | 0.063 | 2.29 | 2.19E-02 | 2.99E-02 |
| *MOBP* | 0.063 | 2.29 | 2.19E-02 | 2.99E-02 |
| *MYLK* | 0.063 | 2.28 | 2.25E-02 | 3.03E-02 |
| *RDH11* | 0.062 | 2.26 | 2.36E-02 | 3.14E-02 |
| *BCDIN3D* | 0.061 | 2.23 | 2.55E-02 | 3.35E-02 |
| *AAMDC* | 0.061 | 2.22 | 2.64E-02 | 3.42E-02 |
| *QDPR* | 0.061 | 2.21 | 2.72E-02 | 3.47E-02 |
| *LOC100507487* | 0.061 | 2.20 | 2.76E-02 | 3.48E-02 |
| *LLGL2* | 0.060 | 2.18 | 2.94E-02 | 3.67E-02 |
| *TCFL5* | 0.058 | 2.11 | 3.46E-02 | 4.22E-02 |
| *FRMD5* | 0.058 | 2.11 | 3.47E-02 | 4.22E-02 |
| *PLEKHB1* | 0.056 | 2.03 | 4.22E-02 | 5.07E-02 |
| *IGFLR1* | 0.055 | 2.00 | 4.56E-02 | 5.42E-02 |
| *KATNA1* | 0.055 | 1.99 | 4.66E-02 | 5.47E-02 |
| *VKORC1* | 0.055 | 1.98 | 4.73E-02 | 5.50E-02 |
| *DENND5A* | 0.054 | 1.98 | 4.81E-02 | 5.52E-02 |
| *FAT1* | 0.053 | 1.93 | 5.40E-02 | 6.13E-02 |
| *SEPT4* | 0.052 | 1.91 | 5.64E-02 | 6.33E-02 |
| *PPIF* | 0.050 | 1.82 | 6.87E-02 | 7.62E-02 |
| *BCAR3* | 0.047 | 1.72 | 8.49E-02 | 9.32E-02 |
| *KLC1* | -0.047 | -1.71 | 8.81E-02 | 9.57E-02 |
| *C9orf72* | 0.046 | 1.67 | 9.49E-02 | 1.02E-01 |
| *TST* | 0.045 | 1.64 | 1.01E-01 | 1.08E-01 |
| *RCOR2* | 0.042 | 1.54 | 1.24E-01 | 1.30E-01 |
| *ICK* | 0.042 | 1.52 | 1.30E-01 | 1.35E-01 |
| *LAP3* | 0.034 | 1.22 | 2.21E-01 | 2.28E-01 |
| *RGS1* | 0.031 | 1.12 | 2.63E-01 | 2.68E-01 |
| *SESTD1* | 0.017 | 0.62 | 5.33E-01 | 5.39E-01 |
| *FAM19A4* | 0.015 | 0.54 | 5.90E-01 | 5.90E-01 |

#### Supplementary Table 15a: Sensitivity analyses comprising Limma linear model statistics for normative spatial expression of the hypothesis-driven transcripts in the early-20s male donor.

| **Transcript** | ***ß*** | ***t*** | ***p*** | **Adjusted *p*** |
|---|---|---|---|---|
| *NLGN4X* | -0.034 | -3.03 | 4.02E-03 | 1.53E-01 |
| *S100A6* | 0.029 | 1.92 | 6.15E-02 | 8.70E-01 |
| *TARDBP* | 0.025 | 1.79 | 7.96E-02 | 8.70E-01 |
| *MET* | 0.010 | 1.48 | 1.46E-01 | 8.70E-01 |
| *CALB2* | -0.009 | -1.24 | 2.22E-01 | 8.70E-01 |
| *VCP* | -0.014 | -1.22 | 2.28E-01 | 8.70E-01 |
| *CNTNAP2* | 0.021 | 1.11 | 2.71E-01 | 8.70E-01 |
| *TCF7L2* | -0.004 | -1.01 | 3.18E-01 | 8.70E-01 |
| *FUS* | -0.023 | -1.00 | 3.25E-01 | 8.70E-01 |
| *GAD1* | -0.032 | -0.99 | 3.29E-01 | 8.70E-01 |
| *NES* | 0.018 | 0.94 | 3.53E-01 | 8.70E-01 |
| *MAPT* | -0.018 | -0.92 | 3.61E-01 | 8.70E-01 |
| *CALB1* | -0.025 | -0.92 | 3.61E-01 | 8.70E-01 |
| *HES1* | 0.010 | 0.80 | 4.28E-01 | 8.70E-01 |
| *OPTN* | 0.013 | 0.73 | 4.68E-01 | 8.70E-01 |
| *DLX1* | 0.003 | 0.70 | 4.85E-01 | 8.70E-01 |
| *CHCHD10* | -0.017 | -0.69 | 4.93E-01 | 8.70E-01 |
| *TMEM106B* | -0.007 | -0.69 | 4.96E-01 | 8.70E-01 |
| *RGS16* | -0.006 | -0.64 | 5.25E-01 | 8.70E-01 |
| *CEACAM21* | -0.014 | -0.63 | 5.33E-01 | 8.70E-01 |
| *SQSTM1* | 0.012 | 0.62 | 5.41E-01 | 8.70E-01 |
| *SLC32A1* | -0.024 | -0.61 | 5.46E-01 | 8.70E-01 |
| *SLC24A5* | -0.009 | -0.60 | 5.50E-01 | 8.70E-01 |
| *TBK1* | -0.012 | -0.55 | 5.87E-01 | 8.70E-01 |
| *DLX2* | 0.006 | 0.53 | 5.97E-01 | 8.70E-01 |
| *LHX2* | 0.008 | 0.51 | 6.13E-01 | 8.70E-01 |
| *SOX2* | 0.005 | 0.44 | 6.64E-01 | 8.70E-01 |
| *C9orf72* | -0.008 | -0.43 | 6.68E-01 | 8.70E-01 |
| *EWSR1* | -0.007 | -0.43 | 6.72E-01 | 8.70E-01 |
| *CHMP2B* | 0.008 | 0.41 | 6.87E-01 | 8.70E-01 |
| *SLC17A6* | -0.003 | -0.32 | 7.47E-01 | 9.16E-01 |
| *CCNF* | 0.006 | 0.29 | 7.75E-01 | 9.21E-01 |
| *PVALB* | 0.002 | 0.17 | 8.64E-01 | 9.56E-01 |
| *GRN* | -0.002 | -0.15 | 8.79E-01 | 9.56E-01 |
| *TIA1* | -0.003 | -0.15 | 8.80E-01 | 9.56E-01 |
| *TAF15* | 0.001 | 0.08 | 9.39E-01 | 9.89E-01 |
| *FOXP2* | -0.001 | -0.04 | 9.65E-01 | 9.89E-01 |
| *NRGN* | 0.000 | 0.01 | 9.89E-01 | 9.89E-01 |

Supplementary Table 15b: Sensitivity analyses comprising Limma linear model statistics for normative spatial expression of the hypothesis-driven transcripts in the early-30s male donor.

| **Transcript** | ***ß*** | ***t*** | ***p*** | **Adjusted *p*** |
|---|---|---|---|---|
| *TBK1* | -0.125 | -2.49 | 2.01E-02 | 3.33E-01 |
| *NES* | 0.068 | 2.08 | 4.89E-02 | 3.33E-01 |
| *S100A6* | 0.113 | 2.04 | 5.26E-02 | 3.33E-01 |
| *CCNF* | -0.081 | -2.02 | 5.48E-02 | 3.33E-01 |
| *SLC24A5* | -0.028 | -1.93 | 6.60E-02 | 3.33E-01 |
| *SLC32A1* | 0.098 | 1.91 | 6.86E-02 | 3.33E-01 |
| *TAF15* | 0.057 | 1.83 | 8.03E-02 | 3.33E-01 |
| *HES1* | 0.055 | 1.79 | 8.61E-02 | 3.33E-01 |
| *CHCHD10* | -0.064 | -1.78 | 8.79E-02 | 3.33E-01 |
| *SQSTM1* | 0.065 | 1.65 | 1.13E-01 | 3.33E-01 |
| *FOXP2* | -0.054 | -1.64 | 1.14E-01 | 3.33E-01 |
| *CNTNAP2* | -0.071 | -1.63 | 1.16E-01 | 3.33E-01 |
| *NLGN4X* | -0.060 | -1.57 | 1.30E-01 | 3.33E-01 |
| *OPTN* | 0.037 | 1.56 | 1.31E-01 | 3.33E-01 |
| *MET* | 0.050 | 1.56 | 1.32E-01 | 3.33E-01 |
| *PVALB* | -0.067 | -1.32 | 1.98E-01 | 4.59E-01 |
| *RGS16* | -0.055 | -1.30 | 2.05E-01 | 4.59E-01 |
| *MAPT* | 0.034 | 1.23 | 2.33E-01 | 4.75E-01 |
| *CALB1* | -0.057 | -1.21 | 2.38E-01 | 4.75E-01 |
| *SOX2* | 0.035 | 1.17 | 2.55E-01 | 4.84E-01 |
| *TCF7L2* | -0.049 | -1.11 | 2.80E-01 | 4.88E-01 |
| *CHMP2B* | 0.045 | 1.10 | 2.83E-01 | 4.88E-01 |
| *LHX2* | 0.027 | 0.98 | 3.35E-01 | 5.45E-01 |
| *CALB2* | 0.054 | 0.97 | 3.44E-01 | 5.45E-01 |
| *SLC17A6* | -0.046 | -0.87 | 3.93E-01 | 5.97E-01 |
| *TMEM106B* | -0.042 | -0.83 | 4.13E-01 | 6.04E-01 |
| *TIA1* | 0.019 | 0.75 | 4.64E-01 | 6.36E-01 |
| *C9orf72* | 0.015 | 0.74 | 4.69E-01 | 6.36E-01 |
| *GAD1* | 0.015 | 0.56 | 5.81E-01 | 7.51E-01 |
| *CEACAM21* | -0.019 | -0.54 | 5.93E-01 | 7.51E-01 |
| *GRN* | 0.007 | 0.46 | 6.46E-01 | 7.92E-01 |
| *VCP* | 0.004 | 0.32 | 7.52E-01 | 8.78E-01 |
| *EWSR1* | 0.013 | 0.27 | 7.87E-01 | 8.78E-01 |
| *NRGN* | 0.008 | 0.27 | 7.91E-01 | 8.78E-01 |
| *FUS* | -0.010 | -0.25 | 8.08E-01 | 8.78E-01 |
| *TARDBP* | 0.004 | 0.16 | 8.74E-01 | 9.23E-01 |
| *DLX2* | 0.001 | 0.04 | 9.67E-01 | 9.78E-01 |
| *DLX1* | 0.001 | 0.03 | 9.78E-01 | 9.78E-01 |

Supplementary Table 15c: Sensitivity analyses comprising Limma linear model statistics for normative spatial expression of the hypothesis-driven transcripts in the late-30s male donor.

| Transcript | *ß* | *t* | *p* | Adjusted *p* |
|---|---|---|---|---|
| *CALB1* | -0.098 | -3.88 | 2.73E-04 | 1.04E-02 |
| *NLGN4X* | -0.098 | -3.25 | 1.91E-03 | 3.64E-02 |
| *S100A6* | 0.081 | 3.04 | 3.52E-03 | 4.46E-02 |
| *TBK1* | -0.078 | -2.92 | 4.93E-03 | 4.69E-02 |
| *HES1* | 0.055 | 2.65 | 1.02E-02 | 7.79E-02 |
| *NES* | 0.070 | 2.55 | 1.34E-02 | 8.50E-02 |
| *CNTNAP2* | -0.056 | -2.40 | 1.94E-02 | 1.05E-01 |
| *CHMP2B* | 0.048 | 2.29 | 2.54E-02 | 1.21E-01 |
| *TARDBP* | 0.035 | 1.99 | 5.13E-02 | 2.17E-01 |
| *SLC24A5* | 0.043 | 1.88 | 6.52E-02 | 2.35E-01 |
| *SQSTM1* | 0.050 | 1.86 | 6.79E-02 | 2.35E-01 |
| *TIA1* | 0.047 | 1.73 | 8.91E-02 | 2.82E-01 |
| *GRN* | 0.033 | 1.64 | 1.06E-01 | 3.10E-01 |
| *OPTN* | 0.037 | 1.54 | 1.28E-01 | 3.35E-01 |
| *TCF7L2* | -0.045 | -1.53 | 1.32E-01 | 3.35E-01 |
| *TAF15* | 0.030 | 1.37 | 1.77E-01 | 3.73E-01 |
| *GAD1* | -0.040 | -1.35 | 1.83E-01 | 3.73E-01 |
| *C9orf72* | 0.028 | 1.34 | 1.85E-01 | 3.73E-01 |
| *EWSR1* | 0.033 | 1.34 | 1.86E-01 | 3.73E-01 |
| *NRGN* | -0.028 | -1.30 | 1.97E-01 | 3.75E-01 |
| *CHCHD10* | -0.020 | -0.86 | 3.93E-01 | 7.12E-01 |
| *SOX2* | 0.017 | 0.78 | 4.36E-01 | 7.29E-01 |
| *RGS16* | -0.024 | -0.71 | 4.80E-01 | 7.29E-01 |
| *CEACAM21* | 0.015 | 0.71 | 4.83E-01 | 7.29E-01 |
| *LHX2* | -0.017 | -0.70 | 4.87E-01 | 7.29E-01 |
| *SLC32A1* | -0.020 | -0.68 | 4.99E-01 | 7.29E-01 |
| *FUS* | 0.013 | 0.57 | 5.72E-01 | 8.05E-01 |
| *SLC17A6* | -0.013 | -0.37 | 7.15E-01 | 8.86E-01 |
| *MAPT* | -0.007 | -0.37 | 7.16E-01 | 8.86E-01 |
| *MET* | 0.009 | 0.36 | 7.21E-01 | 8.86E-01 |
| *CALB2* | -0.008 | -0.33 | 7.44E-01 | 8.86E-01 |
| *DLX2* | 0.006 | 0.30 | 7.62E-01 | 8.86E-01 |
| *DLX1* | 0.007 | 0.29 | 7.69E-01 | 8.86E-01 |
| *CCNF* | -0.004 | -0.17 | 8.69E-01 | 9.71E-01 |
| *PVALB* | 0.002 | 0.09 | 9.32E-01 | 9.91E-01 |
| *VCP* | -0.001 | -0.04 | 9.65E-01 | 9.91E-01 |
| *TMEM106B* | -0.001 | -0.03 | 9.76E-01 | 9.91E-01 |
| *FOXP2* | 0.0003 | 0.01 | 9.91E-01 | 9.91E-01 |

Supplementary Table 15d: Sensitivity analyses comprising Limma linear model statistics for normative spatial expression of the hypothesis-driven transcripts in the female donor.

| Transcript | *ß* | *t* | *p* | Adjusted *p* |
|---|---|---|---|---|
| *S100A6* | 0.160 | 2.98 | 6.02E-03 | 8.36E-02 |
| *CHMP2B* | 0.150 | 2.91 | 7.16E-03 | 8.36E-02 |
| *NES* | 0.160 | 2.79 | 9.67E-03 | 8.36E-02 |
| *SLC32A1* | 0.195 | 2.69 | 1.20E-02 | 8.36E-02 |
| *CALB2* | 0.138 | 2.65 | 1.34E-02 | 8.36E-02 |
| *TCF7L2* | 0.176 | 2.60 | 1.50E-02 | 8.36E-02 |
| *GRN* | 0.124 | 2.59 | 1.54E-02 | 8.36E-02 |
| *CHCHD10* | 0.146 | 2.53 | 1.78E-02 | 8.43E-02 |
| *EWSR1* | -0.155 | -2.39 | 2.40E-02 | 8.66E-02 |
| *MET* | 0.086 | 2.37 | 2.54E-02 | 8.66E-02 |
| *SOX2* | 0.117 | 2.33 | 2.76E-02 | 8.66E-02 |
| *RGS16* | 0.140 | 2.33 | 2.79E-02 | 8.66E-02 |
| *TMEM106B* | 0.095 | 2.29 | 3.02E-02 | 8.66E-02 |
| *MAPT* | -0.088 | -2.26 | 3.23E-02 | 8.66E-02 |
| *PVALB* | 0.153 | 2.22 | 3.49E-02 | 8.66E-02 |
| *FUS* | -0.097 | -2.20 | 3.64E-02 | 8.66E-02 |
| *C9orf72* | 0.087 | 2.13 | 4.25E-02 | 9.49E-02 |
| *FOXP2* | 0.134 | 2.03 | 5.26E-02 | 1.05E-01 |
| *NRGN* | -0.102 | -2.02 | 5.36E-02 | 1.05E-01 |
| *HES1* | 0.075 | 2.00 | 5.53E-02 | 1.05E-01 |
| *SLC17A6* | 0.156 | 1.95 | 6.16E-02 | 1.12E-01 |
| *TIA1* | -0.094 | -1.93 | 6.49E-02 | 1.12E-01 |
| *SQSTM1* | 0.071 | 1.83 | 7.78E-02 | 1.24E-01 |
| *GAD1* | 0.082 | 1.83 | 7.82E-02 | 1.24E-01 |
| *TAF15* | -0.065 | -1.74 | 9.42E-02 | 1.43E-01 |
| *LHX2* | -0.069 | -1.66 | 1.08E-01 | 1.54E-01 |
| *CEACAM21* | 0.062 | 1.66 | 1.09E-01 | 1.54E-01 |
| *TBK1* | 0.088 | 1.60 | 1.21E-01 | 1.61E-01 |
| *CALB1* | -0.057 | -1.59 | 1.23E-01 | 1.61E-01 |
| *OPTN* | 0.040 | 1.51 | 1.44E-01 | 1.82E-01 |
| *CNTNAP2* | 0.064 | 1.31 | 2.02E-01 | 2.43E-01 |
| *DLX1* | -0.047 | -1.30 | 2.05E-01 | 2.43E-01 |
| *NLGN4X* | -0.059 | -1.20 | 2.41E-01 | 2.77E-01 |
| *TARDBP* | -0.044 | -1.08 | 2.91E-01 | 3.25E-01 |
| *VCP* | -0.022 | -0.65 | 5.23E-01 | 5.66E-01 |
| *DLX2* | -0.022 | -0.63 | 5.36E-01 | 5.66E-01 |
| *SLC24A5* | -0.014 | -0.34 | 7.39E-01 | 7.40E-01 |
| *CCNF* | -0.017 | -0.34 | 7.40E-01 | 7.40E-01 |

Supplementary Table 15e: Sensitivity analyses comprising Limma linear model statistics for normative spatial expression of the hypothesis-driven transcripts in the first mid-to-late fifties male donor.

| **Transcript** | ***ß*** | ***t*** | ***p*** | **Adjusted *p*** |
|---|---|---|---|---|
| *CEACAM21* | 0.082 | 1.99 | 5.79E-02 | 9.65E-01 |
| *S100A6* | -0.064 | -1.66 | 1.09E-01 | 9.65E-01 |
| *CHMP2B* | 0.062 | 1.63 | 1.17E-01 | 9.65E-01 |
| *PVALB* | 0.060 | 1.21 | 2.39E-01 | 9.65E-01 |
| *HES1* | -0.039 | -1.16 | 2.58E-01 | 9.65E-01 |
| *SLC32A1* | -0.055 | -1.09 | 2.86E-01 | 9.65E-01 |
| *TMEM106B* | 0.055 | 1.08 | 2.93E-01 | 9.65E-01 |
| *TBK1* | 0.074 | 1.04 | 3.11E-01 | 9.65E-01 |
| *SQSTM1* | 0.043 | 1.02 | 3.18E-01 | 9.65E-01 |
| *FUS* | -0.037 | -1.01 | 3.23E-01 | 9.65E-01 |
| *EWSR1* | -0.042 | -0.91 | 3.70E-01 | 9.65E-01 |
| *TARDBP* | 0.034 | 0.90 | 3.77E-01 | 9.65E-01 |
| *CCNF* | -0.033 | -0.89 | 3.80E-01 | 9.65E-01 |
| *NLGN4X* | 0.043 | 0.87 | 3.94E-01 | 9.65E-01 |
| *MAPT* | -0.028 | -0.73 | 4.75E-01 | 9.65E-01 |
| *DLX2* | 0.025 | 0.52 | 6.09E-01 | 9.65E-01 |
| *VCP* | -0.008 | -0.46 | 6.47E-01 | 9.65E-01 |
| *GAD1* | 0.019 | 0.41 | 6.89E-01 | 9.65E-01 |
| *NRGN* | -0.024 | -0.40 | 6.90E-01 | 9.65E-01 |
| *C9orf72* | 0.021 | 0.38 | 7.04E-01 | 9.65E-01 |
| *RGS16* | 0.027 | 0.34 | 7.34E-01 | 9.65E-01 |
| *DLX1* | 0.015 | 0.34 | 7.39E-01 | 9.65E-01 |
| *TCF7L2* | -0.021 | -0.26 | 7.94E-01 | 9.65E-01 |
| *CALB2* | -0.015 | -0.26 | 7.95E-01 | 9.65E-01 |
| *SLC17A6* | -0.020 | -0.26 | 8.01E-01 | 9.65E-01 |
| *LHX2* | -0.014 | -0.25 | 8.05E-01 | 9.65E-01 |
| *CALB1* | -0.013 | -0.23 | 8.20E-01 | 9.65E-01 |
| *CNTNAP2* | 0.007 | 0.23 | 8.20E-01 | 9.65E-01 |
| *TIA1* | -0.010 | -0.22 | 8.30E-01 | 9.65E-01 |
| *GRN* | 0.004 | 0.18 | 8.55E-01 | 9.65E-01 |
| *OPTN* | -0.006 | -0.15 | 8.84E-01 | 9.65E-01 |
| *SLC24A5* | 0.004 | 0.13 | 8.98E-01 | 9.65E-01 |
| *SOX2* | -0.005 | -0.09 | 9.32E-01 | 9.65E-01 |
| *MET* | -0.005 | -0.08 | 9.35E-01 | 9.65E-01 |
| *FOXP2* | -0.005 | -0.08 | 9.36E-01 | 9.65E-01 |
| *NES* | 0.003 | 0.08 | 9.39E-01 | 9.65E-01 |
| *CHCHD10* | 0.003 | 0.08 | 9.40E-01 | 9.65E-01 |
| *TAF15* | 0.001 | 0.02 | 9.86E-01 | 9.86E-01 |

Supplementary Table 15f: Sensitivity analyses comprising Limma linear model statistics for normative spatial expression of the hypothesis-driven transcripts in the second mid-to-late fifties male donor.

| **Transcript** | ***ß*** | ***t*** | ***p*** | **Adjusted *p*** |
|---|---|---|---|---|
| *HES1* | 0.059 | 2.91 | 7.72E-03 | 2.09E-01 |
| *NLGN4X* | -0.086 | -2.76 | 1.10E-02 | 2.09E-01 |
| *C9orf72* | -0.071 | -2.48 | 2.09E-02 | 2.09E-01 |
| *S100A6* | 0.089 | 2.41 | 2.41E-02 | 2.09E-01 |
| *TMEM106B* | -0.091 | -2.33 | 2.89E-02 | 2.09E-01 |
| *OPTN* | 0.070 | 2.19 | 3.83E-02 | 2.09E-01 |
| *TBK1* | -0.087 | -2.19 | 3.85E-02 | 2.09E-01 |
| *EWSR1* | -0.071 | -2.11 | 4.60E-02 | 2.16E-01 |
| *CALB1* | -0.100 | -2.05 | 5.13E-02 | 2.16E-01 |
| *CALB2* | 0.052 | 1.79 | 8.56E-02 | 3.25E-01 |
| *MET* | 0.036 | 1.72 | 9.82E-02 | 3.39E-01 |
| *TCF7L2* | -0.035 | -1.54 | 1.37E-01 | 4.35E-01 |
| *RGS16* | -0.033 | -1.41 | 1.70E-01 | 4.74E-01 |
| *SOX2* | -0.025 | -1.40 | 1.75E-01 | 4.74E-01 |
| *NRGN* | 0.030 | 1.17 | 2.55E-01 | 6.22E-01 |
| *CHMP2B* | 0.047 | 1.15 | 2.62E-01 | 6.22E-01 |
| *VCP* | 0.019 | 1.04 | 3.07E-01 | 6.87E-01 |
| *MAPT* | -0.046 | -1.00 | 3.28E-01 | 6.93E-01 |
| *GAD1* | -0.039 | -0.90 | 3.79E-01 | 7.59E-01 |
| *PVALB* | -0.009 | -0.83 | 4.12E-01 | 7.84E-01 |
| *CHCHD10* | -0.016 | -0.75 | 4.59E-01 | 8.15E-01 |
| *DLX2* | 0.015 | 0.73 | 4.72E-01 | 8.15E-01 |
| *SLC24A5* | 0.018 | 0.62 | 5.42E-01 | 8.88E-01 |
| *CEACAM21* | -0.028 | -0.57 | 5.74E-01 | 8.88E-01 |
| *CNTNAP2* | 0.019 | 0.55 | 5.90E-01 | 8.88E-01 |
| *FOXP2* | 0.016 | 0.52 | 6.08E-01 | 8.88E-01 |
| *NES* | 0.016 | 0.45 | 6.57E-01 | 9.22E-01 |
| *TAF15* | 0.008 | 0.42 | 6.79E-01 | 9.22E-01 |
| *TIA1* | 0.015 | 0.37 | 7.18E-01 | 9.23E-01 |
| *FUS* | 0.012 | 0.33 | 7.44E-01 | 9.23E-01 |
| *LHX2* | 0.009 | 0.32 | 7.53E-01 | 9.23E-01 |
| *SQSTM1* | 0.006 | 0.23 | 8.24E-01 | 9.78E-01 |
| *SLC17A6* | -0.007 | -0.17 | 8.66E-01 | 9.86E-01 |
| *GRN* | 0.003 | 0.14 | 8.86E-01 | 9.86E-01 |
| *DLX1* | 0.002 | 0.09 | 9.26E-01 | 9.86E-01 |
| *SLC32A1* | -0.003 | -0.06 | 9.53E-01 | 9.86E-01 |
| *TARDBP* | 0.001 | 0.03 | 9.72E-01 | 9.86E-01 |
| *CCNF* | -0.001 | -0.02 | 9.86E-01 | 9.86E-01 |

Supplementary Table 15g: Sensitivity analyses comprising Limma linear model statistics for normative spatial expression of the hypothesis-driven transcripts in the two younger AHBA donors in aggregate, and (H) four older AHBA donors in aggregate.

| Transcript | *ß* | *t* | *p* | Adjusted *p* |
|---|---|---|---|---|
| *NLGN4X* | -0.035 | -2.80 | 7.41E-03 | 2.81E-01 |
| *NES* | 0.031 | 2.17 | 3.48E-02 | 4.98E-01 |
| *S100A6* | 0.042 | 2.12 | 3.93E-02 | 4.98E-01 |
| *CALB1* | -0.042 | -1.93 | 5.90E-02 | 5.61E-01 |
| *SQSTM1* | 0.025 | 1.69 | 9.70E-02 | 7.37E-01 |
| *MET* | 0.016 | 1.47 | 1.49E-01 | 8.91E-01 |
| *TBK1* | -0.025 | -1.37 | 1.77E-01 | 8.91E-01 |
| *CEACAM21* | -0.022 | -1.15 | 2.56E-01 | 8.91E-01 |
| *VCP* | -0.012 | -0.99 | 3.25E-01 | 8.91E-01 |
| *HES1* | 0.014 | 0.98 | 3.30E-01 | 8.91E-01 |
| *FUS* | -0.019 | -0.90 | 3.72E-01 | 8.91E-01 |
| *RGS16* | -0.012 | -0.89 | 3.76E-01 | 8.91E-01 |
| *OPTN* | 0.015 | 0.89 | 3.77E-01 | 8.91E-01 |
| *TCF7L2* | -0.009 | -0.85 | 4.00E-01 | 8.91E-01 |
| *GAD1* | -0.019 | -0.85 | 4.01E-01 | 8.91E-01 |
| *LHX2* | 0.012 | 0.83 | 4.09E-01 | 8.91E-01 |
| *C9orf72* | 0.011 | 0.73 | 4.70E-01 | 8.91E-01 |
| *TMEM106B* | -0.008 | -0.72 | 4.78E-01 | 8.91E-01 |
| *SLC24A5* | -0.010 | -0.70 | 4.86E-01 | 8.91E-01 |
| *CHMP2B* | 0.013 | 0.61 | 5.43E-01 | 8.91E-01 |
| *FOXP2* | -0.013 | -0.60 | 5.51E-01 | 8.91E-01 |
| *SLC17A6* | -0.007 | -0.54 | 5.89E-01 | 8.91E-01 |
| *DLX2* | 0.004 | 0.52 | 6.04E-01 | 8.91E-01 |
| *TIA1* | 0.008 | 0.50 | 6.22E-01 | 8.91E-01 |
| *PVALB* | -0.007 | -0.48 | 6.34E-01 | 8.91E-01 |
| *TARDBP* | -0.006 | -0.46 | 6.49E-01 | 8.91E-01 |
| *GRN* | -0.005 | -0.45 | 6.53E-01 | 8.91E-01 |
| *CHCHD10* | -0.008 | -0.44 | 6.63E-01 | 8.91E-01 |
| *TAF15* | 0.006 | 0.38 | 7.06E-01 | 8.91E-01 |
| *SOX2* | 0.009 | 0.38 | 7.07E-01 | 8.91E-01 |
| *SLC32A1* | -0.010 | -0.31 | 7.58E-01 | 8.91E-01 |
| *NRGN* | 0.004 | 0.27 | 7.88E-01 | 8.91E-01 |
| *CALB2* | 0.005 | 0.26 | 8.00E-01 | 8.91E-01 |
| *CCNF* | 0.005 | 0.25 | 8.07E-01 | 8.91E-01 |
| *MAPT* | -0.004 | -0.23 | 8.21E-01 | 8.91E-01 |
| *DLX1* | 0.002 | 0.18 | 8.61E-01 | 8.94E-01 |
| *CNTNAP2* | 0.003 | 0.16 | 8.70E-01 | 8.94E-01 |

| | | | | |
|---|---|---|---|---|
| *EWSR1* | 0.001 | 0.10 | 9.23E-01 | 9.23E-01 |

#### Supplementary Table 15h: Sensitivity analyses comprising Limma linear model statistics for normative spatial expression of the hypothesis-driven transcripts in the four older AHBA donors in aggregate.

| **Transcript** | ***ß*** | ***t*** | ***p*** | **Adjusted *p*** |
|---|---|---|---|---|
| *CALB1* | -0.098 | -4.03 | 1.73E-04 | 6.56E-03 |
| *NES* | 0.059 | 2.59 | 1.22E-02 | 2.09E-01 |
| *C9orf72* | 0.046 | 2.47 | 1.65E-02 | 2.09E-01 |
| *SQSTM1* | 0.050 | 2.10 | 4.02E-02 | 2.82E-01 |
| *HES1* | 0.036 | 2.10 | 4.04E-02 | 2.82E-01 |
| *S100A6* | 0.049 | 1.90 | 6.20E-02 | 2.82E-01 |
| *NLGN4X* | -0.040 | -1.90 | 6.32E-02 | 2.82E-01 |
| *CHMP2B* | 0.032 | 1.88 | 6.57E-02 | 2.82E-01 |
| *SLC24A5* | 0.039 | 1.87 | 6.68E-02 | 2.82E-01 |
| *GAD1* | -0.035 | -1.65 | 1.05E-01 | 3.60E-01 |
| *OPTN* | 0.029 | 1.61 | 1.13E-01 | 3.60E-01 |
| *GRN* | 0.030 | 1.61 | 1.14E-01 | 3.60E-01 |
| *TIA1* | 0.030 | 1.53 | 1.32E-01 | 3.69E-01 |
| *TCF7L2* | -0.041 | -1.47 | 1.48E-01 | 3.69E-01 |
| *TBK1* | -0.039 | -1.44 | 1.56E-01 | 3.69E-01 |
| *CEACAM21* | 0.026 | 1.43 | 1.59E-01 | 3.69E-01 |
| *NRGN* | -0.028 | -1.41 | 1.65E-01 | 3.69E-01 |
| *EWSR1* | 0.034 | 1.35 | 1.81E-01 | 3.82E-01 |
| *CNTNAP2* | -0.024 | -1.31 | 1.94E-01 | 3.88E-01 |
| *LHX2* | -0.027 | -1.22 | 2.26E-01 | 4.29E-01 |
| *MAPT* | -0.017 | -0.95 | 3.45E-01 | 5.92E-01 |
| *TAF15* | 0.020 | 0.95 | 3.48E-01 | 5.92E-01 |
| *RGS16* | -0.027 | -0.93 | 3.58E-01 | 5.92E-01 |
| *PVALB* | 0.019 | 0.86 | 3.95E-01 | 6.13E-01 |
| *TMEM106B* | -0.014 | -0.83 | 4.09E-01 | 6.13E-01 |
| *SLC32A1* | -0.020 | -0.81 | 4.20E-01 | 6.13E-01 |
| *SOX2* | 0.015 | 0.70 | 4.86E-01 | 6.64E-01 |
| *MET* | 0.013 | 0.67 | 5.06E-01 | 6.64E-01 |
| *CALB2* | -0.015 | -0.67 | 5.07E-01 | 6.64E-01 |
| *DLX1* | 0.012 | 0.64 | 5.26E-01 | 6.67E-01 |
| *FUS* | 0.010 | 0.48 | 6.34E-01 | 7.77E-01 |
| *SLC17A6* | -0.014 | -0.44 | 6.64E-01 | 7.88E-01 |
| *CHCHD10* | -0.008 | -0.38 | 7.09E-01 | 7.99E-01 |
| *CCNF* | -0.007 | -0.37 | 7.15E-01 | 7.99E-01 |
| *DLX2* | -0.004 | -0.26 | 7.95E-01 | 8.63E-01 |
| *VCP* | -0.002 | -0.08 | 9.33E-01 | 9.85E-01 |

| | | | | |
|---|---|---|---|---|
| *FOXP2* | 0.001 | 0.03 | 9.76E-01 | 9.94E-01 |
| *TARDBP* | -0.0001 | -0.01 | 9.94E-01 | 9.94E-01 |

#### Supplementary Table 16a: Limma linear model statistics for normative spatial expression of the transcripts of interest in thalamic structures of BrainSpan donors across pre-natal stages to young adulthood.

| Transcript | Log2 Fold Change | *t* | *p* | Adjusted *p* |
|---|---|---|---|---|
| *GAD1* | 3.86 | 8.80 | 1.02E-09 | 3.78E-08 |
| *CEACAM21* | 2.41 | 7.07 | 8.51E-08 | 1.57E-06 |
| *LHX2* | -2.86 | -6.39 | 5.24E-07 | 6.47E-06 |
| *CALB2* | 2.68 | 4.59 | 7.74E-05 | 7.16E-04 |
| *SLC32A1* | 1.61 | 3.49 | 1.57E-03 | 0.012 |
| *SQSTM1* | 0.61 | 2.49 | 0.019 | 0.116 |
| *DLX2* | 0.65 | 2.40 | 0.023 | 0.121 |
| *CCNF* | -0.54 | -2.26 | 0.031 | 0.144 |
| *NES* | -0.65 | -2.17 | 0.038 | 0.156 |
| *TIA1* | -0.66 | -2.10 | 0.044 | 0.158 |
| *DLX1* | 0.68 | 2.05 | 0.049 | 0.158 |
| *TCF7L2* | -0.90 | -2.03 | 0.051 | 0.158 |
| *MAPT* | 0.49 | 1.98 | 0.057 | 0.162 |
| *TMEM106B* | -0.73 | -1.81 | 0.080 | 0.211 |
| *FOXP2* | -0.81 | -1.72 | 0.097 | 0.239 |
| *TARDBP* | -0.50 | -1.67 | 0.105 | 0.243 |
| *PVALB* | 1.07 | 1.57 | 0.126 | 0.270 |
| *C9orf72* | -0.49 | -1.55 | 0.132 | 0.270 |
| *NLGN4X* | 0.40 | 1.28 | 0.212 | 0.412 |
| *TBK1* | -0.32 | -1.18 | 0.247 | 0.436 |
| *FUS* | -0.25 | -1.18 | 0.248 | 0.436 |
| *OPTN* | 0.21 | 0.84 | 0.405 | 0.659 |
| *S100A6* | -0.21 | -0.84 | 0.410 | 0.659 |
| *VCP* | 0.17 | 0.78 | 0.441 | 0.666 |
| *CHCHD10* | 0.21 | 0.77 | 0.450 | 0.666 |
| *SLC24A5* | -0.20 | -0.65 | 0.521 | 0.742 |
| *CHMP2B* | -0.18 | -0.54 | 0.594 | 0.800 |
| *GRN* | 0.15 | 0.49 | 0.627 | 0.800 |
| *CALB1* | 0.24 | 0.47 | 0.639 | 0.800 |
| *MET* | 0.09 | 0.46 | 0.649 | 0.800 |
| *EWSR1* | 0.14 | 0.33 | 0.742 | 0.878 |
| *TAF15* | -0.07 | -0.31 | 0.759 | 0.878 |
| *NRGN* | 0.11 | 0.23 | 0.820 | 0.919 |
| *SLC17A6* | 0.06 | 0.18 | 0.857 | 0.919 |
| *RGS16* | -0.09 | -0.17 | 0.869 | 0.919 |
| *CNTNAP2* | -0.03 | -0.13 | 0.901 | 0.926 |

| | | | | |
|---|---|---|---|---|
| *HES1* | 0.03 | 0.07 | 0.947 | 0.947 |

#### Supplementary Table 16b: Limma linear model statistics for normative spatial expression of the transcripts of interest in occipital structures of BrainSpan donors across pre-natal stages to young adulthood.

| Transcript | Log2 Fold Change | *t* | *p* | Adjusted *p* |
|---|---|---|---|---|
| *CCNF* | -1.79 | -7.66 | 9.46E-09 | 3.50E-07 |
| *RGS16* | -2.23 | -6.18 | 6.24E-07 | 1.15E-05 |
| *TCF7L2* | -2.71 | -5.45 | 5.27E-06 | 6.50E-05 |
| *MAPT* | 1.62 | 5.14 | 1.28E-05 | 1.19E-04 |
| *HES1* | -3.22 | -4.43 | 1.02E-04 | 7.55E-04 |
| *NES* | -1.62 | -3.67 | 8.71E-04 | 5.37E-03 |
| *OPTN* | 1.07 | 3.29 | 2.41E-03 | 1.27E-02 |
| *CALB2* | -1.71 | -2.44 | 2.04E-02 | 0.09 |
| *DLX1* | 1.13 | 1.87 | 0.07 | 0.28 |
| *DLX2* | 0.66 | 1.81 | 0.08 | 0.28 |
| *SLC24A5* | 0.45 | 1.78 | 0.09 | 0.28 |
| *FOXP2* | -1.29 | -1.72 | 0.10 | 0.28 |
| *TAF15* | -0.64 | -1.69 | 0.10 | 0.28 |
| *GRN* | -0.78 | -1.61 | 0.12 | 0.31 |
| *SQSTM1* | 0.65 | 1.58 | 0.12 | 0.31 |
| *FUS* | -0.39 | -1.42 | 0.16 | 0.38 |
| *NRGN* | 0.95 | 1.26 | 0.22 | 0.47 |
| *CNTNAP2* | -0.84 | -1.20 | 0.24 | 0.49 |
| *TMEM106B* | -0.60 | -1.11 | 0.28 | 0.51 |
| *TBK1* | 0.36 | 1.09 | 0.28 | 0.51 |
| *MET* | 1.08 | 1.05 | 0.30 | 0.51 |
| *LHX2* | -0.74 | -1.02 | 0.31 | 0.51 |
| *TARDBP* | -0.30 | -0.96 | 0.34 | 0.51 |
| *GAD1* | 0.71 | 0.96 | 0.34 | 0.51 |
| *CALB1* | 1.01 | 0.95 | 0.35 | 0.51 |
| *S100A6* | 0.32 | 0.90 | 0.37 | 0.51 |
| *SLC32A1* | 0.63 | 0.88 | 0.38 | 0.51 |
| *CHMP2B* | 0.43 | 0.84 | 0.41 | 0.51 |
| *EWSR1* | 0.56 | 0.83 | 0.41 | 0.51 |
| *CEACAM21* | 0.31 | 0.83 | 0.41 | 0.51 |
| *TIA1* | -0.26 | -0.56 | 0.58 | 0.68 |
| *VCP* | 0.16 | 0.55 | 0.58 | 0.68 |
| *PVALB* | -0.60 | -0.50 | 0.62 | 0.70 |
| *CHCHD10* | -0.19 | -0.47 | 0.64 | 0.70 |
| *NLGN4X* | 0.24 | 0.38 | 0.71 | 0.75 |
| *SLC17A6* | 0.05 | 0.05 | 0.96 | 0.97 |
| *C9orf72* | 0.02 | 0.04 | 0.97 | 0.97 |

Supplementary Table 17a: Spearman-rank correlations between *C9orf72* and the transcripts of interest in thalamic structures of BrainSpan donors across pre-natal stages to young adulthood.

| **Transcript** | **rho** | ***p*** | **Adjusted *p*** |
|---|---|---|---|
| *C9orf72* | 1 | 0 | 0 |
| *TBK1* | 0.74 | 1.31E-05 | 1.92E-04 |
| *CHMP2B* | 0.73 | 2.03E-05 | 1.92E-04 |
| *TMEM106B* | 0.73 | 2.08E-05 | 1.92E-04 |
| *TAF15* | -0.59 | 1.32E-03 | 9.75E-03 |
| *CALB2* | 0.50 | 6.93E-03 | 0.043 |
| *NLGN4X* | 0.48 | 9.82E-03 | 0.052 |
| *CALB1* | 0.44 | 0.022 | 0.100 |
| *EWSR1* | -0.42 | 0.026 | 0.107 |
| *SQSTM1* | 0.40 | 0.034 | 0.125 |
| *SLC17A6* | 0.36 | 0.059 | 0.199 |
| *TIA1* | -0.35 | 0.070 | 0.216 |
| *SLC24A5* | -0.32 | 0.098 | 0.278 |
| *MET* | -0.27 | 0.167 | 0.439 |
| *CNTNAP2* | 0.26 | 0.178 | 0.439 |
| *NES* | -0.24 | 0.223 | 0.515 |
| *CCNF* | 0.21 | 0.284 | 0.619 |
| *FOXP2* | 0.19 | 0.319 | 0.656 |
| *FUS* | -0.18 | 0.361 | 0.686 |
| *CHCHD10* | 0.18 | 0.371 | 0.686 |
| *DLX2* | 0.15 | 0.430 | 0.757 |
| *HES1* | -0.14 | 0.472 | 0.767 |
| *S100A6* | 0.14 | 0.477 | 0.767 |
| *GAD1* | -0.13 | 0.518 | 0.799 |
| *PVALB* | -0.10 | 0.609 | 0.858 |
| *CEACAM21* | -0.10 | 0.611 | 0.858 |
| *GRN* | 0.09 | 0.631 | 0.858 |
| *SLC32A1* | 0.09 | 0.663 | 0.858 |
| *TARDBP* | -0.08 | 0.687 | 0.858 |
| *VCP* | 0.08 | 0.702 | 0.858 |
| *MAPT* | -0.06 | 0.748 | 0.858 |
| *DLX1* | 0.06 | 0.752 | 0.858 |
| *NRGN* | 0.06 | 0.777 | 0.858 |
| *TCF7L2* | -0.05 | 0.788 | 0.858 |
| *LHX2* | -0.03 | 0.877 | 0.884 |
| *OPTN* | -0.03 | 0.879 | 0.884 |
| *RGS16* | 0.03 | 0.884 | 0.884 |

Supplementary Table 17b: Spearman-rank correlations between *C9orf72* and the transcripts of interest in occipital structures of BrainSpan donors across pre-natal stages to young adulthood.

| **Transcript** | **rho** | ***p*** | **Adjusted *p*** |
|---|---|---|---|
| *C9orf72* | 1 | 0 | 0 |
| *HES1* | -0.61 | 2.23E-04 | 4.13E-03 |
| *CHCHD10* | 0.49 | 4.24E-03 | 0.052 |
| *TBK1* | 0.45 | 8.92E-03 | 0.080 |
| *PVALB* | 0.44 | 0.011 | 0.080 |
| *FUS* | -0.37 | 0.036 | 0.211 |
| *TMEM106B* | 0.36 | 0.040 | 0.211 |
| *LHX2* | -0.32 | 0.073 | 0.319 |
| *VCP* | 0.31 | 0.078 | 0.319 |
| *CHMP2B* | 0.29 | 0.102 | 0.326 |
| *CEACAM21* | -0.29 | 0.103 | 0.326 |
| *NES* | -0.28 | 0.109 | 0.326 |
| *SLC17A6* | 0.28 | 0.114 | 0.326 |
| *RGS16* | -0.27 | 0.135 | 0.358 |
| *OPTN* | 0.24 | 0.186 | 0.459 |
| *TAF15* | -0.22 | 0.212 | 0.491 |
| *SLC24A5* | -0.21 | 0.243 | 0.508 |
| *S100A6* | -0.21 | 0.247 | 0.508 |
| *TCF7L2* | -0.20 | 0.270 | 0.526 |
| *GRN* | 0.17 | 0.337 | 0.623 |
| *CALB2* | 0.15 | 0.415 | 0.704 |
| *GAD1* | 0.14 | 0.432 | 0.704 |
| *NRGN* | -0.14 | 0.438 | 0.704 |
| *TARDBP* | -0.12 | 0.520 | 0.735 |
| *SQSTM1* | 0.11 | 0.531 | 0.735 |
| *EWSR1* | -0.11 | 0.538 | 0.735 |
| *DLX1* | -0.11 | 0.548 | 0.735 |
| *SLC32A1* | 0.10 | 0.562 | 0.735 |
| *MAPT* | 0.10 | 0.576 | 0.735 |
| *FOXP2* | 0.09 | 0.618 | 0.754 |
| *TIA1* | -0.09 | 0.632 | 0.754 |
| *CCNF* | 0.08 | 0.671 | 0.776 |
| *CALB1* | 0.06 | 0.756 | 0.847 |
| *NLGN4X* | -0.04 | 0.820 | 0.892 |
| *CNTNAP2* | -0.04 | 0.845 | 0.893 |
| *DLX2* | -0.01 | 0.940 | 0.952 |
| *MET* | 0.01 | 0.952 | 0.952 |
